# Factors associated with long-term opioid therapy discontinuation for people with chronic non-cancer pain in UK primary care: a population-based retrospective cohort study

**DOI:** 10.1101/2025.03.19.25324292

**Authors:** Qian Cai, Yun-Ting (Joyce) Huang, Thomas Allen, Charlotte Morris, Christos Grigoroglou, Evangelos Kontopantelis

## Abstract

**Objectives:** To identify factors associated with long-term opioid therapy (L-TOT) discontinuation in people with chronic non-cancer pain (CNCP).

**Design and setting:** Population-based retrospective cohort study using UK Clinical Practice Research Datalink Aurum data between 01/01/2000-31/12/2020.

**Population:** The study cohort comprised adults (≥18 years) with CNCP who were using L-TOT, defined as ≥3 opioid prescriptions within 90 days, or a ≥90 days of supply within the first year, excluding the first 30 days.

**Main outcome measures:** Discontinuation was defined as opioid-free for ≥180 days following an episode of L-TOT. Mixed-effects logistic models with a random intercept for general practice were used to identify sociodemographic, comorbidities, lifestyle and pharmacological factors associated with L-TOT discontinuation.

**Results:** Among 573,639 L-TOT users, 5.2% (n=29,589) discontinued (mean age 56.45±18.41, female 60.04%) within the first year. Factors significantly associated with a higher likelihood of L-TOT discontinuation included being Asian (adjusted odds ratio: 1.31, 95% confidence interval: 1.23 to 1.39) or Black (1.21, 1.13 to 1.31), a non-smoker (1.09, 1.05 to 1.12) or a former smoker (1.05, 1.02 to 1.09), living in the least deprived area (1.09, 1.04 to 1.14), using weak and short-acting opioids (1.50, 1.39 to 1.61), coexisting osteoarthritis (1.06, 1.02 to 1.11) or anxiety (1.04, 1.01 to 1.07), and concurrently using non-steroidal anti-inflammatory drugs (1.06, 1.03 to 1.09) or benzodiazepines (1.08, 1.05 to 1.11). Those younger (0.990, 0.988 to 0.992), with lower daily dose (0.981, 0.980 to 0.982), substance use disorder (0.77, 0.72 to 0.83), and taking gabapentinoids (0.83, 0.69 to 1.00) or antidepressants (0.89, 0.87 to 0.92) were less likely to discontinue L-TOT.

**Conclusions:** This study identified key characteristics of people with CNCP who are more likely to discontinue L-TOT in UK primary care. Such information can help to guide the development of targeted, personalised interventions to support safe and effective opioid discontinuation.

**What is already known on this topic:**

- Existing evidence on L-TOT discontinuation primarily comes from US-based studies, revealing several common associated factors, including younger age, mental health conditions, and substance use disorders.
- However, conclusions vary across studies due to differences in study populations and L-TOT definitions. Some studies link discontinuations to lower opioid doses, while others associate it with higher doses, concurrent use of benzodiazepine, or being unmarried.

**What this study adds:**

- Our study identified additional factors associated with a greater likelihood of L-TOT discontinuation that have not been previously reported, including being Asian or Black, a former or a non-smoker, living in less deprived areas, co-existing osteoarthritis or anxiety, and concurrent NSAIDs use.
- L-TOT discontinuation rates varied across regions, with the likelihood decreasing in more northern regions, compared to London. Addressing these geographical inequities through a fairer distribution of healthcare resources, particularly in underserved areas, could improve support for patients attempting to discontinue L-TOT.
- Demographics, comorbidities, medication profiles, and lifestyle factors should be considered when devising L-TOT discontinuation strategies among people with CNCP in UK primary care.

## Introduction

Long-term opioid therapy (L-TOT) has been associated with serious risks, including emergency department visits, ^1^ hospitalisation, ^1^ bone fractures, ^2, 3^ and opioid-related death.^4^ Our previous work found that in United Kingdom (UK) primary care, one in ten individuals who initiated opioid use for chronic non-cancer pain (CNCP) transitioned to L-TOT, and the discontinuation rate has significantly declined over the past two decades. ^5^ Despite clinical guidelines such as the Faculty of Pain Medicine’s ‘Tapering and Stopping’ ^6^ and Oxford University Hospitals’ ‘Guidance for Opioid Reduction in Primary Care’ ^7^ advocating for reducing opioid prescriptions, the practical implementation of opioid discontinuation remains challenging. Patients on L-TOT often develop physical and psychological dependence, and may fear unmanaged pain or withdrawal symptoms, making them reluctant to reduce opioid use. ^8^ Clinicians, also face several barriers, including limited consultation time, and restricted availability or accessibility of non-opioid pain management options. ^9^ Additionally, healthcare providers may lack confidence or training in reducing or discontinuing opioids, making it more difficult to follow guideline recommendations.

Given these challenges, understanding factors that contribute to successful L-TOT discontinuation is crucial for informing targeted discontinuation strategies for CNCP population in UK primary care. However, current evidence on this topic remains limited. Most available studies are conducted in the United States (US) with small sample sizes, and focus on specific populations such as veterans, that limit their generalisability to UK primary care. These studies have identified various determinants of L-TOT discontinuation, including both being younger and older in age, lower opioid dosage, concurrent benzodiazepine use, mental health disorders and unmarried status. ^10–16^ However, given the differences in healthcare systems, regulatory environments, prescribers’ behaviours and population characteristics, these findings may not be directly transferable to the UK’s primary care context.

This study aimed to identify sociodemographic characteristics, comorbidities, lifestyle and pharmacological factors that influence the likelihood of L-TOT discontinuation among a CNCP population, with a special focus within UK primary care settings, where the majority of opioid prescriptions are issued. ^17^

## Methods

### Study design and population

The study design was described in detail in our previous work. ^5^ Briefly, in this population-based retrospective cohort study, we used data from the Clinical Practice Research Datalink (CPRD) Aurum, a database of anonymised UK primary care electronic health records, ^18^ covering 20% of the national population (approximately 13 million active registrants). We included people aged ≥18 years on the index date (the first opioid prescription following a 12-month opioid-free period) with a diagnosis of CNCP (code list is provided in Supplementary Table S1) and had ≥ 1 episode of L-TOT between 1^st^ January 2000 and 31^st^ December 2020. L-TOT was defined as receiving ≥ 3 opioid prescriptions within a 90-day period, or a sum of opioid supply days lasting ≥ 90 days, within the first year of follow-up (excluding the first 30 days). ^19^ CNCP diagnosis could occur up to 6 months after or any time before the index date. Patients with a cancer diagnosis, except for non-melanoma skin cancer, in the previous ten years were excluded due to different drug utilisation patterns and different guidelines for opioid prescribing (Supplementary figure S1). To be eligible, patients were required to have been registered with the CPRD practice for at least 12 months and have available linked data for the Index of Multiple Deprivation (IMD) 2019, ^20^ a measure of socioeconomic status in England; the Hospital Episode Statistics Admitted Patient Care (HES APC) database, which contains records of hospital admissions; and the Office for National Statistics (ONS) mortality data, which records death registrations.

### Opioid use

All non-parental opioid analgesics (code list is provided in Supplementary Table S2) were included, except for methadone and sublingual buprenorphine, as their primary indication is for opioid addiction rather than pain management. Opioid drug exposure data were processed using the logic of a previously published drug preparation algorithm. ^19, 21^ Details of the relevant decisions are provided in Supplementary figure S2. Opioid doses were converted to oral morphine milligram equivalents (MME) using standardised conversion factors. ^22^

### Outcomes

L-TOT discontinuation was defined as no opioid prescription records for ≥ 180 days following a period of L-TOT.

### Potential factors

Patient characteristics (age, sex) and lifestyle choices (drinking and smoking status) were collected on the index date. Ethnicity data was obtained through the linkage with HES APC data. ^23^ Geographical regions included North East, North West, Yorkshire & the Humber, East Midlands, West Midlands, East of England, South West, South Central, South East Coast and London. Socioeconomic status was measured using the IMD 2019 ^20^ and categorised into quintiles (Q1-least deprived, Q5-most deprived). ^24^

Comorbidities were measured using the Charlson comorbidity index (CCI) score and categorised into three grades: mild (0-2); moderate (3-4); and severe (≥5). ^25^ The presence of other conditions, including substance use disorders (SUDs), alcohol dependence, anxiety, depression, schizophrenia, osteoarthritis, rheumatoid arthritis, and epilepsy, was identified within five years prior to the index date. We also identified concurrent medications including benzodiazepines, gabapentinoids, antidepressants, Z-drugs, muscle relaxants or non-steroidal anti-inflammatory drugs (NSAIDs) within five years prior to the index date.

### Statistical analysis

Mixed-effects logistic regression analysis, with a random intercept for general practice, was conducted to identify factors associated with L-TOT discontinuation. The model included all potential factors previously mentioned as independent variables, and L-TOT discontinuation as the dependent variable. For categorical variables (i.e., ethnicity, IMD, drinking status, and smoking status), missing data were categorised as a separate group as “unknown” and included in the regression models. All other data were complete. The results were reported as adjusted odds ratio (aOR) with 95% confidence intervals (CIs) for each factor. All analyses were conducted with Stata/MP 18.0.

### Sensitivity analysis

To assess the robustness of our results, we conducted two sensitivity analyses: 1) a Cox-proportional hazards regression to model the time to L-TOT discontinuation, with practice as random effect (shared frailty), and age, average daily dose, rheumatoid arthritis, anxiety, weak and short-acting opioids, gabapentinoids, antidepressants, and benzodiazepines as time dependent variables based on the result of a proportional hazards assumption test; 2) a competing-risks (Fine-Gray) proportional hazards regression, considering all-cause mortality (extracted from the ONS mortality dataset) as the competing event for L-TOT discontinuation.

### Patient and public involvement

We have engaged patient groups, including the PRIMER (Primary Care Research in Manchester Engagement Resource, https://sites.manchester.ac.uk/primer/) at the University of Manchester, to assist in developing our dissemination strategy.

## Results

### Patient Characteristics

This study identified 573,639 L-TOT users in the first-year follow-up after opioid initiation, of whom 5.2% (n=29,589) discontinued their L-TOT within the following one year (Supplementary figure S3). Table 1 summarised patient characteristics between L-TOT discontinuers and L-TOT users. In both groups, 80% were white and 60% were female. L-TOT discontinuers tended to be younger (mean age 56.42±18.41 years) and were prescribed lower daily dose (17.82±11.52 MME/day). The majority (>99%) of both groups received doses ≤50 MME/day and were prescribed with weak opioids or short-acting opioids. L-TOT discontinuers, descriptively compared to L-TOT users, were more frequently non-smokers, with anxiety, depression, lower CCI score (0-2), living in London or East of England, and concurrently using NSAIDs, muscle relaxants and benzodiazepines (Table 1).

**Table 1.** Demographics, comorbidity, concurrent drug, and opioid characteristics, among L-TOT users and L-TOT discontinuers between 01/01/2000 - 31/12/2020.

|  |  | L-TOT users (n= 544,050) | L-TOT discontinuers (n=29,589) |
| --- | --- | --- | --- |
| Sex | Female | 321,840 (59.16%) | 17,764 (60.04%) |
|  | Male | 222,210 (40.84%) | 11,825 (39.96%) |
| Age (mean±SD) |  | 59.30±18.27 | 56.45±18.41 |
| Age group | 18-29 years | 33,205 (6.10%) | 2,500 (8.45%) |
|  | 30-39 years | 58,519 (10.76%) | 3,774 (12.75%) |
|  | 40-49 years | 80,365 (14.77%) | 4,777 (16.14%) |
|  | 50-59 years | 90,177 (16.58%) | 4,967 (16.79%) |
|  | 60-69 years | 102,412 (18.82%) | 5,330 (18.01%) |
|  | 70-79 years | 96,173 (17.68%) | 4,791 (16.19%) |
|  | 80-89 years | 66,284 (12.18%) | 2,830 (9.56%) |
|  | ≥90 years | 16,915 (3.11%) | 620 (2.10%) |
| Ethnicity | White | 451,966 (83.07%) | 23,627 (79.85%) |
|  | Asian | 19,474 (3.58%) | 1,582 (5.35%) |
|  | Black | 12,044 (2.21%) | 907 (3.07%) |
|  | Mixed | 2,682 (0.49%) | 187 (0.63%) |
|  | Other | 5,639 (1.04%) | 358 (1.21%) |
|  | Unknown | 52,245 (9.60%) | 2,928 (9.90%) |
| IMD quintile | 5 (most deprived) | 129,752 (23.85%) | 6,866 (23.20%) |
|  | 4 | 110,654 (20.34%) | 6,059 (20.48%) |
|  | 3 | 101,009 (18.57%) | 5,592 (18.90%) |
|  | 2 | 101,925 (18.73%) | 5,432 (18.36%) |
|  | 1 (least deprived) | 89,918 (16.53%) | 5,011 (16.94%) |
|  | Unknown | 10,792 (1.98%) | 629 (2.13%) |
| Region | London | 67,346 (12.38%) | 4,322 (14.61%) |
|  | North East | 24,330 (4.47%) | 1,163 (3.93%) |
|  | North West | 122,552 (22.53%) | 6,517 (22.03%) |
|  | Yorkshire & the Humber | 23,405 (4.30%) | 1,152 (3.89%) |
|  | East Midlands | 13,101 (2.41%) | 728 (2.46%) |
|  | West Midlands | 95,689 (17.59%) | 5,147 (17.39%) |
|  | East of England | 22,153 (4.07%) | 1,368 (4.62%) |
|  | South West | 70,758 (13.01%) | 3,813 (12.89%) |
|  | South Central | 59,352 (10.91%) | 3,067 (10.37%) |
|  | South East Coast | 45,364 (8.34%) | 29,589 (7.81%) |
| Opioid characteristics |  |  |  |
|  | Average daily dose (MME/day) | 19.73±11.88 | 17.82±11.52 |

|  |  | **L-TOT users (n= 544,050)** | **L-TOT discontinuers (n=29,589)** |
| --- | --- | --- | --- |
|  | <50 MME/day | 539,204 (99.11%) | 29,415 (99.41%) |
|  | [50, 90) MME/day | 4,215 (0.77%) | 155 (0.52%) |
|  | [90,120) MME/day | 214 (0.04%) | 9 (0.03%) |
|  | >=120 MME/day | 417 (0.08%) | 10 (0.03%) |
|  | Weak opioids | 455,650 (83.75%) | 25,292 (85.48%) |
|  | Short-acting opioids | 520,138 (95.60%) | 28,683 (96.94%) |
| Drinking status | Current | 286,064 (52.58%) | 15,504 (52.40%) |
|  | Never | 92,098 (16.93%) | 4,837 (16.35%) |
|  | Former | 7,075 (1.30%) | 339 (1.15%) |
|  | Unknown | 158,813 (29.19%) | 8,909 (30.11%) |
| Smoking status | Current | 111,402 (20.48%) | 5,881 (19.88%) |
|  | Never | 254,768 (46.83%) | 14,452 (48.84%) |
|  | Former | 174,291 (32.04%) | 9,082 (30.69%) |
|  | Unknown | 3,589 (0.66%) | 174 (0.59%) |
| Comorbidity | RA | 20,620 (3.79%) | 1,000 (3.38%) |
|  | OA | 61,163 (11.24%) | 3,125 (10.56%) |
|  | Anxiety | 180,583 (33.19%) | 10,276 (34.73%) |
|  | Depression | 233,067 (42.84%) | 12,786 (43.21%) |
|  | Schizophrenia | 12,681 (2.33%) | 604 (2.04%) |
|  | Epilepsy | 16,771 (3.07%) | 842 (2.85%) |
|  | Substance Use Disorder | 20,662 (3.80%) | 873 (2.95%) |
|  | Alcohol dependence | 22,614 (4.16%) | 1,028 (3.47%) |
| CCI | High (≥5) | 71,897 (13.22%) | 3,675 (12.42%) |
|  | Medium (3-4) | 104,305 (19.17%) | 5,379 (18.18%) |
|  | Low (0-2) | 367,848 (67.61%) | 20,535 (69.40%) |
| Concurrent medications | Gabapentinoids | 2,980 (0.55%) | 120 (0.41%) |
|  | Antidepressants | 243,801 (44.81%) | 12,545 (42.40%) |
|  | Muscle relaxants | 6,302 (1.16%) | 423 (1.43%) |
|  | NSAIDs | 353,788 (65.03%) | 20,099 (67.96%) |
|  | Benzodiazepines | 123,889 (22.77%) | 6,915 (23.37%) |
|  | Z-drugs | 65,917 (12.12%) | 3,403 (11.50%) |

Note: IMD = Index of Multiple Deprivation; MME = Morphine Milligram Equivalents; RA = Rheumatoid Arthritis; OA = Osteoarthritis; CCI = Charlson Comorbidity Index; NSAIDs = Non-steroidal anti-inflammatory drugs; L-TOT users means those who did not discontinue opioids

### Factors associated with a higher likelihood of L-TOT discontinuation

Figure 1 presented the aOR and 95% CIs for each factor associated with L-TOT discontinuation. A higher likelihood of discontinuation was observed among Asians (aOR 1.31, 95% CI 1.23 to 1.39) and Blacks (aOR 1.21, 95% CI 1.13 to 1.31) compared to the White. Compared to current smokers, former smokers (aOR 1.05, 95% CI 1.02 to 1.09) and people who never smoked (aOR 1.09, 95% CI 1.05 to 1.12) were more likely to discontinue L-TOT. The use of weak and short-acting opioids significantly increased the likelihood of discontinuation by 50% (aOR 1.50, 95% CI 1.39 to 1.61).

**Figure 1.**
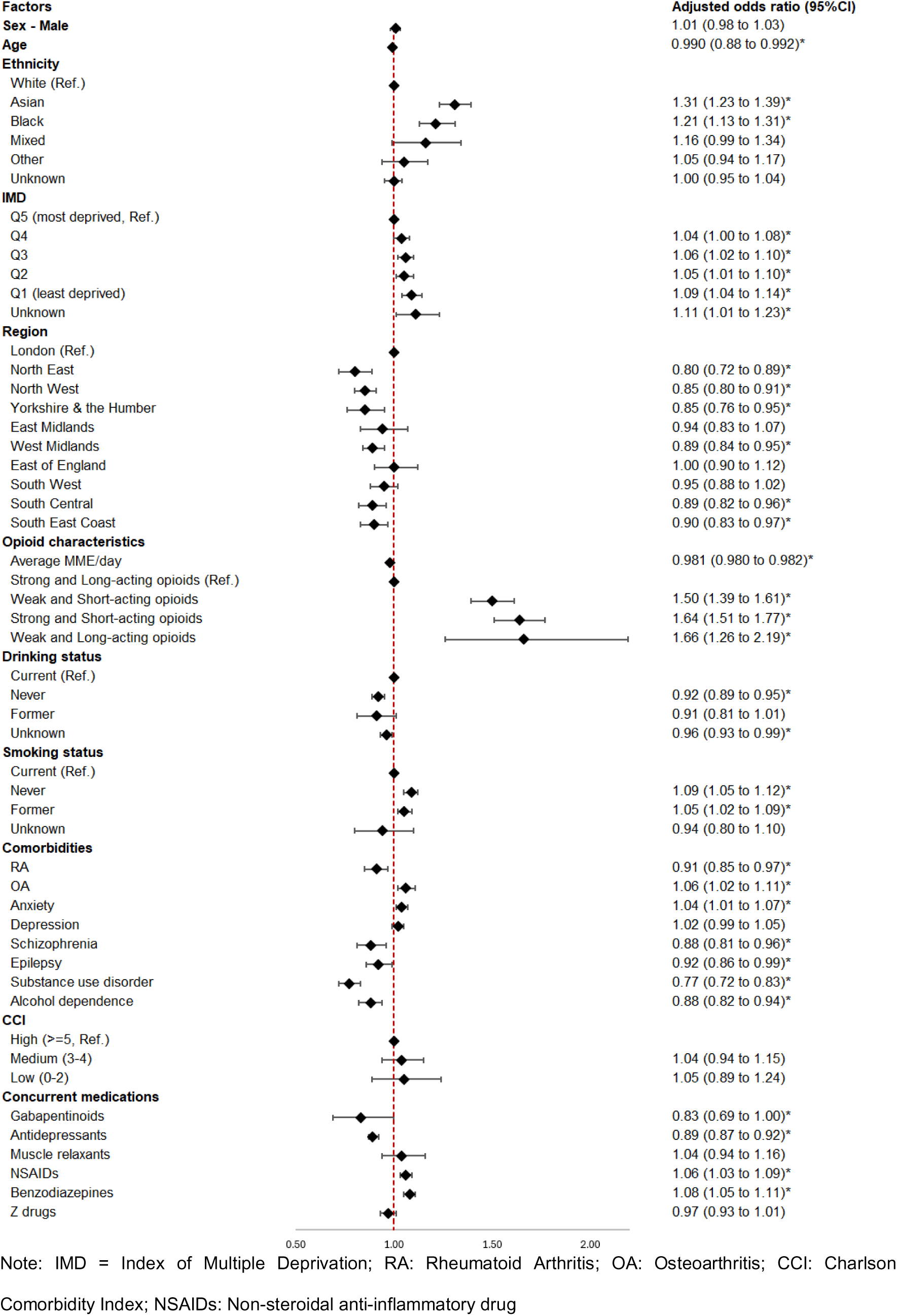
Factors associated with L-TOT discontinuation within one year after for patients with CNCP.

Geographical location also influenced L-TOT discontinuation, with the likelihood decreasing in more northern regions. For example, compared to residents in London, discontinuation was least likely in the North East (aOR 0.80, 95% CI 0.72 to 0.89), followed by the North West (aOR 0.85, 95% CI 0.80 to 0.91) and Yorkshire & the Humber (aOR 0.85, 95% CI 0.76 to 0.95). Some southern regions, such as South Central (aOR 0.89, 95% CI 0.82 to 0.96) and South East Coast (aOR 0.90, 95% CI 0.83 to 0.97), also had lower discontinuation rates but to a lesser extent.

Socioeconomic status, measured by IMD and presented in quintiles, was associated with discontinuation, with the least deprived (Q1) showing the highest likelihood (aOR 1.09, 95% CI 1.04 to 1.23). Co-existing osteoarthritis (aOR 1.06, 95% CI 1.02 to 1.11) or anxiety (aOR 1.04, 95% CI 1.01 to 1.07) were significantly associated with a higher likelihood of discontinuation. Concurrent use of non-steroidal anti-inflammatory drugs ([NSAIDs] aOR 1.06, 95% CI 1.03 to 1.09), and benzodiazepines (aOR 1.08, 95% CI 1.05 to 1.11) also significantly increased the likelihood of L-TOT discontinuation.

### Factors associated with a lower likelihood of L-TOT discontinuation

Older people were less likely to discontinue L-TOT, with approximately 1% decrease in the likelihood per one-year increase in age (aOR 0.990, 95% CI 0.988 to 0.992). Those on higher opioid doses (MME/day) were less likely to discontinue L-TOT, with an aOR of 0.981 (95% CI 0.980 to 0.982). Additional factors associated with decreased discontinuation rates included never drinking alcohol (aOR 0.92, 95% CI 0.89 to 0.95), co-existing rheumatoid arthritis (aOR 0.91, 95% CI 0.85 to 0.97), schizophrenia (aOR 0.88, 95% CI 0.81 to 0.96), epilepsy (aOR 0.92, 95% CI 0.86 to 0.99), having a history of SUDs (aOR 0.77, 95% CI 0.72 to 0.83), and alcohol dependence (aOR 0.88, 95% CI 0.82 to 0.94). L-TOT discontinuation was also less common for CNCP patients who co-used gabapentinoids (aOR 0.83, 95% CI 0.69 to 1.00) or antidepressants (aOR 0.89, 95% CI 0.87 to 0.92). No significant associations were found between L-TOT discontinuation and factors such as sex, comorbid depression, CCI scores, co-administration of Z-drugs or muscle relaxants.

### Sensitivity analysis

Results of the time varying cox regression model and competing risk model (Supplementary Table S3) were compared with the main analysis in Figure 1. Overall, the three regression models yielded consistent estimates in both direction and magnitude, with variations ranging from 2% to 40%. However, some differences in statistical significance suggested that time-varying effects and competing risks influenced specific factors, including sex, mixed ethnicity, former alcohol use, comorbid depression, CCI scores, concurrent use of muscle relaxants or Z-drugs.

## Discussion

This study identified several factors associated with the likelihood of L-TOT discontinuation among people with CNCP, which can be categorised into modifiable and non-modifiable determinants. The former primarily reflected patient lifestyle behaviours (e.g., being a non-smoker or a former smoker) and prescriber behaviours (e.g., prescribing lower daily doses or weak, short-acting opioids, and co-prescribing other pain medications such as benzodiazepines or NSAIDs). These factors are actionable for developing interventions. For example, clinicians could optimise prescribing practices that emphasise opioid tapering strategies and adjunctive pain management approaches, potentially increasing the likelihood of L-TOT discontinuation. Additionally, encouraging healthier lifestyle behaviours, i.e., supporting smoking cessation among current smokers could further enhance discontinuation efforts.

Non-modifiable factors linked with a higher likelihood of L-TOT discontinuation included sociodemographic and clinical characteristics, such as younger age, being Asian or Black, having lower deprivation scores, and comorbidities such as anxiety or osteoarthritis. These factors are less amenable to direct intervention but provide valuable information for risk stratification and personalised care. For instance, identifying patients with these characteristics could help tailor clinical strategies, ensuring those more likely to discontinue receive appropriate support, while those less likely to discontinue are offered additional resources, closer monitoring, or gradual tapering plans to facilitate safe opioid reduction. Conversely, certain non-modifiable factors, such as a history of substance use disorders, alcohol dependence, or co-existing conditions like epilepsy or schizophrenia, were associated with a reduced likelihood of L-TOT discontinuation.

### Comparison with other studies

Our findings were consistent with previous studies, ^10, 26^ that younger age was significantly associated with a higher likelihood of discontinuation, with increasing age marginally decreasing the likelihood. This may be because younger people are more likely to experience reversible causes of pain, such as post-operative pain or traumatic injuries (e.g., ligament injuries), which tend to heal over time. In contrast, older people often suffer from chronic, progressive conditions like osteoarthritis or degenerative diseases, which may require prolonged opioid use, making discontinuation more challenging. The finding that lower long-term doses of opioids was associated with higher rate of discontinuation also aligned with existing studies ^11, 15, 27^, as higher doses (defined as ≥90 MME/day in previous studies) are often associated with greater physical dependence, ^12^ making discontinuation more challenging.

Benzodiazepines have been found to be associated with L-TOT discontinuation in previous research, ^13^ consistent with our findings. We also found that concurrent use of NSAIDs was linked to a greater likelihood of discontinuation. However, there is insufficient evidence to support these medications as substitutes for opioids in CNCP management. For example, NICE (National Institute for Health and Care Excellence) ^28^ has highlighted the limited benefits of benzodiazepines and NSAIDs for chronic pain, noting that benzodiazepines may worsen psychological and physical functioning compared to placebo and carry addictive risks, and NSAIDs may lead to gastrointestinal bleeding, leading the committee to recommend against their long-term use. Gabapentinoids are often prescribed for chronic pain, even in non-neuropathic conditions, and when prescribed, are typically given to patients with severe pain, ^29^ making opioid discontinuation less likely, as confirmed by our findings. Although antidepressants such as duloxetine and amitriptyline are not licensed for CNCP, they are routinely used in UK clinical practice for treating pain. Given this, their concurrent use may be expected to promote L-TOT discontinuation. However, our findings suggested the opposite. This may have been because we considered antidepressants as a single category, which will have included antidepressants routinely used for their analgesic properties (e.g., as neuropathic painkillers), and antidepressants used solely for the indication of depression. Previous research has shown that the concurrent use of antidepressants was often associated with continued opioid use, ^30^ suggesting that patients who require antidepressants may be experiencing both depression and more severe pain. This dual burden likely leads to prolonged opioid use, making discontinuation less likely.

Previous research using CPRD Gold data among individuals with axial spondylarthritis or psoriatic arthritis identified smoking as a factor associated with increased L-TOT. ^30^ Our findings suggested that non-smoking status was associated with a higher likelihood of L-TOT discontinuation among individuals with CNCP. While this may superficially seem complementary, i.e., smoking is linked to increased L-TOT and non-smoking is associated with discontinuation, it is important to consider the differences in study populations and contexts. A possible underlying mechanism that could explain our findings is that smoking may accelerate opioid metabolism through the induction of hepatic cytochrome P450 enzymes, particularly CYP1A2 and CYP3A4. ^31, 32^ This increased metabolism could reduce the efficacy of opioids, leading to higher doses or prolonged use to achieve the desired analgesic effect, thereby making discontinuation more challenging. Conversely, non-smokers may experience more stable opioid pharmacokinetics, which could facilitate successful opioid cessation. Additionally, non-smokers may engage in healthier behaviours and be more proactive in seeking medical support, which may further contribute to their higher likelihood of discontinuing L-TOT.

According to the Faculty of Pain Medicine in the UK ^33^ as well as the US guidelines, ^34, 35^ long-acting opioids are more recommended for CNCP management. However, in our sample, most patients (∼96%) used short-acting opioids, consistent with the UK recommendations that initial treatment should begin with short-acting opioids for short-term before people reach the criteria to be diagnosed as having CNCP. Our finding that short-acting and weak opioids were associated with a greater likelihood of discontinuation aligned with current evidence. ^26, 27^ These formulations are associated with a reduced risk of developing opioid tolerance and physical dependence, ^36, 37^ allowing prescribers for regular reassessment and exploring alternative therapies sooner, therefore avoiding prolonged opioid use. This finding is important and actionable by prescribers.

Prior research ^11, 16^ has found that non-White people, particularly Asians and Blacks had higher rates of discontinuation, which aligned with our findings. These groups are often prescribed lower doses of opioids compared to their White counterparts, ^38^ thus less likely to develop long-term dependence on opioids, making discontinuation easier. Beyond ethnic disparities, geographic location and socioeconomic status also significantly influenced L-TOT discontinuation rates. Specifically, people living in less deprived areas had a higher likelihood of L-TOT discontinuation, while residing outside London linked with a lower likelihood. This may be attributed to the better access to healthcare resources, enhanced patient education, and increased awareness of the risks associated with long-term opioid use and the importance of self-management for CNCP in less deprived regions. ^39, 40^ By contrast, the most deprived regions face significant challenges, particularly in accessing specific chronic pain management services. For example, Shrujal et al. (2022) ^41^ found that pain multidisciplinary teams in these areas tend to have fewer types of professionals (≤3), which may limit the availability and quality of comprehensive pain management options. These regional inequities highlight the need for a fairer distribution of healthcare resources to improve the support for patients attempting to discontinue L-TOT.

Contradictory to earlier studies, where schizophrenia and SUDs increased the chance of discontinuation, ^10, 11^, our study found schizophrenia, epilepsy, SUDs and alcohol dependence were associated with a decrease in discontinuation rates. This discrepancy may be explained by differences in study populations and contexts. Prior studies were predominantly conducted among veterans receiving high-dose opioids within structured healthcare systems, where opioid monitoring programmes ^42^ and a proactive opioid taper decision tool ^43^ are often available, and prescribers often take greater care to prevent potential risks of serious adverse events (e.g. overdose death, respiratory depression) ^44, 45^ for this high-risk group with co-morbid mental health conditions or SUDs. In contrast, our study reflected a broader primary care population characterised by lower opioid doses and different healthcare system.

### Strengths and limitations of this study

To our knowledge, this is the first study in UK primary care to identify sociodemographic characteristics, comorbidities, lifestyle and pharmacological factors associated with successful L-TOT discontinuation among people with CNCP. The large sample size and the comprehensive data from CPRD Aurum provides robust evidence that may inform the development of future opioid reduction or discontinuation strategies.

However, several limitations need to be acknowledged. First, data from CPRD Aurum provides information on opioid prescriptions, but it may not capture opioid utilisation from other sources. For example, over-the-counter opioids like codeine and dihydrocodeine are available in the UK, which may not be reflected in the prescription records. This could potentially lead to an underrepresentation of the overall opioid utilisation in the study population. Second, the availability of prescription records does not guarantee that patients actually took the medications as prescribed. This limitation is inherent to the use of electronic health records and the reliance on prescription records. Third, our research conclusions may not be generalisable to other healthcare settings and populations that differ to the UK. Finally, while efforts have been made to account for as many confounders as possible, this observational study is subject to potential residual confounding, such as genetic, occupational, or healthcare system-related factors, that could influence the study outcomes. Particularly, data on pain severity and the discontinuation or reduction strategies implemented in different practices or regions were not available, despite their critical role in successful L-TOT discontinuation. This information is not captured in CPRD, limiting our ability to assess its impact.

### Clinical implications

This study highlights the need for tailored and personalised strategies to facilitate L-TOT discontinuation among people with CNCP. Addressing lifestyle behaviours, such as smoking cessation, may improve outcomes by reducing factors that contribute to prolonged opioid use. Additionally, careful medication selection at initiation, including choosing appropriate opioid types and considering the concurrent use of other medications, could enhance the likelihood of successful discontinuation while preventing long-term dependence. However, high-risk patients, such as those with severe pain, significant mental health comorbidities, or complex social challenges, require more intensive support beyond simple dose adjustments. For these people, a comprehensive, multidisciplinary approach integrating pain management, mental health care, and social services is essential. By incorporating these considerations, clinicians can implement more effective strategies to optimise opioid reduction and achieve L-TOT discontinuation while preventing CNCP exacerbation.

### Conclusions

This study identified several factors associated with the likelihood of L-TOT discontinuation in patients with CNCP, which can be categorised into modifiable and non-modifiable determinants. Our findings suggested that prescribers in primary care should consider a holistic approach when initiating L-TOT discontinuation, taking into account patients’ health conditions, medication profiles, and lifestyle factors. Prescribing short acting, weaker opioids at lower doses is recommended in guidance, and our study shows that this is an effective measure in increasing the likelihood of discontinuation. A fairer distribution of healthcare resources, particularly in underserved areas, could also help to improve support for patients attempting to discontinue L-TOT and reduce the burden of chronic pain management across the UK. Further research is warranted to explore the clinical and patient-reported outcomes following L-TOT discontinuation, as well as reasons for discontinuation. Such insights will be informative in developing effective pain management strategies and promoting the safe reduction of opioid use.

### Ethical approval

This study was approved by the CPRD’s Independent Scientific Advisory Committee (protocol number: 23_002909, available at https://www.cprd.com/approved-studies/predictors-discontinuation-or-reduction-long-term-opioid-therapy-and-its). CPRD also has ethical approval from the Health Research Authority to support research using anonymised patient data (research ethics committee reference 21/EM/0265). Individual patient consent was not required as all data were deidentified. This study was reported according to the Strengthening the Reporting of Observational Studies in Epidemiology (STROBE) Statement (Supplementary Table S4). ^46^

## Data availability

This study used anonymised individual-level data from the CPRD Aurum. The data cannot be shared publicly due to CPRD regulations and ethical reasons. Other researchers can use the CPRD data in a secure environment by submitting a research protocol to the CPRD Independent Scientific Advisory Committee. Details of the application process and conditions of access are provided by the CPRD at https://www.cprd.com/Data-access.

## Acknowledgments

We appreciate Professor Daren Ashcroft and the NIHR Greater Manchester Patient Safety Research Collaboration, for covering the data access costs. We also thank all data providers and GPs for making anonymised data available for research.

## Footnotes

**Contributors:** QC conceived and designed the study. EK, TA, CG and YTH supervised the conduct of the study. CM reviewed the code lists. QC conducted the data analysis and drafted the initial version of the manuscript. QC, EK, TA, CG, YTH contributed to the interpretation of the findings. All authors have reviewed the manuscript and contributed to revisions. QC is the guarantor. The corresponding author (QC) attests that all listed authors meet authorship criteria and that no others meeting the criteria have been omitted.

## Transparency

The lead author (QC) confirms that the manuscript is an honest, accurate, and transparent account of the study, with no important omissions or unexplained discrepancies from the planned study.

## Dissemination to participants and related patient/public communities

As this study used anonymised CPRD data, direct dissemination to individuals is not possible. The University of Manchester networks will further disseminate the results, as the study forms a significant part of QC’s PhD.

