## Supplementary Table S1-S4, Supplementary Figure S1-S3 for "Factors associated with long-term opioid therapy discontinuation for people with chronic non-cancer pain in UK primary care: a population-based retrospective cohort study"

**Supplementary materials**

### **Supplementary Table S1. CNCP code list**

| **medcodeid** | **Description** |
| --- | --- |
| 1634018 | Occipital headache |
| 8258016 | Repetitive strain injury |
| 10927012 | Congenital fusion of sacroiliac joint |
| 12536011 | Fracture of shaft of tibia |
| 16579010 | Meningococcal arthropathy |
| 16833013 | Ankylosing spondylitis |
| 16926014 | Fracture of scapula |
| 18370015 | Open fracture of eight OR more ribs |
| 30355011 | Post-herpetic trigeminal neuralgia |
| 33856019 | Closed fracture of seven ribs |
| 41990019 | Headache |
| 49786013 | Closed fracture of scapula |
| 53428014 | Fracture of tibia |
| 58058010 | Coccygodynia |
| 58293010 | Pyogenic arthritis of multiple sites |
| 62415013 | Multiple fractures of hand bones |
| 64723010 | Tuberculosis of spinal meninges |
| 65676015 | Congenital anomaly of spinal meninges |
| 65958012 | Closed fracture of four ribs |
| 68961012 | Thoracic spinal stenosis |
| 69073019 | Temporal headache |
| 71168014 | Peripheral neuropathy |
| 73439011 | Open fracture of five ribs |
| 75611015 | Closed fracture of one rib |
| 79758010 | Open fracture of scapula |
| 80709010 | Gouty arthritis |
| 80749019 | Fracture of vault of skull |
| 82623012 | Ophthalmoplegic migraine |
| 84797011 | Fracture of shaft of humerus |
| 85039010 | Fracture of patella |
| 86706011 | Malunion of fracture |
| 89215016 | Gonococcal spondylitis |
| 98501012 | Hemiplegic migraine |
| 99770017 | Sprain of cruciate ligament of knee |
| 107463010 | Closed fracture of calcaneus |
| 108529013 | Polymyalgia rheumatica |
| 110116013 | Fracture of humerus |
| 118420016 | Salmonella arthritis |
| 126406018 | Spinal stenosis |
| 127546017 | Epidemic cervical myalgia |
| 148612017 | Closed fracture of six ribs |
| 150580019 | Closed fracture of eight OR more ribs |
| 158460019 | Ophthalmic migraine |
| 178449010 | Ankylosis of joint |
| 178460018 | Periarthritis of wrist |
| 178862018 | Open fracture of trachea |
| 178863011 | Open fracture of clavicle |
| 216227011 | Pathological fracture due to metastatic bone disease |
| 216665012 | [Q] Fractures involving the epiphyseal plate |
| 251645017 | H/O: migraine |
| 251646016 | H/O: trigeminal neuralgia |
| 251794010 | H/O: rheumatoid arthritis |
| 252316013 | C/O - upper back ache |
| 266618015 | Packing of maxilla to correct blow-out fracture of orbit |
| 293392012 | Gouty arthropathy |
| 297456010 | Phantom limb syndrome |
| 297551015 | Chronic painful diabetic neuropathy |
| 297554011 | Polyneuropathy in amyloidosis |
| 297564019 | Polyneuropathy in porphyria |
| 297566017 | Polyneuropathy in sarcoidosis |
| 299135012 | Malleus ankylosis |
| 305093012 | Menopausal headache |
| 309459014 | Staphylococcal arthritis and polyarthritis |
| 309477016 | Sexually acquired reactive arthropathy of multiple sites |
| 309518017 | Arthropathy in Whipple's disease |
| 309555017 | Helminthiasis with arthropathy of the ankle and foot |
| 309556016 | Helminthiasis with arthropathy of multiple sites |
| 309570016 | Reactive arthropathy of hip |
| 309572012 | Reactive arthropathy of knee |
| 309574013 | Reactive arthropathy of ankle |
| 309575014 | Reactive arthropathy of subtalar joint |
| 309576010 | Reactive arthropathy of talonavicular joint |
| 309744019 | Arthropathy in Crohn's disease |
| 309749012 | Arthropathy associated with dermatological disorders |
| 309787016 | Rheumatoid arthritis of cervical spine |
| 309790010 | Rheumatoid arthritis of sternoclavicular joint |
| 309792019 | Rheumatoid arthritis of elbow |
| 309794018 | Rheumatoid arthritis of wrist |
| 309803012 | Rheumatoid arthritis of subtalar joint |
| 309816012 | Flare of rheumatoid arthritis |
| 309877012 | Erosive osteoarthrosis |
| 309878019 | Heberden's nodes with arthropathy |
| 309892013 | Localised, primary osteoarthritis of the hand |
| 309914014 | Localised, secondary osteoarthritis |
| 309933016 | Localised, secondary osteoarthritis of the ankle and foot |
| 310053015 | Traumatic arthropathy of the hand |
| 310054014 | Traumatic arthropathy of the pelvic region and thigh |
| 310062018 | Traumatic arthropathy-elbow |
| 310085017 | Allergic arthritis of the hand |
| 310086016 | Allergic arthritis of the pelvic region and thigh |
| 310088015 | Allergic arthritis of the ankle and foot |
| 310099018 | Climacteric arthritis of the hand |
| 310102018 | Climacteric arthritis of the ankle and foot |
| 310159018 | Generalised arthritis |
| 310543012 | Ankylosis of the wrist joint |
| 310544018 | Wrist joint ankylosis |
| 310551010 | Knee joint ankylosis |
| 310552015 | Ankle joint ankylosis |
| 310787017 | Palindromic rheumatism of multiple sites |
| 310818014 | Arthralgia of sternoclavicular joint |
| 310826018 | Arthralgia of wrist |
| 311112016 | Brucella spondylitis |
| 311114015 | Neuropathic spondylopathy |
| 311230016 | Idiopathic thoracic spinal stenosis |
| 311231017 | Degenerative thoracic spinal stenosis |
| 311238011 | Degenerative lumbar spinal stenosis |
| 311239015 | Iatrogenic lumbar spinal stenosis |
| 311252013 | Lumbago with sciatica |
| 311285017 | Thoracic spine ankylosis |
| 311287013 | Lumbar spine ankylosis |
| 311288015 | Atlanto-occipital instability |
| 311292010 | Thoracic spine instability |
| 311307012 | Rheumatism, excluding the back |
| 311687015 | Rheumatism and fibrositis unspecified |
| 311690014 | Muscular rheumatism |
| 312165013 | Postmenopausal osteoporosis with pathological fracture |
| 312802011 | Thoracic spinal meningocele |
| 318117015 | Closed fracture vault of skull with intracranial injury |
| 318198012 | Fracture of lower jaw, closed |
| 318200018 | Closed fracture of mandible, condylar process |
| 318214018 | Open fracture of mandible, subcondylar |
| 318215017 | Open fracture of mandible, coronoid process |
| 318217013 | Open fracture of mandible, angle of jaw |
| 318223015 | Open fracture of mandible, multiple sites |
| 318239010 | Fracture of malar and maxillary bones |
| 318251012 | Fracture of palate, open |
| 318381019 | Closed fracture axis, posterior arch |
| 318386012 | Closed fracture cervical vertebra, spinous process |
| 318388013 | Closed fracture cervical vertebra, posterior arch |
| 318389017 | Closed fracture cervical vertebra, tricolumnar |
| 318390014 | Multiple closed fractures of cervical vertebrae |
| 318423017 | Open fracture axis, transverse process |
| 318427016 | Open fracture cervical vertebra, wedge |
| 318430011 | Open fracture cervical vertebra, transverse process |
| 318433013 | Multiple open fractures of cervical vertebrae |
| 318438016 | Closed fracture thoracic vertebra, spondylolysis |
| 318440014 | Closed fracture thoracic vertebra, transverse process |
| 318441013 | Closed fracture thoracic vertebra, posterior arch |
| 318442018 | Closed fracture thoracic vertebra, tricolumnar |
| 318446015 | Open fracture thoracic vertebra |
| 318450010 | Open fracture thoracic vertebra, spinous process |
| 318460018 | Closed fracture lumbar vertebra, posterior arch |
| 318461019 | Closed fracture lumbar vertebra, tricolumnar |
| 318464010 | Open fracture lumbar vertebra, wedge |
| 318466012 | Open fracture lumbar vertebra, spinous process |
| 318477018 | Open compression fracture sacrum |
| 318482013 | Fracture of first cervical vertebra |
| 318483015 | Fracture of second cervical vertebra |
| 318484014 | Multiple fractures of cervical spine |
| 318485010 | Fracture of lumbar spine and pelvis |
| 318500017 | Closed fracture of cervical spine with cord lesion |
| 318514017 | Open fracture of cervical spine with spinal cord lesion |
| 318518019 | Open spinal fracture with central cervical cord lesion, C1-4 |
| 318524013 | Open spinal fracture with central cervical cord lesion, C5-7 |
| 318542015 | Open fracture of thoracic spine with spinal cord lesion |
| 318556018 | Closed fracture of lumbar spine with spinal cord lesion |
| 318558017 | Closed spinal fracture with complete lumbar cord lesion |
| 318559013 | Closed spinal fracture with anterior lumbar cord lesion |
| 318560015 | Closed spinal fracture with central lumbar cord lesion |
| 318561016 | Closed spinal fracture with posterior lumbar cord lesion |
| 318569019 | Open spinal fracture with cauda equina lesion |
| 318573016 | Closed fracture of sacrum with complete cauda equina lesion |
| 318583017 | Open fracture of sacrum with complete cauda equina lesion |
| 318595017 | Open fracture of coccyx with complete cauda equina lesion |
| 318633016 | Open fracture larynx and trachea |
| 318635011 | Open fracture of hyoid bone |
| 318636012 | Open fracture of thyroid cartilage |
| 318641016 | Cough fracture of ribs |
| 318654011 | Fracture or disruption of pelvis |
| 318656013 | Closed fracture acetabulum, posterior lip alone |
| 318658014 | Closed fracture acetabulum, posterior column |
| 318659018 | Closed fracture acetabulum, floor |
| 318660011 | Closed fracture acetabulum, double column transverse |
| 318691017 | Closed fracture pelvis, ischial tuberosity |
| 318695014 | Closed vertical fracture of ilium |
| 318707016 | Open vertical fracture of ilium |
| 318746019 | Open fracture scapula, spine |
| 318755016 | Closed fracture of humerus, upper epiphysis |
| 318756015 | Closed fracture proximal humerus, three part |
| 318767019 | Open fracture proximal humerus, greater tuberosity |
| 318768012 | Open fracture proximal humerus, head |
| 318769016 | Open fracture of humerus, upper epiphysis |
| 318770015 | Open fracture proximal humerus, three part |
| 318787018 | Closed fracture of distal humerus, trochlea |
| 318788011 | Closed fracture distal humerus, lateral epicondyle |
| 318790012 | Closed fracture distal humerus, bicondylar (T-Y fracture) |
| 318791011 | Closed fracture of distal humerus, multiple |
| 318804015 | Open fracture distal humerus, bicondylar (T-Y fracture) |
| 318815013 | Closed fracture olecranon, extra-articular |
| 318822017 | Open fracture olecranon, extra-articular |
| 318824016 | Open fracture proximal ulna, comminuted |
| 318825015 | Open fracture proximal radius, comminuted |
| 318845011 | Closed fracture of ulna, lower epiphysis |
| 318857019 | Closed fracture radial styloid |
| 318875018 | Open fracture of ulna, styloid process |
| 318876017 | Open fracture of ulna, lower epiphysis |
| 318887013 | Open dorsal Barton's fracture |
| 318889011 | Open fracture distal radius, intra-articular, die-punch |
| 318917016 | Closed fracture hamate, hook |
| 318919018 | Closed fracture scaphoid, waist, transverse |
| 318923014 | Closed fracture carpal bones, multiple |
| 318927010 | Open fracture hamate, hook |
| 318929013 | Open fracture scaphoid, waist, transverse |
| 318936014 | Fracture at wrist and hand level |
| 318941018 | Fracture of metacarpal bone |
| 318959015 | Closed fracture thumb metacarpal head |
| 318972019 | Open fracture of thumb metacarpal |
| 318979011 | Closed fracture sesamoid bone of hand |
| 318980014 | Open fracture sesamoid bone of hand |
| 318989010 | Closed fracture thumb proximal phalanx |
| 318991019 | Closed fracture thumb proximal phalanx, shaft |
| 318998013 | Closed fracture thumb distal phalanx, mallet |
| 318999017 | Closed fracture finger proximal phalanx |
| 319002011 | Closed fracture finger proximal phalanx, neck |
| 319004012 | Closed fracture finger proximal phalanx, multiple |
| 319009019 | Closed fracture finger middle phalanx, head |
| 319012016 | Closed fracture finger distal phalanx, shaft |
| 319013014 | Closed fracture finger distal phalanx, tuft |
| 319015019 | Closed fracture finger distal phalanx, multiple |
| 319016018 | Closed fractures of phalanx or phalanges, multiple sites |
| 319026013 | Open fracture thumb proximal phalanx, head |
| 319029018 | Open fracture thumb distal phalanx, shaft |
| 319032015 | Open fracture finger proximal phalanx |
| 319034019 | Open fracture finger proximal phalanx, shaft |
| 319035018 | Open fracture finger proximal phalanx, neck |
| 319036017 | Open fracture finger proximal phalanx, head |
| 319037014 | Open fracture finger proximal phalanx, multiple |
| 319039012 | Open fracture finger middle phalanx, base |
| 319048019 | Open fracture finger distal phalanx, multiple |
| 319057013 | Ill-defined fractures of upper limb |
| 319058015 | Ill-defined fracture of arm |
| 319059011 | Closed ill-defined fractures of upper limb |
| 319062014 | Open ill-defined fractures of upper limb |
| 319071017 | Multiple fractures of clavicle, scapula and humerus |
| 319074013 | Multiple fractures of forearm |
| 319094016 | Closed fracture proximal femur, subcapital, Garden grade III |
| 319095015 | Closed fracture proximal femur, subcapital, Garden grade IV |
| 319108013 | Open fracture proximal femur,subcapital, Garden grade unspec |
| 319111014 | Open fracture proximal femur,subcapital, Garden grade III |
| 319118015 | Closed fracture of proximal femur, pertrochanteric |
| 319133011 | Open fracture proximal femur, intertrochanteric, two part |
| 319163019 | Closed fracture distal femur, lateral condyle |
| 319172010 | Open fracture of femur, lower epiphysis |
| 319173017 | Open fracture distal femur, medial condyle |
| 319174011 | Open fracture distal femur, lateral condyle |
| 319175012 | Open fracture distal femur, bicondylar (T-Y fracture) |
| 319183018 | Closed fracture patella, distal pole |
| 319188010 | Closed fracture patella, comminuted (stellate) |
| 319190011 | Open fracture patella, proximal pole |
| 319191010 | Open fracture patella, distal pole |
| 319192015 | Open fracture patella, vertical |
| 319200010 | Closed fracture proximal tibia, medial condyle (plateau) |
| 319202019 | Closed fracture proximal tibia, bicondylar |
| 319205017 | Closed fracture fibula, neck |
| 319214010 | Open fracture proximal tibia, bicondylar |
| 319227014 | Closed fracture distal tibia |
| 319228016 | Closed fracture distal tibia, extra-articular |
| 319229012 | Closed fracture distal tibia, intra-articular |
| 319232010 | Open fracture distal tibia, intra-articular |
| 319262016 | Closed fracture ankle, bimalleolar, high fibular fracture |
| 319264015 | Open fracture ankle, bimalleolar, high fibular fracture |
| 319265019 | Closed fracture ankle, trimalleolar, low fibular fracture |
| 319266018 | Closed fracture ankle, trimalleolar, high fibular fracture |
| 319273011 | Fracture of ankle, NOS |
| 319297019 | Closed fracture metatarsal shaft |
| 319299016 | Closed fracture metatarsal head |
| 319300012 | Closed fracture metatarsal, multiple |
| 319301011 | Closed tarsal fractures, multiple |
| 319305019 | Open fracture talus, head |
| 319306018 | Open fracture talus, neck |
| 319319019 | Closed fracture proximal phalanx, toe |
| 319320013 | Closed fracture middle phalanx, toe |
| 319321012 | Closed fracture distal phalanx, toe |
| 319322017 | Closed fracture multiple phalanges, toe |
| 319327011 | Open fracture multiple phalanges, toe |
| 319340018 | Multiple fractures of femur |
| 319342014 | Multiple fractures of foot |
| 319764019 | Closed spinal dislocation with anterior thoracic cord lesion |
| 319765018 | Closed spinal dislocation with central thoracic cord lesion |
| 319770013 | Closed spinal dislocation with central lumbar cord lesion |
| 319771012 | Closed spinal dislocation with posterior lumbar cord lesion |
| 319836018 | Closed spinal subluxation with complete cervical cord lesion |
| 319838017 | Closed spinal subluxation with central cervical cord lesion |
| 319865012 | Closed spinal subluxation with complete thoracic cord lesion |
| 319871018 | Closed spinal subluxation with anterior lumbar cord lesion |
| 319872013 | Closed spinal subluxation with central lumbar cord lesion |
| 319873015 | Closed spinal subluxation with posterior lumbar cord lesion |
| 319970018 | Open fracture dislocation wrist |
| 320119017 | Sprain, shoulder joint, posterior |
| 320122015 | Sprain, triceps tendon |
| 320137016 | Sprain, elbow joint, medial collateral ligament |
| 320139018 | Radiohumeral sprain |
| 320140016 | Ulnohumeral sprain |
| 320145014 | Sprain of wrist and hand |
| 320148011 | Carpal joint sprain |
| 320150015 | Distal radioulnar joint sprain |
| 320167010 | Sprain volar intercarpal ligament or V ligament |
| 320178019 | Metacarpophalangeal sprain |
| 320180013 | Midcarpal joint sprain |
| 320212019 | Sprain wrist flexors |
| 320218015 | Sprain, flexor digitorum profundus tendon |
| 320219011 | Sprain, extensor digitorum tendon |
| 320258018 | Sprain of ankle and foot |
| 320260016 | Sprain, ankle joint, medial |
| 320262012 | Sprain, ankle joint, lateral |
| 320282011 | Sprain pelvic ligament |
| 320284012 | Sprain, lumbosacral ligament |
| 320287017 | Sprain, sacrospinous ligament |
| 320289019 | Sprain, iliolumbar ligament |
| 320295018 | Neck sprain |
| 320472012 | Open division sacroiliac ligament |
| 320558017 | Chondrocostal joint sprain |
| 320563018 | Sternoclavicular sprain |
| 320564012 | Chondrosternal sprain |
| 320565013 | Xiphoid cartilage sprain |
| 320567017 | Pelvis sprain or complete tear |
| 323827010 | Open injury sciatic nerve |
| 325243011 | Fractures involving multiple body regions |
| 325247012 | Fractures involving thorax with lower back and pelvis |
| 325248019 | Fractures involving multiple regions of one upper limb |
| 325252019 | Fractures involving multiple regions of both lower limbs |
| 325265011 | Dislocations, sprains and strains involving head with neck |
| 345367011 | Chronic post-traumatic headache |
| 345580013 | Intercostal neuropathy |
| 346003014 | Intracranial destruction of trigeminal ganglion |
| 354567013 | Beta-2 microglobulin arthropathy |
| 356490015 | Amyloid arthropathy |
| 356491016 | Arthropathy in amyloidosis |
| 359267014 | Wrist pyogenic arthritis |
| 359310019 | Juvenile arthritis in psoriasis |
| 359321011 | Distal interphalangeal psoriatic arthropathy |
| 359433019 | Osteoarthritis of lumbar spine |
| 359513016 | Giant cell arteritis with polymyalgia rheumatica |
| 359719015 | Viral myalgia |
| 360154013 | Postmeningococcal arthritis |
| 364017010 | Closed fracture of tibia and fibula, proximal |
| 369417015 | Neuropathic pain |
| 391281018 | Fracture of upper jaw, closed |
| 391312019 | Fracture of nasal bones |
| 391345019 | Fracture of lower end of humerus |
| 391354016 | Fracture of lower end of radius |
| 391359014 | Fracture of upper end of ulna |
| 391360016 | Fracture of shaft of ulna |
| 391366010 | Fracture of lower end of both ulna and radius |
| 391392015 | Subtrochanteric fracture |
| 391394019 | Open fracture base of neck of femur |
| 391396017 | Fracture of lower end of femur |
| 391413011 | Fracture of talus |
| 391423019 | March fracture |
| 397840012 | H/O: musculoskeletal disease |
| 397993014 | Frontal headache |
| 400175010 | Generalised osteoarthritis of the hand |
| 400224017 | Joint ankylosis of the pelvic region and thigh |
| 400226015 | Joint ankylosis of the ankle and foot |
| 400246013 | Arthralgia of shoulder |
| 400249018 | Arthralgia of the pelvic region and thigh |
| 400338014 | Pathological fracture |
| 402827014 | Fracture of malar or maxillary bones, open |
| 402837016 | Closed fracture of fourth cervical vertebra |
| 402838014 | Closed fracture of fifth cervical vertebra |
| 402840016 | Closed fracture of seventh cervical vertebra |
| 402841017 | Open fracture of cervical spine |
| 402845014 | Open fracture of fourth cervical vertebra |
| 402846010 | Open fracture of fifth cervical vertebra |
| 402859019 | Closed Colles' fracture |
| 402877012 | Fracture of one or more tarsal and metatarsal bones |
| 402911018 | Sprain of elbow and forearm |
| 405143010 | Pathological fracture of thoracic vertebra |
| 405550017 | Congenital instability of hip joint |
| 410903011 | K wiring of fracture |
| 411067016 | FH: Arthritis |
| 411301012 | Shoulder strain |
| 411306019 | Open fracture of femur, subcapital |
| 416143014 | Lumbago |
| 419567010 | Fracture of lateral malleolus |
| 423262010 | Open wound of sacroiliac region |
| 443804014 | Elbow fracture - closed |
| 451079015 | Open fracture of distal fibula |
| 451463012 | Pathological fracture of cervical vertebra |
| 453181013 | Lumbosacral strain |
| 455444016 | Closed multiple fractures of thoracic spine |
| 455445015 | Open multiple fracture of thoracic spine |
| 455451013 | Closed fracture of great toe |
| 457118013 | Localised, primary osteoarthritis of the wrist |
| 457121010 | Localised, primary osteoarthritis of toe |
| 479288015 | Closed fracture of proximal radius and ulna |
| 481611016 | Jaw sprain |
| 481614012 | Temporomandibular sprain |
| 481879010 | Hip joint ankylosis |
| 484889013 | Ankylosing vertebral hyperostosis |
| 489598015 | Lumbosacral instability |
| 493899010 | Ankle sprain |
| 496626018 | Arthropathy due to hypersensitivity reaction |
| 500216016 | Open fracture of proximal radius and ulna |
| 502155015 | Trigeminal (5th) nerve injury |
| 503422014 | Multiple fractures of fingers |
| 507904017 | Open fracture of radius and ulna, lower end |
| 1219617015 | Arthropathy due to fungal infection |
| 1219674017 | Open fracture radial neck |
| 1219675016 | Open fracture shaft of fibula |
| 1220722015 | Fracture of neck |
| 1220724019 | Fracture of lumbar vertebra |
| 1221098013 | Open fracture larynx |
| 1221230012 | Fracture of palate, closed |
| 1223089018 | Arm fracture |
| 1223146015 | Open fracture of humerus, shaft |
| 1223154018 | Open fracture of the proximal radius |
| 1223219019 | Closed fracture intermediate cuneiform |
| 1224865018 | Hereditary motor and sensory neuropathy type IV |
| 1225039011 | Closed fracture shaft of femur |
| 1225141010 | Open fracture ankle, bimalleolar |
| 1225880017 | Closed fracture fibula, head |
| 1227930011 | Closed fracture of the radial shaft |
| 1228153010 | Open orbital blow-out fracture |
| 1228726016 | Closed fracture metatarsal |
| 1229658014 | Open fracture nose |
| 1229659018 | Open fracture nasal bone |
| 1229825015 | Pyogenic arthritis of the forearm |
| 1229927010 | Open fracture of the ulnar shaft |
| 1230293011 | Open fracture cuneiforms |
| 1230529018 | Closed fracture zygoma |
| 1230612011 | Closed fracture clavicle, medial end |
| 1230691015 | Arthralgia of hip |
| 1230706011 | Closed orbital blow-out fracture |
| 1231287012 | Open fracture medial cuneiform |
| 1231810014 | Collar bone fracture |
| 1231853014 | Closed fracture distal humerus, supracondylar |
| 1231985013 | Closed fracture proximal fibula |
| 1232117011 | Closed fracture rib |
| 1232174014 | Pyogenic arthritis of the hand |
| 1232781014 | Open fracture shaft of femur |
| 1232881017 | Open fracture trapezium |
| 1232935018 | Closed fracture medial cuneiform |
| 1233343014 | Open fracture clavicle, shaft |
| 1233578010 | Open fracture navicular |
| 1234036019 | Closed fracture finger distal phalanx |
| 1234136017 | Open fracture acetabulum |
| 1234168013 | Closed fracture shaft of fibula |
| 1234424019 | Open fracture distal femur, supracondylar |
| 1234612017 | Closed fracture nose |
| 1235211014 | Open fracture sternum |
| 1235651015 | Familial amyloid polyneuropathy type III |
| 1485119015 | H/O: fragility fracture |
| 1485120014 | H/O: hip fracture |
| 1485121013 | H/O: vertebral fracture |
| 1485122018 | H/O: fracture |
| 1485123011 | FH: Maternal hip fracture |
| 1490703015 | Closed fracture radius and ulna, distal |
| 1494757018 | Thyroid cartilage sprain |
| 1494827019 | Open fracture of mandible, symphysis of body |
| 1494890011 | Sprain of hip and thigh |
| 1494891010 | Thigh sprain |
| 1494944016 | Closed fracture radius and ulna, proximal |
| 1777664015 | Hereditary motor and sensory neuropathy |
| 1777779015 | Amyloid polyneuropathy type I |
| 1785082014 | Rheumatic pain |
| 1786078018 | Tension headache |
| 1786163010 | Hereditary motor and sensory neuropathy type II |
| 2159844013 | Rheumatoid arthritis particle agglutination test |
| 2477015019 | Andrade type amyloid polyneuropathy |
| 2551528018 | Closed fracture pelvis, ischium |
| 2645979015 | Open fracture of base of fifth metatarsal |
| 2838283014 | Paroxysmal hemicrania |
| 110111000006117 | Test request : Rheumatoid Factor |
| 140101000000116 | H/O: non-vertebral fracture |
| 162221000006115 | Rheumatoid arthritis screening test |
| 230611000006118 | Phantom limb syndrome with pain |
| 299791000000114 | Open fracture of femur, upper epiphysis |
| 303071000000118 | History of irritable bowel syndrome |
| 622771000000113 | Pars interarticularis stress fracture |
| 878961000006118 | Tuberculosis - meninges/CNS |
| 883161000006115 | Drug/toxic polyneuropathy |
| 886361000006114 | Irritable bowel - IBS |
| 889881000006119 | Osteoarthritis - elbow joint |
| 889901000006117 | Osteoarthritis - hand joint |
| 890371000006116 | Ankylosis - multiple joint |
| 890421000006111 | Ankylosis - hand joint |
| 890431000006114 | Ankylosis - hip joint |
| 890451000006119 | Ankylosis - ankle/foot |
| 890981000006117 | Spinal stenosis excl. cervical |
| 891861000006112 | Neuralgia/neuritis - NOS |
| 892321000006118 | Hypertroph. pulm. osteoarthrop |
| 892401000006119 | Path.fracture - NOS |
| 892481000006111 | Path.fracture - multiple |
| 895271000006115 | #Cervical spine-no cord lesion |
| 895281000006117 | #Thoracic spine-no cord lesion |
| 895351000006119 | #Sacrum/coccyx + cord lesion |
| 895561000006112 | Colles fracture |
| 895861000006114 | Fracture Left Ankle |
| 896171000006115 | Dislocations/sprains/strains |
| 896231000006112 | Sprain - wrist |
| 896241000006119 | Sprained finger/thumb |
| 896261000006115 | Sprained thigh - upper leg |
| 896291000006111 | Sprained knee |
| 896301000006112 | Sprain - lateral knee ligament |
| 896321000006119 | Sprain -cruciate knee ligament |
| 896341000006114 | Sprained ankle |
| 896441000006118 | Sacral/coccyx sprain |
| 896481000006112 | Sprained ribs |
| 896491000006110 | Sprained sternum |
| 990251000006114 | Fracture malunion - shoulder |
| 990261000006111 | Fracture malunion - hand |
| 991391000006118 | Sprain - hand NOS |
| 991411000006118 | Sprain - ankle NOS |
| 991421000006114 | Sprain - foot NOS |
| 2214301000000116 | Chronic regional pain syndrome |
| 2254241000000117 | On musculoskeletal care pathway |
| 2360741000000115 | Axial spondyloarthritis |
| 2415191000000113 | Diabetic peripheral neuropathic pain |
| 309704014 | Crystal arthropathy NOS, of the pelvic region and thigh |
| 317148013 | [D]Nervous or musculoskeletal symptoms NOS |
| 41386013 | Fibromyalgia |
| 123542016 | Pauciarticular juvenile rheumatoid arthritis |
| 297355013 | Other forms of migraine NOS |
| 297428017 | Trigeminal neuralgia NOS |
| 297430015 | Trigeminal nerve disorder NOS |
| 297493016 | Other upper limb mononeuritis |
| 297511018 | Hereditary and idiopathic peripheral neuropathy |
| 297530018 | Other idiopathic peripheral neuropathy NOS |
| 299338017 | [X]Other migraine |
| 299361015 | [X]Other specified mononeuropathies |
| 299362010 | [X]Other mononeuropathies in diseases classified elsewhere |
| 299364011 | [X]Other hereditary and idiopathic neuropathies |
| 299366013 | [X]Other specified polyneuropathies |
| 308730016 | Psoriatic arthropathy NOS |
| 309441011 | Pyogenic arthritis of unspecified site |
| 309485013 | Arthropathy in Behcet's syndrome of the forearm |
| 309488010 | Arthropathy in Behcet's syndrome of the lower leg |
| 309492015 | Arthropathy in Behcet's syndrome NOS |
| 309548019 | Helminthiasis with arthropathy of unspecified site |
| 309554018 | Helminthiasis with arthropathy of the lower leg |
| 309557013 | Helminthiasis with arthropathy of other specified site |
| 309600012 | Infective arthritis NOS, of the pelvic region and thigh |
| 309601011 | Infective arthritis NOS, of the lower leg |
| 309605019 | Infective arthritis NOS, of acromioclavicular joint |
| 309606018 | Infective arthritis NOS, of elbow |
| 309614012 | Infective arthritis NOS, of hip |
| 309625015 | Infective arthritis NOS, of multiple sites |
| 309682019 | Gouty arthritis of other specified site |
| 309687013 | Other crystal arthropathies of the shoulder |
| 309689011 | Other crystal arthropathies of the forearm |
| 309690019 | Other crystal arthropathies of the hand |
| 309699018 | Crystal arthropathy NOS, site unspecified |
| 309760015 | Other general diseases with associated arthropathy |
| 309761016 | Arthritis associated with other disease, shoulder |
| 309766014 | Arthritis associated with other disease, wrist |
| 309778013 | Arthritis associated with other disease, other tarsal joint |
| 309783017 | Arthropathy associated with disorders EC NOS |
| 309911018 | Localised, primary osteoarthritis NOS |
| 309977011 | Oligoarticular osteoarthritis, unspecified, of hand |
| 310010015 | Osteoarthritis NOS, other specified site |
| 310014012 | Osteoarthritis NOS, of elbow |
| 310026019 | Osteoarthritis NOS, of ankle |
| 310029014 | Osteoarthritis NOS, of other tarsal joint |
| 310049017 | Traumatic arthropathy of unspecified site |
| 310052013 | Traumatic arthropathy of the forearm |
| 310084018 | Allergic arthritis of the forearm |
| 310087013 | Allergic arthritis of the lower leg |
| 310089011 | Allergic arthritis of other specified site |
| 310139017 | Transient arthropathy of other tarsal joint |
| 310152010 | Unspecified polyarthropathy of the hand |
| 310154011 | Unspecified polyarthropathy of the lower leg |
| 310157016 | Unspecified polyarthropathy of multiple sites |
| 310183014 | Other specified arthropathy of the shoulder region |
| 310184015 | Other specified arthropathy of the upper arm |
| 310187010 | Other specified arthropathy of the pelvic region and thigh |
| 310188017 | Other specified arthropathy of the lower leg |
| 310189013 | Other specified arthropathy of the ankle and foot |
| 310537010 | Joint ankylosis of other specified site |
| 310538017 | Ankylosis of other joint of the shoulder girdle |
| 310778014 | Palindromic rheumatism of unspecified site |
| 310780015 | Palindromic rheumatism of the upper arm |
| 310792015 | Arthralgia of unspecified site |
| 310815012 | Arthralgia of other specified site |
| 310847019 | Arthralgia of other tarsal joint |
| 311036019 | Arthropathies NOS |
| 311197011 | Cervical spinal stenosis secondary to other disease |
| 311229014 | Spinal stenosis of unspecified region |
| 311236010 | Thoracic spinal stenosis secondary to other disease |
| 311267012 | Sacral ankylosis NOS |
| 311496011 | Fibrosing alveolitis associated with rheumatoid arthritis |
| 311767011 | Other specified nonarticular rheumatism |
| 312518013 | [X]Other seropositive rheumatoid arthritis |
| 312520011 | [X]Other specified rheumatoid arthritis |
| 312534011 | [X]Seropositive rheumatoid arthritis, unspecified |
| 312804012 | Spinal meningocele NOS |
| 312946016 | Other specified spinal cord anomalies |
| 318199016 | Closed fracture mandible (site unspecified) |
| 318209017 | Fracture of mandible, closed, NOS |
| 318216016 | Open fracture of mandible, ramus, unspecified |
| 318222013 | Open fracture of mandible, body, other and unspecified |
| 318250013 | Fracture of orbit NOS, open |
| 318252017 | Fracture of other facial bones,open, NOS |
| 318256019 | Other and unqualified skull fractures |
| 318353013 | Closed fracture of unspecified cervical vertebra |
| 318391013 | Closed fracture of cervical spine not otherwise specified |
| 318443011 | Other specified closed fracture thoracic vertebra |
| 318445016 | Closed fracture thoracic vertebra not otherwise specified |
| 318582010 | Open fracture of sacrum with unspecified spinal cord lesion |
| 318584011 | Open fracture of sacrum with other cauda equina injury |
| 318590010 | Closed fracture of coccyx with other cauda equina injury |
| 318592019 | Closed fracture of coccyx with spinal cord lesion NOS |
| 318594018 | Open fracture of coccyx with unspecified spinal cord lesion |
| 318613015 | Closed fracture of rib(s) NOS |
| 318661010 | Closed fracture acetabulum, double column unspecified |
| 318662015 | Other specified closed fracture acetabulum |
| 318672017 | Open fracture acetabulum NOS |
| 318688017 | Closed fracture of ilium, unspecified |
| 318698011 | Other or multiple open fracture of pelvis |
| 318699015 | Open fracture of ilium, unspecified |
| 318719012 | Open fracture of pelvis NOS |
| 318729017 | Fracture of neck and trunk NOS |
| 318731014 | Closed fracture of clavicle NOS |
| 318732019 | Open fracture of clavicle, unspecified part |
| 318733012 | Open fracture of clavicle NOS |
| 318742017 | Open fracture of scapula, unspecified part |
| 318748018 | Open fracture of scapula NOS |
| 318749014 | Fracture of scapula NOS |
| 318772011 | Open fracture of proximal humerus not otherwise specified |
| 318774012 | Closed fracture of humerus NOS |
| 318775013 | Closed fracture of humerus, shaft or unspecified part NOS |
| 318778010 | Open fracture of humerus NOS |
| 318783019 | Closed fracture of elbow, unspecified part |
| 318786010 | Closed fracture of distal humerus, condyle(s) unspecified |
| 318792016 | Closed fracture of distal humerus, not otherwise specified |
| 318810015 | Fracture of humerus NOS |
| 318829014 | Closed fracture of radius, shaft, unspecified |
| 318862018 | Closed fracture distal radius, intra-articular, other type |
| 318912010 | Open fracture of radius (alone), unspecified |
| 318924015 | Closed fracture of other carpal bone |
| 318940017 | Fracture of carpal bone NOS |
| 318961012 | Closed fractures of multiple sites of unspecified metacarpus |
| 318977013 | Open fractures of multiple sites of unspecified metacarpus |
| 318978015 | Open fracture of metacarpal bone(s) NOS |
| 319017010 | Closed fracture of one or more phalanges of hand NOS |
| 319154016 | Open fracture of femur, unspecified part |
| 319155015 | Open fracture of femur, shaft or unspecified part, NOS |
| 319159014 | Closed fracture of distal femur, unspecified |
| 319243019 | Closed fracture of tibia and fibula, unspecified part |
| 319249015 | Open fracture of tibia and fibula, unspecified part, NOS |
| 319304015 | Open fracture of tarsal bone, unspecified |
| 319343016 | Other, multiple and ill-defined fractures of lower limb NOS |
| 320142012 | Other forearm sprain |
| 320144013 | Forearm sprain NOS |
| 320169013 | Wrist sprain NOS |
| 320181012 | Hand sprain NOS |
| 320235016 | Other thigh sprain |
| 320236015 | Hip sprain NOS |
| 320257011 | Leg sprain NOS |
| 320259014 | Ankle sprain, unspecified |
| 320548016 | Jaw sprain NOS |
| 320562011 | Sternum sprain unspecified |
| 320568010 | Sprain of pelvis, unspecified |
| 320579017 | Other and ill-defined sprains and strains NOS |
| 321974013 | Late effect of other fracture of leg |
| 321975014 | Late effect of multiple and unspecified fracture of bones |
| 323940016 | Other face and neck injuries NOS |
| 325294016 | [X]Fractures of other skull and facial bones |
| 325295015 | [X]Fracture of skull and facial bones, part unspecified |
| 325375012 | [X]Other multiple injuries of abdomen, lower back and pelvis |
| 325450012 | [X]Fractures of other parts of lower leg |
| 325508014 | [X]Fracture of unspecified body region |
| 329379013 | Fracture, cause unspecified |
| 359168010 | Ankle arthritis NOS |
| 378021011 | Congenital deformity of musculoskeletal system NEC |
| 391330014 | Closed fracture of axis without spinal cord lesion |
| 391382011 | Upper leg fracture NOS |
| 400187015 | Osteoarthritis NOS, of the lower leg |
| 400192018 | Unspecified polyarthropathy or polyarthritis NOS |
| 400248014 | Arthralgia of the forearm |
| 400941013 | Congenital musculoskeletal anomalies NOS |
| 402828016 | Fracture of facial bone NOS |
| 402829012 | Skull fracture NOS |
| 402879010 | Fracture of unspecified bones |
| 402918012 | Neck sprain, unspecified |
| 402920010 | Sprains and strains NOS |
| 402974016 | Late effect of sprain without mention of tendon injury |
| 450558019 | Polyneuropathy unspecified |
| 455453011 | Closed fracture of lower limb, level unspecified |
| 1209790019 | Arthritis of spine |
| 2474635010 | Rheumatology management plan given |
| 2476044012 | Osteoarthritis of spine NOS |
| 28251000006113 | Other infect/parasit dis with arthropathy of the forearm |
| 28261000006110 | Other infect/parasit dis with arthropathy of the hand |
| 28271000006115 | Other infect/parasit dis with arthropathy of the lower leg |
| 28281000006117 | Other infect/parasit dis with arthropathy of the upper arm |
| 29251000006116 | Other infect/parasit dis with arthropathy of ankle and foot |
| 29481000006111 | Other fracture-dislocation or subluxation |
| 31041000006113 | Osteoarthritis NOS, of distal radio-ulnar joint |
| 92851000006118 | Traumatic arthropathy of tibio-fibular joint |
| 100941000006118 | Thumb fracture excluding base |
| 129911000006119 | Sponanteous fracture |
| 130611000006116 | Sprain finger |
| 130901000006112 | Sprain radio-lunate ligament |
| 130941000006114 | Sprain scapho-trapezium ligament |
| 130951000006111 | Sprain short intrinsic ligament non-specific |
| 131051000006117 | Sprain thumb, interphalangeal joint, non specific |
| 131111000006119 | Sprain ulno-lunate ligament |
| 131231000006119 | Sprain, coraco-clavicular ligament |
| 135161000006118 | Spinal meningeal adhesions |
| 164801000006119 | Rotational mal-union of fracture |
| 168291000006116 | Rheumat.dis.treatment changed |
| 168661000006110 | Rheumatism - gonococcal |
| 168751000006115 | Rheumatoid arthritis and other inflammatory polyarthropathy |
| 217691000006119 | Postdysenteric reactive arthropathy of the ankle and foot |
| 218101000006119 | Posterior fossa fracture |
| 219611000000112 | Osteoarthritis cervical spine |
| 221481000000114 | Crystal arthritis |
| 222341000000116 | Open fracture distal femur |
| 222381000000112 | Os calcis fracture |
| 222451000000113 | Headache - post traumatic |
| 255571000006116 | Open fracture-subluxation peri-lunate trans-scaphoid |
| 255611000006114 | Open fracture-subluxation sterno-clavicular joint, anterior |
| 255631000006115 | Open fracture-subluxation superior radio-ulnar joint |
| 255661000006112 | Open fracture-subluxation, distal interphalangeal joint |
| 255771000006110 | Open fracture-subluxation, patello-femoral joint |
| 255801000006112 | Open fracture-subluxation, tarsometatarsal joint |
| 255971000006112 | Open fracture-dislocation other carpal |
| 256071000006115 | Open fracture-dislocation, distal interphalangeal joint |
| 256081000006117 | Open fracture-dislocation, distal radio-ulnar joint |
| 256181000006116 | Open fracture-dislocation, patello-femoral joint |
| 256211000006117 | Open fracture-dislocation, tarsometatarsal joint |
| 256231000006111 | Open fracture-subluxation digit |
| 256241000006118 | Open fracture-subluxation digit, unspecified |
| 256261000006119 | Open fracture-subluxation elbow joint |
| 256301000006111 | Open fracture-subluxation multiple digits |
| 256671000006111 | Open fracture-dislocation carpometacarpal joint |
| 256681000006114 | Open fracture-dislocation digit |
| 256691000006112 | Open fracture-dislocation digit, unspecified |
| 256711000006110 | Open fracture-dislocation elbow joint |
| 256721000006119 | Open fracture-dislocation IPJ, unspecified |
| 257181000006118 | Open fracture of tibia and fibula, unspecified part, NOS |
| 257561000006119 | Open fracture proximal femur, transcervical, NOS |
| 258191000006119 | Open fracture of proximal femur, pertrochanteric, NOS |
| 259761000006117 | Open fracture dislocation of sacro-iliac joint |
| 261541000006119 | Open dorsal Barton's fracture-dislocation |
| 264481000006111 | Oligoarticular osteoarthritis, unspecified, of shoulder |
| 264501000006118 | Oligoarticular osteoarthritis, unspecified, other spec sites |
| 265861000006116 | Oligoarticular osteoarthritis, unspecified, multiple sites |
| 318551000006118 | [Q] Open fracture grade 3A |
| 363841000006110 | [X]Arthropathy in hypersensitivity reactions CE |
| 363861000006114 | [X]Arthropathy in other blood disorders CE |
| 377471000006118 | [X]Disl'n sprain/strain unsp joint&ligamt upr limb lvl unsp |
| 377511000006111 | [X]Dislocat/sprains/strains involv oth comb of body regions |
| 405421000006113 | [X]Oth specified acquired deformities/musculoskeletal system |
| 412771000006110 | [X]Other postinfectious arthropathies in diseases CE |
| 422931000006116 | [X]Polyneuropathy in infectious+parasitic diseases CE |
| 422981000006115 | [X]Polyneuropathy/other musculoskeletal disorders CE |
| 424491000006118 | [X]Reactive arthropathy in other diseases CE |
| 427441000006117 | [X]Sprain & strain of oth & unspecif parts of wrist & hand |
| 427491000006114 | [X]Sprain/strain of joint/ligam of oth & unsp part of neck |
| 483991000006118 | Ankylosis of other CMC joint |
| 484091000006115 | Ankylosis of the hip joint |
| 484111000006112 | Ankylosis of the knee joint |
| 484121000006116 | Ankylosis of the shoulder joint |
| 492061000006117 | Arthralgia of tibio-fibular joint |
| 492101000006119 | Arthritis associated with other disease, 1st MTP joint |
| 492191000006114 | Arthritis associated with other disease, lesser MTP joint |
| 492251000006111 | Arthritis associated with other disease, sternoclavic joint |
| 492281000006115 | Arthritis associated with other disease, tibio-fibular joint |
| 492781000006112 | Arthropathy due to haemophilia |
| 493021000006115 | Arthropathy NOS |
| 493161000006113 | Arthropathy with other bacterial disease, of hand |
| 493201000006119 | Arthropathy with other bacterial disease, of pelvic/thigh |
| 493211000006116 | Arthropathy with other bacterial disease, of shoulder region |
| 493221000006112 | Arthropathy with other bacterial disease, of unspec site |
| 493231000006110 | Arthropathy with other bacterial disease, of upper arm |
| 531671000006113 | C5 vertebra open fracture without spinal cord lesion |
| 531711000006112 | C7 vertebra open fracture without spinal cord lesion |
| 538811000006114 | Carpal instability, dorsal subluxation |
| 544811000006116 | Cervico-thoracic ankylosis |
| 556181000006113 | Chronic post-rheumatic arthropathy |
| 561321000006113 | Closed dorsal Barton's fracture-dislocation |
| 561821000006117 | Closed fracture dislocation of sacro-iliac joint |
| 562841000006118 | Closed fracture of coccyx with unspec spinal cord lesion |
| 563961000006116 | Closed fracture of thoracic spine with cord lesion NOS |
| 565241000006110 | Closed fracture-dislocation digit, unspecified |
| 565341000006115 | Closed fracture-dislocation of sternum |
| 565351000006118 | Closed fracture-dislocation other carpal |
| 565361000006116 | Closed fracture-dislocation peri-lunate (dorsal) |
| 565441000006111 | Closed fracture-dislocation, distal interphalangeal joint |
| 565511000006113 | Closed fracture-dislocation, midtarsal joint |
| 565551000006114 | Closed fracture-dislocation, tarsometatarsal joint |
| 565591000006115 | Closed fracture-subluxation elbow |
| 565611000006114 | Closed fracture-subluxation IPJ, unspecified |
| 565651000006110 | Closed fracture-subluxation of pelvis |
| 565801000006112 | Closed fracture-subluxation, hip joint |
| 565821000006119 | Closed fracture-subluxation, IPJ, multiple toes |
| 565871000006118 | Closed fracture-subluxation, patello-femoral joint |
| 567791000006119 | Closed spinal subluxation with cervical cord lesion, unspec |
| 567861000006117 | Closed spinal subluxation with thoracic cord lesion, unspec |
| 569501000006112 | Closed volar Barton fracture-subluxation |
| 570831000006116 | Cls spinal fracture with unspec thoracic cord lesion, T7-12 |
| 583951000006116 | Congenital musculoskeletal deformities |
| 598231000006114 | Costal cartilage sprain |
| 601221000006114 | Crush inj of oth & unspecif parts of abdom/low back/pelv |
| 612291000006116 | Delayed union of fracture |
| 613741000006112 | Depressed skull fracture NOS |
| 632621000006112 | Dupuytren's fracture, fibula |
| 666351000006116 | Familial neuropathic amyloid |
| 693491000006111 | Musculoskeletal or connective tissue diseases OS |
| 694101000006113 | Muscle injury / strain |
| 750531000006118 | Late effect of face fracture |
| 750551000006113 | Late effect of fracture neck of femur |
| 750731000006111 | Late effect of musculoskeletal/connective tissue injury NOS |
| 755441000006119 | Juvenile rheumatoid arthritis - Still's disease |
| 768961000006113 | Intracranial injury, excluding those with skull fracture NOS |
| 779361000006114 | Infective arthritis NOS, of distal radio-ulnar joint |
| 779391000006118 | Infective arthritis NOS, of IP joint of toe |
| 779471000006110 | Infective arthritis NOS, of sacro-iliac joint |
| 780201000006117 | Infective arthritis NOS, of 1st MTP joint |
| 792901000006112 | Fracture of lower leg, part unspecified |
| 828201000006113 | Horton's (histamine) neuralgia |
| 309700017 | Crystal arthropathy NOS, of the shoulder region |
| 309708012 | Crystal arthropathy NOS, of sternoclavicular joint |
| 309731012 | Crystal arthropathy NOS, of multiple sites |
| 318602018 | Fracture of rib(s), sternum, larynx and trachea |
| 603481000006111 | Crystal arthropathy NOS, of DIP joint of finger |
| 603491000006114 | Crystal arthropathy NOS, of distal radio-ulnar joint |
| 603541000006116 | Crystal arthropathy NOS, of lesser MTP joint |
| 603551000006119 | Crystal arthropathy NOS, of MCP joint |
| 778621000006116 | Inflammatory polyneuropathy, unspecified |
| 913981000006115 | Insulin dependent diab mell with neuropathic arthropathy |
| 310055010 | Traumatic arthropathy of the lower leg |
| 400225016 | Joint ankylosis of the lower leg |
| 892411000006116 | Path.fracture - ankle/foot |
| 892431000006110 | Path.fracture - pelvis/thigh |
| 989021000006118 | Other peripheral neuropathy |
| 312681016 | [X]Other osteoporosis with pathological fracture |
| 130991000006117 | Sprain thumb |
| 212321000006111 | Postprocedural musculoskeletal disorder, unspecified |
| 318480017 | Open fracture pelvis, coccyx |
| 319204018 | Closed fracture tubercle, tibia |
| 402849015 | Fracture of vertebra with spinal cord lesion |
| 461576019 | [Q] Central spinal stenosis |
| 318600014 | Open fracture of spine with spinal cord lesion unspecified |
| 461007015 | [V]Unspecified psychological or physical strain |
| 363831000006117 | [X]Arthropathies/oth endocrin,nutritionl+metabolic disorders |
| 416711000006117 | [X]Other superfic injuries of abdomen, lower back & pelvis |
| 297571012 | Other toxic agent polyneuropathy |
| 309522010 | Arthropathy associated with other viral diseases |
| 309527016 | Arthropathy with other viral disease, of hand |
| 309533013 | Arthropathy associated with other viral disease NOS |
| 309915010 | Localised, secondary osteoarthritis of unspecified site |
| 309930018 | Localised, secondary osteoarthritis of the lower leg |
| 309949019 | Localised osteoarthritis, unspecified, of the upper arm |
| 309971012 | Localised osteoarthritis, unspecified, NOS |
| 310109010 | Transient arthropathy of the forearm |
| 310550011 | Ankylosis of other pelvic joint |
| 312494011 | Other specified musculoskeletal disorders |
| 318632014 | Closed fracture of larynx and trachea NOS |
| 1230477016 | Open fracture distal humerus, medial epicondyle |
| 130571000006114 | Sprain & strain of other and unspecified parts of thorax |
| 217751000006115 | Postdysenteric reactive arthropathy of the upper arm |
| 333341000006111 | [SO]Specified part of musculoskeletal system NEC |
| 493241000006117 | Arthropathy with other viral disease, of ankle and foot |
| 493281000006111 | Arthropathy with other viral disease, of multiple sites |
| 493291000006114 | Arthropathy with other viral disease, of other spec site |
| 493301000006110 | Arthropathy with other viral disease, of pelvic region/thigh |
| 493311000006113 | Arthropathy with other viral disease, of shoulder region |
| 493321000006117 | Arthropathy with other viral disease, of unspecified site |
| 312516012 | [X]Inflammatory polyarthropathies |
| 877711000006118 | Splinting of fracture |
| 889871000006117 | Osteoarthritis -shoulder joint |
| 889921000006110 | Osteoarthritis - knee joint |
| 889941000006115 | Osteoarthritis - other joint |
| 890411000006115 | Ankylosis - wrist joint |
| 890461000006117 | Ankylosis - other joint |
| 891021000006111 | Back disorder/symptom NOS |
| 891571000006115 | Rheumatism/fibrositis NOS |
| 891601000006110 | Rheumatism NOS - hip |
| 891641000006112 | Rheumatism NOS - shoulder |
| 891681000006118 | Myalgia/myositis - lower leg |
| 891691000006115 | Myalgia/myositis -pelvis/thigh |
| 891701000006115 | Myalgia/myositis - hand |
| 891711000006117 | Myalgia/myositis - fore-arm |
| 891721000006113 | Myalgia/myositis - upper arm |
| 891841000006113 | Neuralgia/neuritis - multiple |
| 891851000006110 | Neuralgia/neuritis NOS |
| 892511000006115 | Fracture malunion - lower leg |
| 892531000006114 | Fracture malunion - hand |
| 892551000006119 | Fracture malunion - upper arm |
| 892711000006110 | Musculoskeletal problems NOS |
| 892731000006116 | Musculoskeletal disease NOS |
| 894711000006115 | Headache symptom NOS [D] |
| 895921000006112 | Fractures |
| 896181000006117 | Sprain - upper arm |
| 989341000006110 | Osteoarthritis - elbow joint |
| 989351000006112 | Osteoarthritis - wrist joint |
| 989621000006112 | Multiple fractures |
| 990171000006111 | Sprain - lower leg |
| 990331000006110 | Sprain - upper arm |
| 991371000006119 | Other sprains NOS |
| 991401000006116 | Sprained knee NOS |
| 1464018 | Monoarthritis |
| 40395011 | Skull base fracture |
| 216206018 | Pathological fracture due to osteoporosis |
| 257899015 | Rheumatoid arthritis latex test |
| 297165012 | Disease related peripheral neuropathy |
| 297338014 | Cluster headache syndrome |
| 297427010 | Trigeminal neuralgia |
| 297528015 | Idiopathic peripheral neuropathy |
| 299360019 | Mononeuropathy of lower limb |
| 309400018 | Arthropathy |
| 309972017 | Osteoarthritis of multiple joints |
| 310012011 | Osteoarthritis of sternoclavicular joint |
| 310013018 | Osteoarthritis of acromioclavicular joint |
| 310028018 | Osteoarthritis of talonavicular joint |
| 310855014 | Arthralgia |
| 311041010 | Sacroiliitis |
| 311045018 | Spondylitis |
| 311688013 | Rheumatism |
| 311707014 | Neuritis |
| 312504018 | Musculoskeletal disorder |
| 312510018 | Streptococcal arthritis |
| 312521010 | Psoriasis with arthropathy |
| 312703010 | Hypertrophic osteoarthropathy |
| 312729010 | Musculoskeletal and connective tissue disorder |
| 317264010 | [D]Headache |
| 318196011 | Fracture of base of skull |
| 318224014 | Open fracture of mandible |
| 318493010 | Closed fracture of vertebral column |
| 318603011 | Closed fracture of rib |
| 318614014 | Open fracture of rib |
| 318690016 | Multiple closed fractures of pelvis with disruption of pelvic circle |
| 318718016 | Closed fracture of pelvis |
| 318773018 | Closed fracture of humerus |
| 318785014 | Closed fracture of medial condyle of humerus |
| 318962017 | Closed fracture of metacarpal bone |
| 319052019 | Fracture of phalanx of finger |
| 319146018 | Fracture of femur |
| 319241017 | Closed fracture of tibia |
| 319248011 | Open fracture of tibia AND fibula |
| 319271013 | Closed fracture of ankle |
| 319332012 | Fracture of phalanx of foot |
| 319333019 | Fracture of lower limb |
| 319337018 | Fractures of multiple bones of lower limb |
| 319351018 | Closed fracture of bone |
| 320109011 | Fracture dislocation of joint |
| 320143019 | Elbow sprain |
| 320147018 | Wrist sprain |
| 320220017 | Sprain of wrist and/or hand |
| 320234017 | Sprain of hip |
| 320318018 | Back sprain |
| 320560015 | Sprained rib |
| 325382011 | Fracture of shoulder |
| 325451011 | Sprain of knee |
| 325460015 | Fracture of lower leg |
| 325468010 | Fracture of tarsal bone |
| 325691019 | Late effect of fracture of lower extremities |
| 359167017 | Arthritis of foot |
| 359388019 | Osteoarthrosis of the carpometacarpal joint of the thumb |
| 391249010 | Ligament sprain |
| 391357011 | Fracture of ulna |
| 391373017 | Sternal fracture |
| 400188013 | Localised, primary osteoarthritis of the ankle and/or foot |
| 400263012 | Arthralgia of the ankle and/or foot |
| 400314010 | Myalgia |
| 400878010 | Congenital anomaly of musculoskeletal system |
| 402860012 | Closed fracture of distal end of radius |
| 402862016 | Open fracture of distal end of radius |
| 403161016 | Fracture |
| 406699016 | Ankle instability |
| 461055015 | Convalescence after fracture treatment |
| 18051000006110 | Osteoarthritis of proximal interphalangeal joint |
| 40871000006119 | Osteoporosis with pathological fracture of cervical vertebrae |
| 40891000006118 | Osteoporosis with pathological fracture of thoracic vertebrae |
| 41301000006112 | Osteoarthritis of first metatarsophalangeal joint |
| 41971000006117 | Fracture of orbital roof |
| 44421000006110 | Open spinal fracture with anterior cervical cord lesion, C5-7 |
| 52931000006111 | Open fracture of vault of skull with intracranial injury, with more than 24 hours loss of consciousness and return to pre-existing conscious level |
| 52941000006118 | Open fracture of vault of skull with intracranial injury, with 1-24 hours loss of consciousness |
| 54541000006116 | Closed fracture of wrist |
| 87721000006119 | Trigeminal nerve disorder |
| 92671000006112 | Traumatic arthropathy of lesser metatarsophalangeal joint |
| 106921000006111 | Thoracic back sprain |
| 130631000006110 | Sprain finger, distal interphalangeal joint, ulnar collateral ligament |
| 130641000006117 | Sprain finger, metacarpophalangeal joint, radial collateral ligament |
| 130651000006115 | Sprain finger, metacarpophalangeal joint, ulnar collateral ligament |
| 130671000006113 | Sprain finger, proximal interphalangeal joint, ulnar collateral ligament |
| 130711000006112 | Sprain finger, proximal interphalangeal joint, nonspecific |
| 130721000006116 | Strain of gastrocnemius tendon |
| 130931000006116 | Sprain of scapholunate ligament |
| 131171000006111 | Sprain of wrist |
| 131391000006114 | Sprain of interphalangeal joint of toe |
| 131401000006111 | Sprain of lateral collateral ligament of knee |
| 131461000006112 | Strain of patellar tendon |
| 131481000006119 | Sprain, posterior sacroiliac ligament |
| 131551000006111 | Strain of subscapularis tendon |
| 131561000006113 | Strain of supraspinatus tendon |
| 131591000006117 | Strain of Achilles tendon |
| 140551000006117 | Arthritis of shoulder region joint |
| 155721000006112 | Injury of sciatic nerve |
| 159761000006115 | Sprain of sacrococcygeal ligament |
| 159821000006119 | Sprain of ligament of sacroiliac joint |
| 162101000006115 | Rheumatoid arthritis of lesser metatarsophalangeal joint |
| 162111000006117 | Rheumatoid arthritis of metacarpophalangeal joint |
| 162641000006113 | Sprain of costal cartilage |
| 168241000006113 | Rheumatology follow-up assessment |
| 168261000006112 | Rheumatology symptom change |
| 192631000006118 | Reactive arthritis |
| 192711000006116 | Reactive arthropathy of interphalangeal joint of toe |
| 192731000006110 | Reactive arthropathy of lesser metatarsophalangeal joint |
| 198881000006116 | Psoriatic spondylitis |
| 211231000006113 | Post-herpetic polyneuropathy |
| 219261000000112 | Crystal arthropathy |
| 221511000000115 | Osteoarthritis of hip |
| 222361000000115 | Pott's fracture of ankle |
| 244171000006112 | Fracture of parietal bone |
| 253461000006114 | Outfracture of nasal turbinate |
| 256091000006119 | Open fracture dislocation, foot |
| 256131000006117 | Open fracture dislocation, interphalangeal joint, single toe |
| 257781000006117 | Open fracture of upper end of humerus |
| 258471000006114 | Open intertrochanteric fracture |
| 259321000006112 | Open fracture of shaft of metacarpal bone |
| 259461000006112 | Open fracture of head of femur |
| 259601000006113 | Open fracture of base of skull with intracranial injury |
| 259881000006117 | Open fracture of medial condyle of humerus |
| 264141000006110 | Open fracture of base of skull with intracranial injury, with 1-24 hours loss of consciousness |
| 264431000006110 | Open fracture of vault of skull with intracranial injury, with less than 1 hour loss of consciousness |
| 265451000006116 | Open fracture of base of skull with intracranial injury, with more than 24 hours loss of consciousness without return to pre-existing conscious level |
| 268071000006113 | Occipital fracture |
| 359201000006116 | Tension-type headache |
| 394571000006116 | Chronic arthritis of juvenile onset |
| 428461000006117 | [X]Symptoms and signs involving the nervous and musculoskeletal systems |
| 483971000006119 | Ankylosis of metatarsophalangeal joint |
| 484031000006119 | Ankylosis of proximal interphalangeal joint |
| 491791000006117 | Arthralgia of 1st metatarsophalangeal joint |
| 491861000006115 | Arthralgia of interphalangeal joint of toe |
| 491891000006111 | Arthralgia of metacarpophalangeal joint |
| 492521000006116 | Arthropathy associated with non-infective gastrointestinal disorders |
| 492911000006119 | Arthropathy in Behcet's syndrome of the pelvic region and thigh |
| 498181000006114 | Atrophic nonunion of fracture |
| 531611000006116 | Open fracture of second cervical vertebra without mention of spinal cord lesion |
| 531641000006117 | Closed fracture of fourth cervical vertebra without spinal cord injury |
| 531681000006111 | Closed fracture of sixth cervical vertebra without spinal cord injury |
| 531701000006114 | Closed fracture of seventh cervical vertebra without spinal cord injury |
| 557941000006115 | Closed fractures involving multiple regions of both upper limbs |
| 559951000006113 | Closed fracture of base of skull with intracranial injury, with more than 24 hours loss of consciousness without return to pre-existing conscious level |
| 561441000006114 | Closed bimalleolar fracture |
| 562171000006118 | Closed fracture of shaft of metacarpal bone |
| 562981000006117 | Closed intertrochanteric fracture |
| 563371000006110 | Closed fracture of phalanx of toe |
| 563891000006116 | Closed fracture of upper end of tibia |
| 564071000006114 | Closed fracture of tibia AND fibula |
| 564101000006116 | Closed fracture of tibia or fibula, shaft |
| 564111000006118 | Closed fracture of condyle of tibia |
| 565191000006110 | Closed fracture of vault of skull without intracranial injury |
| 565221000006115 | Closed fracture dislocation acromioclavicular joint |
| 565391000006112 | Closed fracture dislocation shoulder joint |
| 565421000006116 | Closed fracture dislocation, ankle joint |
| 565501000006110 | Closed fracture dislocation, metacarpophalangeal joint |
| 565671000006117 | Closed fracture subluxation of the wrist |
| 565731000006112 | Closed fracture subluxation shoulder joint |
| 565841000006114 | Closed fracture subluxation, knee joint |
| 567631000006110 | Closed spinal dislocation with posterior cervical cord lesion |
| 570561000006111 | Closed fracture proximal femur, intertrochanteric, comminuted |
| 570671000006117 | Closed spinal fracture with anterior cervical cord lesion, C1-4 |
| 570711000006118 | Closed spinal fracture with central cervical cord lesion, C5-7 |
| 570721000006114 | Closed spinal fracture with central thoracic cord lesion, T1-6 |
| 570731000006112 | Closed spinal fracture with central thoracic cord lesion, T7-12 |
| 570751000006117 | Closed spinal fracture with complete cervical cord lesion, C5-7 |
| 570771000006110 | Closed spinal fracture with posterior cervical cord lesion, C1-4 |
| 570791000006111 | Closed spinal fracture with posterior thoracic cord lesion, T1-6 |
| 570801000006112 | Closed spinal fracture with posterior thoracic cord lesion, T7-12 |
| 570811000006110 | Closed fracture of C1-C4 level with spinal cord injury |
| 623361000006116 | Dislocations, sprains and strains involving thorax with lower back and pelvis |
| 623391000006112 | Dislocations, sprains and strains involving multiple regions of lower limb(s) |
| 623401000006114 | Dislocations, sprains and strains involving multiple regions of upper limb(s) and lower limb(s) |
| 694981000006118 | Multiple fractures of skull |
| 702621000006114 | Fracture of middle fossa |
| 777831000006118 | Injury of nerves and lumbar spinal cord at abdomen, lower back and pelvis level |
| 792961000006113 | Closed fracture of malar AND/OR maxillary bones |
| 793501000006112 | Fracture of shaft of radius and ulna |
| 793671000006119 | Fracture of transverse process of spine with spinal cord lesion |
| 793901000006115 | Fracture dislocation or subluxation hip |
| 793921000006113 | Fracture dislocation or subluxation of wrist |
| 794881000006119 | Fracture of frontal bone |
| 800731000006117 | Generalised osteoarthritis |
| 800751000006112 | Polyarticular osteoarthritis |
| 807281000006112 | Groin strain |
| 819781000006110 | Arthropathy associated with helminthiasis |
| 1715221000006117 | Cervicogenic headache |
| 1716421000006114 | Chronic headache disorder |
| 1747081000000119 | History of migraine with aura |
| 1786091000006118 | Benign exertion headache |
| 1871131000006116 | Lower back injury |
| 309703015 | Crystal arthropathy of hand |
| 309706011 | Crystal arthropathy of ankle AND/OR foot |
| 309707019 | Crystal arthropathy of shoulder region |
| 309718019 | Crystal arthropathy of hip |
| 309722012 | Crystal arthropathy of ankle AND/OR foot |
| 318040019 | [X]Other chronic pain |
| 318902015 | Closed fracture of forearm |
| 319145019 | Open fracture of neck of femur |
| 455736015 | Multiple fractures |
| 483745010 | Closed fracture of shaft of radius and ulna |
| 16911000006114 | Rheumatoid arthritis with organ / system involvement |
| 221661000006114 | Inflammatory polyarthritis |
| 563401000006113 | Closed fracture of one or more tarsal and metatarsal bones |
| 451461014 | Seropositive erosive rheumatoid arthritis |
| 2839291014 | Disease activity score 28 joint in rheumatoid arthritis |
| 310944013 | Arthropathy of ankle and/or foot |
| 359261000006115 | Periprosthetic fracture |
| 318831017 | Closed fracture of shaft of radius and/or ulna |
| 256415010 | Musculoskeletal test abnormal |
| 318479015 | Closed fracture of coccyx |
| 461537015 | Buckle fracture |
| 1222484010 | Musculoskeletal symptom |
| 318601013 | Fracture of vertebral column with spinal cord injury |
| 422921000006119 | [X]Polyneuropathies and other disorders of the peripheral nervous system |
| 890401000006118 | Ankylosis - elbow joint |
| 890441000006116 | Ankylosis - knee joint |
| 891581000006117 | Rheumatism NOS - ankle/foot |
| 891591000006119 | Rheumatism NOS - knee |
| 891631000006119 | Rheumatism NOS - elbow |
| 891821000006118 | Neuralgia/neuritis - upper arm |
| 892501000006118 | Fracture malunion - ankle/foot |
| 892521000006111 | Fracture malunion-pelvis/thigh |
| 892541000006116 | Fracture malunion - fore arm |
| 892571000006112 | Multiple fracture malunion |
| 896251000006117 | Sprain - hand NOS |
| 896451000006116 | Sprain - back |
| 896511000006116 | Other sprains NOS |
| 990321000006112 | Sprained shoulder |
| 461544012 | [Q] Refracture |
| 309841017 | Other specified inflammatory polyarthropathy |
| 309919016 | Localised, secondary osteoarthritis of the upper arm |
| 309945013 | Localised osteoarthritis, unspecified, of unspecified site |
| 318799013 | Open fracture of distal humerus, condyle(s) unspecified |
| 427451000006115 | [X]Sprain & strain of other and unspecified parts of thorax |
| 736701000006117 | Localised osteoarthritis, unspecified, of other spec site |
| 319216012 | Open fracture tubercle, tibia |
| 402850015 | Fracture of spine with spinal cord lesion |
| 461578018 | [Q] Central and lateral spinal stenosis |
| 377571000006119 | [X]Dislocation, sprain and strain of unspecif body region |
| 792591000006110 | Fracture of bony thorax, part unspecified |
| 309710014 | Crystal arthropathy NOS, of acromioclavicular joint |
| 772131000006111 | Insulin dependent diab mell with neuropathic arthropathy |
| 455509015 | [X]Open multiple fractures unspecified |
| 883171000006110 | Other peripheral neuropathy |
| 359071000006116 | [X] Polyneuropathy, unspecified |
| 366601000006119 | [X]Autonomic neuropathy/endocrine+metabolic diseases CE |
| 399091000006110 | [X]Multiple fractures of clavicle, scapula and humerus |
| 408441000006111 | [X]Other congenital malforms of the musculoskeletal system |
| 422971000006118 | [X]Polyneuropathy/other endocrine+metabolic diseases CE |
| 423971000006114 | [X]Psychogenic headache |
| 484071000006116 | Ankylosis of the ankle joint |
| 492221000006119 | Arthritis associated with other disease, PIP joint of finger |
| 493181000006115 | Arthropathy with other bacterial disease, of multiple sites |
| 562751000006110 | Closed fracture of cervical spine with cord lesion NOS |
| 565271000006119 | Closed fracture-dislocation elbow joint |
| 565661000006112 | Closed fracture-subluxation of sternum |
| 565701000006116 | Closed fracture-subluxation peri-lunate trans-scaphoid |
| 565751000006117 | Closed fracture-subluxation, ankle joint |
| 565861000006113 | Closed fracture-subluxation, midtarsal joint |
| 565881000006115 | Closed fracture-subluxation, proximal interphalangeal joint |
| 570661000006112 | Cls spinal # with incomplete thoracid cord lesion, T7-12 NOS |
| 603521000006111 | Crystal arthropathy NOS, of IP joint of toe |
| 603721000006119 | Crystal arthropathy NOS, of tibio-fibular joint |
| 623781000006113 | Dislocations, sprains & strains involv multiple body regions |
| 631181000006113 | Drug-induced headache, not elsewhere classified |
| 632641000006117 | Dupuytren's fracture, radius - open |
| 673591000006119 | Nodular fibrositis of chronic rheumatic disease |
| 695771000006119 | Multiple fractures involving skull/face with other bones NOS |
| 25171000006116 | Other open fracture-dislocation |
| 43231000006111 | Opn spinal fracture with unspec thoracic cord lesion, T7-12 |
| 49861000006117 | Open volar Barton fracture-subluxation |
| 89511000006113 | Tuberculosis of meninges and central nervous system |
| 105741000006114 | Thigh fracture NOS |
| 130911000006110 | Sprain radio-scapho-capitate ligament |
| 131211000006113 | Sprain, anterior sacro-iliac ligament |
| 131451000006110 | Sprain, mid tarsal joint |
| 211281000006114 | Postimmunization arthropathy |
| 211341000006110 | Postinfective arthropathy in syphilis |
| 214871000006114 | Polyneuropathy due to drugs |
| 217711000006116 | Postdysenteric reactive arthropathy of the hand |
| 255591000006115 | Open fracture-subluxation shoulder |
| 255601000006111 | Open fracture-subluxation shoulder joint |
| 255621000006118 | Open fracture-subluxation sterno-clavicular joint, posterior |
| 255681000006119 | Open fracture-subluxation, foot |
| 255701000006116 | Open fracture-subluxation, interphalangeal joint thumb |
| 255721000006114 | Open fracture-subluxation, IPJ, single toe |
| 255731000006112 | Open fracture-subluxation, knee joint |
| 255761000006115 | Open fracture-subluxation, midtarsal joint |
| 256051000006113 | Open fracture-dislocation superior radio-ulnar joint |
| 256311000006114 | Open fracture-subluxation of pelvis |
| 318541000006115 | [Q] Open fracture grade 3 |
| 318561000006116 | [Q] Open fracture grade 3B |
| 299346016 | [X]Vascular headache, not elsewhere classified |
| 309463019 | Pyogenic arthritis of other specified sites |
| 309484012 | Arthropathy in Behcet's syndrome of the upper arm |
| 309521015 | Arthropathy associated with other bacterial disease NOS |
| 309534019 | Arthropathy associated with mycoses, of unspecified site |
| 309536017 | Arthropathy associated with mycoses, of the upper arm |
| 309594016 | Infective arthritis NOS |
| 309595015 | Infective arthritis NOS, of unspecified site |
| 309598018 | Infective arthritis NOS, of the forearm |
| 309599014 | Infective arthritis NOS, of the hand |
| 309620013 | Infective arthritis NOS, of talonavicular joint |
| 309621012 | Infective arthritis NOS, of other tarsal joint |
| 309693017 | Other crystal arthropathies of the ankle and foot |
| 309828016 | Juvenile rheumatoid arthritis NOS |
| 309899016 | Localised, primary osteoarthritis of other specified site |
| 309976019 | Oligoarticular osteoarthritis, unspecified, of forearm |
| 310051018 | Traumatic arthropathy of the upper arm |
| 310091015 | Allergic arthritis NOS |
| 310150019 | Unspecified polyarthropathy of the upper arm |
| 310151015 | Unspecified polyarthropathy of the forearm |
| 310166017 | Unspecified monoarthritis of unspecified site |
| 310171012 | Unspecified monoarthritis of the pelvic region and thigh |
| 310173010 | Unspecified monoarthritis of the ankle and foot |
| 310190016 | Other specified arthropathy of other specified site |
| 310784012 | Palindromic rheumatism of the lower leg |
| 312513016 | [X]Other reactive arthropathies |
| 312959012 | Other specified spinal cord anomalies NOS |
| 314955017 | Other congenital musculoskeletal anomalies |
| 318203016 | Closed fracture of mandible, ramus, unspecified |
| 318680012 | Closed fracture pubis NOS |
| 318697018 | Other or multiple closed fracture of pelvis NOS |
| 318734018 | Fracture of clavicle NOS |
| 318735017 | Closed fracture of scapula, unspecified part |
| 318828018 | Open fracture of forearm, upper end, NOS |
| 318838011 | Closed fracture of forearm, lower end, unspecified |
| 318849017 | Closed Smith's fracture |
| 318934012 | Open fracture of other carpal bone |
| 318964016 | Open fracture of metacarpal bone(s), site unspecified |
| 319050010 | Open fracture of one or more phalanges of hand NOS |
| 319079015 | Fracture of upper limb, level unspecified |
| 319080017 | Fracture of upper limb NOS |
| 319247018 | Open fracture of fibula, unspecified part, NOS |
| 319250015 | Fracture of tibia and fibula, NOS |
| 320281016 | Ankle and foot sprain NOS |
| 320550012 | Thyroid region sprain, unspecified |
| 325399012 | [X]Fracture of other parts of forearm |
| 378099017 | Endemic polyarthritis |
| 391381016 | Lower leg fracture NOS |
| 400157014 | Pyogenic arthritis of the upper arm |
| 400184010 | Osteoarthritis NOS, of the forearm |
| 400194017 | Arthropathy NOS |
| 400195016 | Arthropathy NOS, of the upper arm |
| 400196015 | Arthropathy NOS, of the forearm |
| 402869013 | Closed fracture proximal femur, other transcervical |
| 402909010 | Other shoulder sprain |
| 39246018 | Stress fracture |
| 158050019 | Polymyalgia |
| 252315012 | Back pain without radiation NOS |
| 277163010 | Other primary external immobilisation of fracture |
| 295346011 | Psychogenic musculoskeletal symptoms NOS |
| 297519016 | Hereditary peripheral neuropathy NOS |
| 55627011 | Psoriatic arthropathy |
| 311696015 | Myalgia and myositis unspecified |
| 1174311000000111 | Arthropathy in cystic fibrosis |
| 875411000006118 | Refracture of bone-osteoclasis |
| 895291000006119 | #Lumbar spine - no cord lesion |
| 895321000006111 | #Cervical spine + cord lesion |
| 895341000006116 | #Lumbar spine + cord lesion |
| 895631000006116 | #Bennett's fracture |
| 896221000006114 | Sprain - hand |
| 2478835016 | Closed fracture of mandible, alveolar border of body |
| 2536029011 | Fracture of thumb |
| 162261000006114 | Rheumatoid factor |
| 359289013 | Arthropathy following intestinal bypass |
| 359312010 | Juvenile ankylosing spondylitis |
| 391100012 | Leg sprain |
| 391101011 | Back sprain excluding lumbosacral |
| 391397014 | Closed fracture of femur, distal end |
| 391398016 | Open fracture of femur, distal end |
| 391414017 | Fracture of calcaneus |
| 391416015 | Heel bone fracture |
| 400159012 | Pyogenic arthritis of the pelvic region and thigh |
| 400250018 | Arthralgia of the lower leg |
| 402832010 | Fracture of spine without mention of spinal cord injury |
| 402834011 | Closed fracture atlas |
| 402847018 | Open fracture of sixth cervical vertebra |
| 402848011 | Open fracture of seventh cervical vertebra |
| 419562016 | Fracture of medial malleolus |
| 426510015 | Rheumatoid arthritis - multiple joint |
| 444977014 | Nonunion of fracture |
| 451061019 | Closed Barton's fracture |
| 451471011 | Closed fracture of distal fibula |
| 455452018 | Open fracture of great toe |
| 309460016 | Pneumococcal arthritis and polyarthritis |
| 309535018 | Arthropathy associated with mycoses, of the shoulder region |
| 309562014 | Reactive arthropathy of sternoclavicular joint |
| 309680010 | Gouty arthritis of the ankle and foot |
| 309802019 | Rheumatoid arthritis of ankle |
| 309827014 | Monarticular juvenile rheumatoid arthritis |
| 309923012 | Localised, secondary osteoarthritis of the hand |
| 310073013 | Traumatic arthropathy of subtalar joint |
| 310090019 | Allergic arthritis of multiple sites |
| 310096013 | Climacteric arthritis of the shoulder region |
| 310782011 | Palindromic rheumatism of the hand |
| 311196019 | Iatrogenic cervical spinal stenosis |
| 311237018 | Idiopathic lumbar spinal stenosis |
| 311290019 | Cervical spine instability |
| 311694017 | Hand rheumatism |
| 311695016 | Rheumatism or fibrositis NOS |
| 312157010 | Osteoporosis of disuse with pathological fracture |
| 312801016 | Cervical spinal meningocele |
| 318197019 | Fracture of mandible, closed |
| 318202014 | Closed fracture of mandible, coronoid process |
| 318205011 | Closed fracture of mandible, symphysis of body |
| 318376011 | Closed fracture atlas, comminuted |
| 318378012 | Closed fracture axis, spondylolysis |
| 318429018 | Open fracture cervical vertebra, spinous process |
| 318448019 | Open fracture thoracic vertebra, wedge |
| 318474013 | Closed compression fracture sacrum |
| 318682016 | Open fracture pelvis, single pubic ramus |
| 318693019 | Closed fracture pelvis, anterior inferior iliac spine |
| 318702010 | Open fracture pelvis, ischial tuberosity |
| 318704011 | Open fracture pelvis, anterior inferior iliac spine |
| 318736016 | Closed fracture scapula, coracoid |
| 318757012 | Closed fracture proximal humerus, four part |
| 318800012 | Open fracture of distal humerus, trochlea |
| 318830016 | Closed fracture radius and ulna, middle |
| 318851018 | Closed volar Barton's fracture |
| 318888015 | Open fracture radial styloid |
| 318932011 | Open fracture scaphoid, tuberosity |
| 318969014 | Open fracture finger metacarpal head |
| 318970010 | Open fracture finger metacarpal |
| 319001016 | Closed fracture finger proximal phalanx, shaft |
| 319006014 | Closed fracture finger middle phalanx, base |
| 319023017 | Open fracture thumb proximal phalanx, base |
| 319025012 | Open fracture thumb proximal phalanx, neck |
| 319038016 | Open fracture finger middle phalanx |
| 319040014 | Open fracture finger middle phalanx, shaft |
| 319041013 | Open fracture finger middle phalanx, neck |
| 319042018 | Open fracture finger middle phalanx, head |
| 319043011 | Open fracture finger middle phalanx, multiple |
| 319086011 | Closed fracture proximal femur, transepiphyseal |
| 319112019 | Open fracture proximal femur,subcapital, Garden grade IV |
| 319134017 | Open fracture proximal femur, subtrochanteric |
| 319135016 | Open fracture proximal femur, intertrochanteric, comminuted |
| 319162012 | Closed fracture distal femur, medial condyle |
| 319189019 | Open fracture patella, transverse |
| 319218013 | Open fracture fibula, neck |
| 319253018 | Closed fracture ankle, lateral malleolus, high |
| 319255013 | Open fracture ankle, lateral malleolus, high |
| 319268017 | Open fracture ankle, trimalleolar, high fibular fracture |
| 319286019 | Open fractures calcaneus, intra-articular |
| 319293015 | Closed fracture talus, head |
| 319294014 | Closed fracture talus, neck |
| 319295010 | Closed fracture talus, body |
| 319308017 | Open fracture metatarsal base |
| 319309013 | Open fracture metatarsal shaft |
| 319310015 | Open fracture metatarsal neck |
| 319326019 | Open fracture distal phalanx, toe |
| 319716011 | Closed spinal dislocation with central cervical cord lesion |
| 319768016 | Closed spinal dislocation with complete lumbar cord lesion |
| 319769012 | Closed spinal dislocation with anterior lumbar cord lesion |
| 320184016 | Sprain thumb, carpometacarpal joint |
| 320210010 | Sprain tendon wrist or hand |
| 320228012 | Ischiocapsular sprain |
| 320250013 | Sprain, plantaris tendon |
| 320279018 | Sprain, extensor tendon, foot |
| 320551011 | Cricoarytenoid sprain |
| 320552016 | Cricothyroid sprain |
| 276492011 | Primary cast stabilisation of spinal fracture |
| 277216017 | Primary cast immobilisation of fracture |
| 293128014 | Tuberculous arthritis |
| 297540015 | Polyneuropathy in collagen vascular disease |
| 297558014 | Polyneuropathy in hypoglycaemia |
| 303172010 | Irritable bowel syndrome with diarrhoea |
| 252313017 | Back pain worse on sneezing |
| 253119015 | Throbbing headache |
| 253120014 | Shooting headache |
| 253121013 | Morning headache |
| 6700013 | Open fracture of astragalus |
| 6701012 | Open fracture of talus |
| 9300010 | Sinus headache |
| 16067019 | Familial amyloid polyneuropathy, Iowa type |
| 16455010 | Gouty neuritis |
| 27272011 | Fracture of ankle |
| 29902012 | Median nerve neuritis |
| 39287017 | Fracture of upper limb |
| 74959012 | Open fracture of one rib |
| 93295014 | Common migraine |
| 93296010 | Atypical migraine |
| 96628016 | Fracture of clavicle |
| 96705013 | Traumatic arthropathy |
| 104572011 | Arthropathy in Behcet's syndrome |
| 111967016 | Instability of joint |
| 114777011 | Closed fracture of trachea |
| 123686019 | Late effect of intracranial injury without skull fracture |
| 125993015 | Fracture of radius AND ulna |
| 147293015 | Closed fracture of thyroid cartilage |
| 149683011 | Tuberculosis of cerebral meninges |
| 158459012 | Chronic paroxysmal hemicrania |
| 166489012 | Unilateral headache |
| 194543011 | Fracture of coccyx |
| 477671011 | Sarcoid arthropathy |
| 485678014 | Arthritis in Lyme disease |
| 498448012 | Neuropathic foot ulcer |
| 499415017 | Behcet's syndrome arthropathy |
| 507743013 | Hand sprain |
| 511384012 | Closed fracture of carpal bone |
| 1220597016 | Closed fracture multiple ribs |
| 1221377017 | Closed fracture clavicle, lateral end |
| 1222298017 | Arthritis due to rubella |
| 1223079017 | [Q] Stress fracture |
| 1228159014 | Closed fracture ankle, lateral malleolus |
| 1229890019 | Open fracture trapezoid |
| 1230391011 | Open fracture ankle, lateral malleolus |
| 1230712018 | Foot sprain |
| 1232779012 | Open fracture of the radial shaft |
| 1235538018 | Closed fracture capitate |
| 1235653017 | British type amyloid polyneuropathy |
| 1235997015 | Closed fracture cuboid |
| 1485124017 | FH: Hip fracture in first degree relative |
| 1485125016 | FH: Fragility fracture |
| 1488516017 | Rheumatology |
| 1494989015 | Lumbosacral sprain |
| 1776248011 | Osteoarthritis |
| 1786700015 | Periarthritis of shoulder |
| 2472447010 | Juvenile rheumatoid arthritis |
| 309695012 | Crystal arthropathy of multiple sites |
| 309829012 | Juvenile arthritis |
| 310170013 | Monoarthritis of hand |
| 311398011 | Periarthritis |
| 312234010 | Fracture malunion |
| 317098012 | Chronic intractable pain |
| 318244015 | Closed fracture of facial bone |
| 318814012 | Closed fracture of upper end of forearm |
| 318891015 | Open intra-articular fracture of distal radius |
| 318911015 | Open fracture of forearm |
| 319292013 | Closed fracture of tarsal bone |
| 319355010 | Fracture of bone |
| 320130019 | Sprain of shoulder |
| 325470018 | Sprain of foot |
| 359169019 | Arthritis of knee |
| 359170018 | Arthritis of hip |
| 359399017 | Primary generalised osteoarthrosis |
| 391347010 | Fracture of radius |
| 402864015 | Closed fracture of ulna |
| 402917019 | Sprain of spinal ligament |
| 40781000006114 | Osteopathies, chondropathies and acquired musculoskeletal deformities |
| 99281000006115 | Toxic neuropathy, NOS |
| 131061000006115 | Sprain thumb, metacarpophalangeal joint nonspecific |
| 131441000006113 | Sprain, metatarsophalangeal joint |
| 162081000006111 | Rheumatoid arthritis of interphalangeal joint of toe |
| 162211000006111 | Rheumatoid arthritis screen |
| 168301000006115 | Rheumatology disorder treatment started |
| 192671000006115 | Reactive arthropathy of distal interphalangeal joint of finger |
| 256011000006112 | Open fracture dislocation shoulder joint |
| 327591000006117 | Musculoskeletal system |
| 483931000006117 | Ankylosis of distal interphalangeal joint |
| 483961000006114 | Ankylosis of metacarpophalangeal joint |
| 483981000006116 | Ankylosis of joint of multiple sites |
| 484061000006111 | Ankylosis of the first carpometacarpal joint |
| 484141000006111 | Ankylosis of the superior radioulnar joint |
| 491881000006113 | Arthralgia of lesser metatarsophalangeal joint |
| 491941000006112 | Arthralgia of sacroiliac joint |
| 492581000006117 | Arthropathy related to infection |
| 492631000006119 | Arthropathy associated with mycoses, of the pelvic region and thigh |
| 492971000006111 | Arthropathy associated with nonspecific urethritis |
| 497071000006114 | Atlantoaxial instability |
| 501991000006110 | Back pain |
| 531601000006119 | Closed fracture of second cervical vertebra without spinal cord injury |
| 560321000006116 | Closed fracture of vault of skull with intracranial injury, with no loss of consciousness |
| 562351000006114 | Closed fracture involving thorax wth lower back and pelvis and limbs |
| 563311000006118 | Closed subcondylar fracture of mandible |
| 563861000006112 | Closed fracture of patella |
| 564541000006113 | Closed fracture of head of humerus |
| 565331000006113 | Closed fracture dislocation of pelvis |
| 565481000006117 | Closed fracture dislocation, interphalangeal joint, single toe |
| 565791000006111 | Closed fracture subluxation, foot |
| 565921000006111 | Closed fractures involving multiple regions of one upper limb |
| 710161000006114 | Sprain of ligament of finger |
| 768881000006110 | Intracranial injury, without skull fracture |
| 792401000006116 | Fractures involing multiple regions of upper limb(s) with lower limb(s) |
| 792711000006112 | Fracture of fibula |
| 793141000006115 | Fracture of phalanx of toe |
| 793681000006116 | Fracture of transverse process of spine without spinal cord lesion |
| 253123011 | Finding of headache character |
| 299375010 | Inflammatory polyneuropathy |
| 309616014 | Septic arthritis of knee |
| 309711013 | Crystal arthropathy of elbow |
| 309713011 | Crystal arthropathy of wrist |
| 309748016 | Arthropathy associated with a haematological disorder |
| 101001000006111 | Sprain of thumb |
| 318266010 | Closed skull fracture with intracranial injury |
| 570931000006114 | Closed fracture of phalanx of finger |
| 793891000006119 | Fracture dislocation or subluxation foot |
| 320131015 | Sprain of upper extremity |
| 362631000006114 | [X]Additional musculoskeletal and connective tissue disease classification terms |
| 297404019 | Meninges disorder NEC |
| 310554019 | Ankylosis of other tarsal joint |
| 1222634011 | Closed fracture distal humerus, medial epicondyle |
| 890041000006110 | Traumatic arthritis NOS |
| 891611000006113 | Rheumatism NOS - hand |
| 891671000006116 | Myalgia/myositis - ankle/foot |
| 891751000006116 | Myalgia/myositis NOS |
| 891781000006112 | Neuralgia/neuritis - lower leg |
| 892721000006119 | Other musculoskelet/connectiv |
| 895361000006117 | #Spine NOS + cord lesion |
| 896191000006119 | Sprained shoulder |
| 896461000006119 | Other sprains |
| 933201000006114 | Benign coital headache |
| 990341000006117 | Sprained elbow |
| 990361000006118 | Sprain - wrist |
| 312509011 | [X]Infectious arthropathies |
| 391113018 | Back dislocation NOS |
| 402910017 | Other upper arm sprain |
| 299811000006115 | [D]Musculosc xray/scan abn NOS |
| 377501000006113 | [X]Dislocat/sprain/strain unsp joint/ligam leg, level unsp |
| 749211000006115 | Le Fort II fracture maxilla |
| 779411000006118 | Infective arthritis NOS, of lesser MTP joint |
| 779461000006115 | Infective arthritis NOS, of PIP joint of finger |
| 792811000006115 | Fracture of inferior maxilla, closed |
| 402851016 | Fracture of rib(s), sternum, larynx or trachea NOS |
| 736781000006114 | Localised osteoarthritis, unspecified, pelvic region/thigh |
| 309812014 | Rheumatoid bursitis |
| 312689019 | [X]Unspecified osteoporosis with pathological fracture |
| 892441000006117 | Path.fracture - hand |
| 892461000006118 | Path.fracture - upper arm |
| 892471000006113 | Path.fracture - shoulder |
| 491831000006112 | Arthralgia of distal radio-ulnar joint |
| 492131000006110 | Arthritis associated with other disease, DIP joint of finger |
| 492231000006116 | Arthritis associated with other disease, sacro-iliac joint |
| 531631000006110 | C3 vertebra open fracture without spinal cord lesion |
| 531651000006115 | C4 vertebra open fracture without spinal cord lesion |
| 538801000006111 | Carpal instability, D.I.S.I. |
| 565541000006112 | Closed fracture-dislocation, subtalar joint |
| 565631000006115 | Closed fracture-subluxation mid carpal |
| 565771000006110 | Closed fracture-subluxation, distal interphalangeal joint |
| 565851000006111 | Closed fracture-subluxation, metacarpophalangeal joint |
| 570841000006114 | Cls spinal fracture with unspec thoracic cord lesion,T1-6 |
| 405142017 | Unspecified polyarthropathy or polyarthritis |
| 455020013 | Closed fracture of bony thorax part unspecified |
| 455022017 | Fracture of other parts of bony thorax |
| 455023010 | Closed fracture of other parts of bony thorax |
| 2472449013 | Juvenile seronegative polyarthritis |
| 18121000006117 | Osteoarthritis NOS, of tibio-fibular joint |
| 25181000006118 | Other open fracture-subluxation |
| 29271000006114 | Other infect/parasit dis with arthropathy of other spec site |
| 31091000006116 | Osteoarthritis NOS, of lesser MTP joint |
| 34801000006115 | Other closed fracture-subluxation |
| 106831000006116 | Thoracic spinal hydromeningocele |
| 107341000006118 | Tarsal bone fracture |
| 130551000006116 | Sprain & strain of oth & unsp parts of lumb spine & pelv |
| 130601000006119 | Sprain dorsal radio-carpal ligament |
| 130701000006114 | Sprain finger, metacarpophalangeal joint, non specific |
| 130921000006119 | Sprain radio-scapho-lunate ligament |
| 131131000006113 | Sprain volar radio-carpal ligament non-specific |
| 131411000006114 | Sprain, knee joint, medial collateral |
| 168271000006117 | Rheumat. treatment change |
| 192771000006113 | Reactive arthropathy of sacro-iliac joint |
| 192821000006117 | Reactive arthropathy of tibio-fibular joint |
| 215551000000114 | Sero negative polyarthritis |
| 255651000006110 | Open fracture-subluxation, carpometacarpal joint |
| 255711000006118 | Open fracture-subluxation, IPJ, multiple toes |
| 255741000006119 | Open fracture-subluxation, metacarpophalangeal joint |
| 255781000006113 | Open fracture-subluxation, proximal interphalangeal joint |
| 256031000006118 | Open fracture-dislocation sterno-clavicular joint, anterior |
| 256171000006119 | Open fracture-dislocation, midtarsal joint |
| 256731000006116 | Open fracture-dislocation lunate (volar) |
| 265871000006111 | Oligoarticular osteoarthritis, unspecified, of ankle/foot |
| 318521000006110 | [Q] Open fracture grade 1 |
| 297162010 | Idiopathic peripheral autonomic neuropathy NOS |
| 297336013 | Common migraine NOS |
| 297352011 | Other forms of migraine |
| 309491010 | Arthropathy in Behcet's syndrome of other specified sites |
| 309544017 | Arthropathy associated with mycoses NOS |
| 309577018 | Reactive arthropathy of other tarsal joint |
| 309596019 | Infective arthritis NOS, of the shoulder region |
| 309602016 | Infective arthritis NOS, of the ankle and foot |
| 309619019 | Infective arthritis NOS, of subtalar joint |
| 309683012 | Gouty arthritis NOS |
| 309696013 | Other crystal arthropathies of other specified sites |
| 309698014 | Crystal arthropathy NOS |
| 309702013 | Crystal arthropathy NOS, of the forearm |
| 309705010 | Crystal arthropathy NOS, of the lower leg |
| 309732017 | Crystal arthropathy NOS, of other specified site |
| 309773016 | Arthritis associated with other disease, knee |
| 309775011 | Arthritis associated with other disease, ankle |
| 309777015 | Arthritis associated with other disease, talonavicular joint |
| 309824019 | Juvenile rheumatoid arthropathy unspecified |
| 309884016 | Localised, primary osteoarthritis of unspecified site |
| 309888018 | Localised, primary osteoarthritis of the upper arm |
| 309889014 | Localised, primary osteoarthritis of the forearm |
| 309941016 | Localised, secondary osteoarthritis NOS |
| 309965018 | Localised osteoarthritis, unspecified, of the ankle and foot |
| 309983014 | Osteoarthritis of more than one site, unspecified, NOS |
| 309987010 | Osteoarthritis NOS, of unspecified site |
| 310024016 | Osteoarthritis NOS, of knee |
| 310079012 | Traumatic arthropathy NOS |
| 310101013 | Climacteric arthritis of the lower leg |
| 310103011 | Climacteric arthritis of other specified site |
| 310149019 | Unspecified polyarthropathy of the shoulder region |
| 310191017 | Other specified arthropathy of multiple sites |
| 310216015 | Arthropathy NOS, of other specified site |
| 310518018 | Joint ankylosis of unspecified site |
| 310788010 | Palindromic rheumatism NOS |
| 311261013 | Thoracic and lumbosacral neuritis NOS |
| 312177015 | Other specified pathological fracture |
| 312960019 | Spinal cord anomalies NOS |
| 318285013 | Open fracture of skull NOS with intracranial injury |
| 318343012 | Fracture of skull NOS |
| 318575011 | Closed fracture of sacrum with other spinal cord injury |
| 318670013 | Open fracture acetabulum, double column unspecified |
| 318671012 | Other specified open fracture acetabulum |
| 318687010 | Other or multiple closed fracture of pelvis |
| 318730010 | Closed fracture of clavicle, unspecified part |
| 318758019 | Closed fracture of proximal humerus not otherwise specified |
| 318796018 | Open fracture of elbow, unspecified part |
| 318935013 | Open fracture of carpal bone NOS |
| 319053012 | Fracture of one or more phalanges of hand NOS |
| 319063016 | Ill-defined fractures of upper limb NOS |
| 319166010 | Closed fracture of distal femur not otherwise specified |
| 319177016 | Open fracture of distal femur not otherwise specified |
| 319194019 | Fracture of patella, NOS |
| 319219017 | Open fracture of tibia and fibula, proximal NOS |
| 319223013 | Closed fracture of tibia and fibula, shaft, NOS |
| 320305014 | Neck sprain NOS |
| 320557010 | Rib sprain unspecified |
| 325439014 | [X]Fractures of other parts of femur |
| 345456011 | Neuritis ulnar nerve |
| 391057017 | Muscle sprain NOS |
| 400182014 | Osteoarthritis NOS |
| 400183016 | Osteoarthritis NOS, of the upper arm |
| 402871013 | Open fracture proximal femur, other transcervical |
| 496410014 | ME - Myalgic encephalomyelitis |
| 496413011 | Myalgic encephalomyelitis |
| 1757331000000118 | Trigeminal autonomic cephalalgia |
| 1175081000000117 | Closed fracture of distal tibia and fibula |
| 875231000006113 | Therapeutic asp.-musculoskelet |
| 895301000006118 | #Sacrum/coccyx-no cord lesion |
| 896351000006111 | Sprain - Achilles tendon |
| 896501000006119 | Sprained symphisis pubis |
| 311711000000114 | Medication overuse headache |
| 545121000000114 | Fragility fracture |
| 2536030018 | Open fracture proximal femur, basicervical |
| 2548472016 | FH: maternal hip fracture before age 75 |
| 309483018 | Arthropathy in Behcet's syndrome of the shoulder region |
| 309490011 | Arthropathy in Behcet's syndrome of multiple sites |
| 309566012 | Reactive arthropathy of wrist |
| 309681014 | Gouty arthritis of multiple sites |
| 309836013 | Juvenile arthritis in ulcerative colitis |
| 309885015 | Localised, primary osteoarthritis of the shoulder region |
| 310060014 | Traumatic arthropathy of sternoclavicular joint |
| 310061013 | Traumatic arthropathy of acromioclavicular joint |
| 310100014 | Climacteric arthritis of the pelvic region and thigh |
| 310519014 | Joint ankylosis of the shoulder region |
| 310540010 | Elbow joint ankylosis |
| 310556017 | Ankylosis of toe joint |
| 310783018 | Palindromic rheumatism of the pelvic region and thigh |
| 310785013 | Palindromic rheumatism of the ankle and foot |
| 310822016 | Arthralgia of elbow |
| 310839019 | Arthralgia of ankle |
| 310842013 | Arthralgia of subtalar joint |
| 312011010 | Hypertrophic pulmonary osteoarthropathy |
| 312159013 | Drug-induced osteoporosis with pathological fracture |
| 312160015 | Idiopathic osteoporosis with pathological fracture |
| 312350016 | Chronic instability of knee |
| 312790015 | Cervical spinal hydromeningocele |
| 318135012 | Open fracture vault of skull with intracranial injury |
| 318208013 | Closed fracture of mandible, multiple sites |
| 318233011 | Open fracture maxilla |
| 318245019 | Fracture of alveolus, closed |
| 318341014 | Multiple fractures involving skull and facial bones |
| 318379016 | Closed fracture axis, spinous process |
| 318380018 | Closed fracture axis, transverse process |
| 318419013 | Open fracture atlas, comminuted |
| 318422010 | Open fracture axis, spinous process |
| 318424011 | Open fracture axis, posterior arch |
| 318454018 | Closed fracture lumbar vertebra |
| 318455017 | Closed fracture lumbar vertebra, burst |
| 318456016 | Closed fracture lumbar vertebra, wedge |
| 318473019 | Closed fracture sacrum |
| 318475014 | Closed vertical fracture of sacrum |
| 318476010 | Open fracture sacrum |
| 318478011 | Open vertical fracture of sacrum |
| 318492017 | Multiple fractures of lumbar spine and pelvis |
| 318546017 | Open spinal fracture with central thoracic cord lesion, T1-6 |
| 318587016 | Closed fracture of coccyx with spinal cord lesion |
| 318627013 | Closed fracture larynx and trachea |
| 318664019 | Open fracture acetabulum, anterior lip alone |
| 318665018 | Open fracture acetabulum, posterior lip alone |
| 318666017 | Open fracture acetabulum, anterior column |
| 318673010 | Closed fracture pubis |
| 318683014 | Open fracture pelvis, multiple pubic rami - stable |
| 318703017 | Open fracture pelvis, anterior superior iliac spine |
| 318705012 | Open fracture pelvis, iliac wing |
| 318744016 | Open fracture scapula, glenoid |
| 318745015 | Open fracture scapula, blade |
| 318784013 | Closed fracture distal humerus, lateral condyle |
| 318803014 | Open fracture distal humerus, capitellum |
| 318817017 | Closed fracture proximal radius, comminuted |
| 318928017 | Open fracture scaphoid, proximal pole |
| 318930015 | Open fracture scaphoid, waist, oblique |
| 318933018 | Open fracture carpal bones, multiple |
| 318938010 | Fracture of first metacarpal bone |
| 318949016 | Closed fracture finger metacarpal neck |
| 318951017 | Closed fracture finger metacarpal |
| 318966019 | Open fracture finger metacarpal base |
| 318971014 | Open fracture finger metacarpal, multiple |
| 318990018 | Closed fracture thumb proximal phalanx, base |
| 318996012 | Closed fracture thumb distal phalanx, shaft |
| 319010012 | Closed fracture finger middle phalanx, multiple |
| 319011011 | Closed fracture finger distal phalanx, base |
| 319028014 | Open fracture thumb distal phalanx, base |
| 319047012 | Open fracture finger distal phalanx, mallet |
| 319049010 | Open fracture of phalanx or phalanges, multiple sites |
| 319103016 | Open fracture proximal femur, midcervical section |
| 319165014 | Closed fracture distal femur, comminuted/intra-articular |
| 319182011 | Closed fracture patella, proximal pole |
| 319283010 | Closed fracture calcaneus, extra-articular |
| 319296011 | Closed fracture metatarsal base |
| 319298012 | Closed fracture metatarsal neck |
| 319311016 | Open fracture metatarsal head |
| 319750019 | Open spinal dislocation with anterior cervical cord lesion |
| 319751015 | Open spinal dislocation with central cervical cord lesion |
| 319867016 | Closed spinal subluxation with central thoracic cord lesion |
| 319961016 | Closed fracture dislocation of wrist |
| 320110018 | Sprains and strains of joints and adjacent muscles |
| 320118013 | Sprain, shoulder joint, anterior |
| 320120011 | Sprain, biceps tendon |
| 320168017 | Sprain dorsal intercarpal ligament |
| 320211014 | Sprain wrist extensors |
| 320213012 | Sprain tendon of thumb |
| 320249013 | Sprain of superior tibiofibular ligament |
| 320263019 | Distal tibiofibular sprain |
| 320303019 | Atlanto-occipital joint sprain |
| 320307018 | Lumbar sprain |
| 320308011 | Lumbar back sprain |
| 324031014 | Multiple open wounds of abdomen, lower back and pelvis |
| 325251014 | Fractures involving multiple regions of one lower limb |
| 345361012 | Chronic tension-type headache |
| 297164011 | Autonomic neuropathy due to amyloid |
| 297354012 | Complicated migraine |
| 297470010 | Mononeuritis of upper limb and mononeuritis multiplex |
| 297641010 | Myopathy due to rheumatoid arthritis |
| 253103010 | Headache site |
| 28671011 | Allergic arthritis |
| 34376013 | Fracture of upper end of tibia |
| 41818011 | Open fracture of calcaneus |
| 48625014 | Closed Monteggia's fracture |
| 50336019 | Morton's neuralgia |
| 55628018 | Psoriatic arthritis |
| 56659013 | Lyme arthritis |
| 59922014 | Chronic arthritis |
| 63055014 | Migraine |
| 70659018 | Polyneuropathy |
| 80383012 | Septic arthritis |
| 94456015 | Polyneuritis cranialis |
| 116082011 | Rheumatoid arthritis |
| 131972016 | Closed fracture of three ribs |
| 132104018 | Closed fracture of talus |
| 492363016 | Open fracture of radius and ulna, shaft |
| 500766011 | Neuropathic arthropathy |
| 504520011 | Haemophilic arthropathy |
| 1219615011 | Arthropathy associated with mycoses |
| 1222720019 | Open fracture shaft of tibia |
| 1227400011 | Closed fracture distal femur, supracondylar |
| 1228300012 | Closed fracture trapezoid |
| 1228677017 | Pyogenic arthritis of the shoulder region |
| 1229173011 | Hip pyogenic arthritis |
| 1229766011 | Open fracture finger distal phalanx |
| 1229933018 | Closed fracture of the scaphoid |
| 1230102017 | Open fracture capitate |
| 1230277019 | Open fracture rib |
| 1231346013 | Open fracture ankle, medial malleolus |
| 1232322015 | Open fracture distal humerus, supracondylar |
| 1233503017 | Open fracture clavicle, lateral end |
| 1234611012 | Closed fracture nasal bone |
| 1235429018 | Open fracture pisiform |
| 1235549012 | Closed fracture of humerus, shaft |
| 1495084019 | Fracture of lower jaw, open |
| 1495085018 | Fracture of mandible, open |
| 1495384010 | Open fracture radius and ulna, proximal |
| 1779323014 | Rheumatoid vasculitis |
| 359268016 | Elbow pyogenic arthritis |
| 359292012 | Seronegative rheumatoid arthritis |
| 359402016 | Osteoarthritis of spinal facet joint |
| 372799017 | Back stiffness |
| 391313012 | Fracture of mandible |
| 391349013 | Fracture of upper end of radius |
| 391384012 | Hip fracture |
| 391401018 | Closed fracture of tibial tuberosity |
| 391420016 | Metatarsal bone fracture |
| 391426010 | Multiple fractures of sternum |
| 400160019 | Pyogenic arthritis of the lower leg |
| 400161015 | Pyogenic arthritis of the ankle and foot |
| 400251019 | Arthralgia of the ankle and foot |
| 402833017 | Closed fracture of cervical spine |
| 402835012 | Closed fracture axis |
| 402856014 | Open fracture of the distal humerus |
| 407069017 | C/O - a headache |
| 415888015 | Chronic low back pain |
| 455447011 | Open multiple fractures of clavicle, scapula and humerus |
| 456416014 | Late effect of fracture of lumbar vertebra |
| 457122015 | Localised, primary osteoarthritis of elbow |
| 459312014 | Type II diabetes mellitus with neuropathic arthropathy |
| 459313016 | Type 2 diabetes mellitus with neuropathic arthropathy |
| 309505013 | Arthropathy associated with bacterial disease |
| 309560018 | Post-infective arthritis |
| 311225015 | Back problem |
| 311260014 | Lumbosacral neuritis |
| 318246018 | Closed fracture of orbit |
| 318721019 | Closed fracture of bones of trunk |
| 318777017 | Open fracture of humerus |
| 318877014 | Open fracture of distal end of ulna |
| 318901010 | Closed fracture of radius AND ulna |
| 318914011 | Open fracture of radius AND ulna |
| 319160016 | Closed fracture of femoral condyle of femur |
| 319246010 | Open fracture of tibia |
| 319272018 | Open fracture of ankle |
| 359171019 | Arthritis of wrist |
| 359384017 | Osteoarthritis of foot joint |
| 359385016 | Osteoarthritis of ankle |
| 391075016 | Tendon strain |
| 391329016 | Closed fracture of atlas without spinal cord injury |
| 391372010 | Rib fracture |
| 391425014 | Fracture of lower jaw |
| 18061000006112 | Inflammation of sacroiliac joint |
| 31031000006115 | Osteoarthritis of distal interphalangeal joint |
| 40881000006116 | Osteoporosis with pathological fracture of lumbar vertebrae |
| 43141000006110 | Open spinal fracture with complete thoracic cord lesion, T1-6 |
| 44351000006113 | Open fracture thumb metacarpal base, intra-articular, Bennett |
| 60781000006118 | Villonodular synovitis of sacroiliac joint |
| 92641000006116 | Traumatic arthropathy of distal interphalangeal joint of finger |
| 92661000006117 | Traumatic arthropathy of interphalangeal joint of toe |
| 92721000006115 | Traumatic arthropathy of proximal interphalangeal joint of finger |
| 108781000006112 | Ankylosis of temporomandibular joint |
| 131031000006112 | Sprain thumb, metacarpophalangeal joint, ulnar collateral ligament |
| 131181000006114 | Sprain of ligament of acromioclavicular joint |
| 131381000006111 | Strain of infraspinatus tendon |
| 131491000006116 | Strain of quadriceps tendon |
| 131521000006119 | Sprain of shoulder joint |
| 131581000006115 | Sprain, tarsometatarsal joint |
| 168281000006119 | Rheumatology disorder - joints affected |
| 192741000006117 | Reactive arthropathy of metacarpophalangeal joint |
| 217741000000112 | Sinus headache |
| 221521000000114 | Osteoarthritis of knee |
| 235151000006117 | Migrainous neuralgia |
| 253061000006117 | Closed fractures of multiple bones of lower limb |
| 256061000006110 | Open fracture dislocation, ankle joint |
| 256141000006110 | Open fracture dislocation, knee joint |
| 256651000006118 | Open fracture of zygoma |
| 257801000006118 | Open fracture proximal tibia |
| 264131000006117 | Open fracture of base of skull with intracranial injury, with more than 24 hours loss of consciousness and return to pre-existing conscious level |
| 491931000006119 | Arthralgia of proximal interphalangeal joint of finger |
| 492541000006111 | Arthropathy associated with another disorder |
| 544821000006112 | Cervicothoracic instability |
| 559981000006117 | Closed fracture of base of skull with intracranial injury, with less than 1 hour loss of consciousness |
| 560271000006112 | Closed fracture of vault of skull with intracranial injury, with more than 24 hours loss of consciousness without return to pre-existing conscious level |
| 561651000006113 | Closed fracture of base of skull with intracranial injury |
| 564861000006114 | Closed fracture of shaft of tibia |
| 565491000006119 | Closed fracture dislocation, knee joint |
| 567841000006116 | Closed spinal subluxation with posterior thoracic cord lesion |
| 570581000006118 | Closed fracture thumb metacarpal base, intra-articular, Bennett |
| 616971000006114 | Diabetic Charcot's arthropathy |
| 695161000006110 | Fracture of multiple ribs |
| 736911000006116 | Localised, primary osteoarthritis of the pelvic region and thigh |
| 750591000006119 | Late effect of fracture of spine and trunk without mention of cord lesion |
| 793161000006116 | Fracture of phalanx of hand |
| 793931000006111 | Fracture dislocation or subluxation shoulder |
| 815501000006110 | Arthritis of hand |
| 793881000006117 | Fracture dislocation of elbow joint |
| 319203012 | Closed fracture spine, tibia |
| 883141000006119 | Mononeuritis - upper limb |
| 883151000006117 | Other mononeuritis -lower limb |
| 891621000006117 | Rheumatism NOS - wrist |
| 891651000006114 | Rheumatism NOS - multiple |
| 891661000006111 | Rheumatism/fibrositis NOS |
| 891741000006118 | Myalgia/myositis - multiple |
| 891791000006110 | Neuralgia/neurit.-pelvis/thigh |
| 891801000006111 | Neuralgia/neuritis - hand |
| 892561000006117 | Fracture malunion - shoulder |
| 896391000006117 | Sprain - foot NOS |
| 933131000006112 | Back injury |
| 989361000006114 | Osteoarthritis - hip joint |
| 990101000006117 | Sprained thigh - upper leg |
| 409930017 | Thoracic back sprain |
| 317103014 | [D]Nervous and musculoskeletal symptoms |
| 309727018 | Crystal arthropathy NOS, of other tarsal joint |
| 309805017 | Rheumatoid arthritis of other tarsal joint |
| 312523013 | [X]Other juvenile arthritis |
| 318247010 | Fracture of other facial bones, closed, NOS |
| 312728019 | [X]Postprocedural musculoskeletal disorder, unspecified |
| 320111019 | Sprain of shoulder and upper arm |
| 492141000006117 | Arthritis associated with other disease, dist rad-uln joint |
| 493001000006113 | Arthropathy in Whipple's disease |
| 493141000006114 | Arthropathy with other bacterial disease, of ankle and foot |
| 538841000006113 | Carpal instability, V.I.S.I. |
| 556881000000111 | Minimal trauma fracture due to unspecified osteoporosis |
| 565381000006114 | Closed fracture-dislocation radiocarpal joint |
| 565581000006118 | Closed fracture-subluxation digit, unspecified |
| 565641000006113 | Closed fracture-subluxation multiple digits |
| 565681000006119 | Closed fracture-subluxation other carpal |
| 565741000006119 | Closed fracture-subluxation superior radio-ulnar joint |
| 567621000006112 | Closed spinal dislocation with thoracic cord lesion, unspec |
| 567831000006114 | Closed spinal subluxation with lumbar cord lesion, unspec |
| 569521000006119 | Closed volar Barton's fracture-dislocation |
| 570641000006113 | Cls spinal # with incomplete cervical cord lesion, C5-7 NOS |
| 593591000006116 | Convalesc. after fracture Rx |
| 615711000006112 | Detrusor instability |
| 675631000006115 | Neuropathic arthritis |
| 693501000006115 | Musculoskeletal pain - joints |
| 698001000006119 | Mononeuritis lower limb |
| 718001000006112 | Marie - Strumpell spondylitis |
| 779351000006112 | Infective arthritis NOS, of DIP joint of finger |
| 793761000006117 | Fracture of upper jaw, open |
| 142901000006114 | Sinus headache |
| 159451000006113 | Rvsn open reduc spinal fracture+other external stabilisation |
| 162051000006115 | Rheumatoid arthritis of distal radio-ulnar joint |
| 192681000006117 | Reactive arthropathy of distal radio-ulnar joint |
| 255691000006116 | Open fracture-subluxation, hip joint |
| 255791000006111 | Open fracture-subluxation, subtalar joint |
| 255961000006117 | Open fracture-dislocation of sternum |
| 255991000006113 | Open fracture-dislocation peri-lunate trans-scaphoid |
| 256001000006114 | Open fracture-dislocation radiocarpal joint |
| 256021000006116 | Open fracture-dislocation shoulder joint |
| 256041000006111 | Open fracture-dislocation sterno-clavicular joint, posterior |
| 256101000006113 | Open fracture-dislocation, hip joint |
| 256111000006111 | Open fracture-dislocation, interphalangeal joint thumb |
| 256151000006112 | Open fracture-dislocation, metacarpophalangeal joint |
| 256251000006116 | Open fracture-subluxation elbow |
| 256351000006110 | Open fracture-subluxation other carpal |
| 257831000006114 | Open fracture of the radius and ulna |
| 376211000006111 | [X]Crystal arthropathy in other metabolic disorders CE |
| 310186018 | Other specified arthropathy of the hand |
| 310192012 | Other specified arthropathy NOS |
| 310781016 | Palindromic rheumatism of the forearm |
| 310786014 | Palindromic rheumatism of other specified site |
| 312178013 | Pathological fracture NOS |
| 312789012 | Spinal hydromeningocele, unspecified |
| 314546014 | Congenital musculoskeletal deformity NOS |
| 315764010 | [X]Other specified congenital musculoskeletal deformities |
| 317576014 | [D]Abdominal migraine |
| 318396015 | Open fracture of unspecified cervical vertebra |
| 318527018 | Open fracture of cervical spine with spinal cord lesion NOS |
| 318580019 | Closed fracture of sacrum with spinal cord lesion NOS |
| 318586013 | Open fracture of sacrum with spinal cord lesion NOS |
| 318591014 | Closed fracture of coccyx with other spinal cord injury |
| 318624018 | Open fracture of rib(s) NOS |
| 318663013 | Closed fracture acetabulum NOS |
| 318723016 | Fracture of ill-defined bone of trunk NOS |
| 318741012 | Closed fracture of scapula NOS |
| 318820013 | Closed fracture of proximal forearm not otherwise specified |
| 318821012 | Open fracture of proximal forearm, unspecified |
| 318868019 | Open fracture of forearm, lower end, unspecified |
| 318925019 | Closed fracture of carpal bone NOS |
| 318926018 | Open fracture of carpal bone, unspecified |
| 318986015 | Closed fracture of phalanx or phalanges, unspecified |
| 319314012 | Open fracture of tarsal and metatarsal bones NOS |
| 320290011 | Other specified sacroiliac sprains |
| 320291010 | Sacroiliac sprain NOS |
| 320554015 | Thyroid region sprain NOS |
| 325324014 | [X]Fracture of other parts of neck |
| 399418016 | Toxic or inflammatory neuropathy NOS |
| 400315011 | Neuralgia, neuritis or radiculitis NOS |
| 402830019 | Multiple fractures involving skull or face with other bones |
| 455454017 | Open fracture of lower limb, level unspecified |
| 460304013 | [V]Personal history of other musculoskeletal disorders |
| 18191000006115 | Osteoarthritis spine |
| 31071000006117 | Osteoarthritis NOS, of IP joint of toe |
| 41261000006115 | Osteoarthritis and allied disorders |
| 41281000006113 | Osteoarthritis NOS |
| 49851000006119 | Open volar Barton fracture-dislocation |
| 52291000006115 | Open Smith's fracture |
| 82841000006116 | Tuberculous arthritis |
| 84391000006111 | Type 1 diabetes mellitus with neuropathic arthropathy |
| 130691000006114 | Sprain finger, distal interphalangeal joint, non specific |
| 130731000006118 | Sprain luno-triquetral ligament |
| 130881000006110 | Sprain proximal radiocarpal ligament non-specific |
| 276496014 | Primary other external stabilisation of spinal fracture |
| 297429013 | Other trigeminal nerve disorder |
| 297574016 | Other toxic or inflammatory neuropathy |
| 309540014 | Arthropathy associated with mycoses, of the lower leg |
| 309543011 | Arthropathy associated with mycoses, of other specified site |
| 309550010 | Helminthiasis with arthropathy of the upper arm |
| 309603014 | Infective arthritis NOS, of shoulder |
| 309679012 | Gouty arthritis of the lower leg |
| 309776012 | Arthritis associated with other disease, subtalar joint |
| 309850015 | Inflammatory polyarthropathy NOS |
| 309979014 | Oligoarticular osteoarthritis, unspecified, of lower leg |
| 310020013 | Osteoarthritis NOS, of hip |
| 310057019 | Traumatic arthropathy of other specified site |
| 310075018 | Traumatic arthropathy of other tarsal joint |
| 310095012 | Climacteric arthritis of unspecified site |
| 310097016 | Climacteric arthritis of the upper arm |
| 310153017 | Unspecified polyarthropathy of the pelvic region and thigh |
| 310155012 | Unspecified polyarthropathy of the ankle and foot |
| 310168016 | Unspecified monoarthritis of the upper arm |
| 310174016 | Unspecified monoarthritis of other specified site |
| 310175015 | Unspecified monoarthritis NOS |
| 310185019 | Other specified arthropathy of the forearm |
| 158497012 | Ulnar neuritis |
| 1175111000000113 | Open fracture of distal tibia and fibula |
| 990121000006110 | Sprain - lateral knee ligament |
| 990271000006116 | Fracture malunion - lower leg |
| 990371000006113 | Sprain - hand |
| 990381000006111 | Sprained finger/thumb |
| 990851000006113 | Fracture malunion - NOS |
| 889731000006111 | Rheumatoid arthritis NOS |
| 889911000006119 | Osteoarthritis - hip joint |
| 889931000006113 | Osteoarthritis - ankle/foot |
| 895911000006116 | Multiple fractures |
| 896201000006116 | Sprain - fore arm |
| 896211000006118 | Sprained elbow |
| 896271000006110 | Sprained hip |
| 896371000006118 | Sprained foot |
| 896471000006114 | Sprained jaw |
| 612281000006119 | Delayed union of fracture |
| 2533505017 | Fracture of tibia AND fibula |
| 2548331017 | Inflammatory polyarthropathy |
| 2674612015 | History of headache |
| 110091000006114 | Test request : R.A. Screen |
| 490703010 | Sciatic nerve lesion |
| 493961016 | Familial amyloid polyneuropathy type II |
| 496729019 | Closed fracture of the radius and ulna |
| 497076011 | Knee sprain |
| 510090013 | Multiple fractures of metacarpal bones |
| 1222629012 | Toe fracture |
| 1223045018 | Late effect of fracture of arm |
| 1229552016 | Arthropathy associated with endocrine and metabolic disorder |
| 1229897016 | Closed fracture of the proximal humerus |
| 1230026018 | Closed fracture pisiform |
| 1231252012 | Closed fracture of the ulnar shaft |
| 1232867019 | Closed fracture ankle, trimalleolar |
| 1233163011 | Open fracture radial head |
| 1234127016 | Open fracture metatarsal |
| 1234550018 | Congenital spinal cord anomaly |
| 1235278014 | Lumbosacral ankylosis |
| 1235556018 | Open fracture ankle, trimalleolar |
| 1488690018 | FH: Ankylosing spondylitis |
| 1777688018 | Muscular headache |
| 2164085015 | Detrusor instability |
| 359266017 | Knee pyogenic arthritis |
| 359273010 | Arthropathy due to parasitic infection |
| 391056014 | Rectus muscle sprain |
| 391098012 | Hamstring sprain |
| 391402013 | Open fracture of tibial tuberosity |
| 391403015 | Open fracture of tibial condyles |
| 391421017 | Fracture of metatarsal bone |
| 397716013 | FH: Musculoskeletal disease |
| 402831015 | Fracture of vertebra without spinal cord lesion |
| 402836013 | Closed fracture of third cervical vertebra |
| 402854012 | Closed fracture of the distal humerus |
| 411307011 | Closed fracture of femur, greater trochanter |
| 411549017 | H/O: arthritis |
| 416146018 | Mechanical low back pain |
| 456414012 | Late effect of fracture of cervical vertebra |
| 297542011 | Polyneuropathy in disseminated lupus erythematosus |
| 297544012 | Polyneuropathy in rheumatoid arthritis |
| 297557016 | Polyneuropathy in herpes zoster |
| 297563013 | Polyneuropathy in pellagra |
| 309486014 | Arthropathy in Behcet's syndrome of the hand |
| 309538016 | Arthropathy associated with mycoses, of the hand |
| 309541013 | Arthropathy associated with mycoses, of the ankle and foot |
| 309542018 | Arthropathy associated with mycoses, of multiple sites |
| 309561019 | Reactive arthropathy of shoulder |
| 309564010 | Reactive arthropathy of elbow |
| 309677014 | Gouty arthritis of the hand |
| 309678016 | Gouty arthritis of the pelvic region and thigh |
| 309791014 | Rheumatoid arthritis of acromioclavicular joint |
| 309804018 | Rheumatoid arthritis of talonavicular joint |
| 310058012 | Traumatic arthropathy of multiple sites |
| 310074019 | Traumatic arthropathy of talonavicular joint |
| 310435017 | Carpal instability |
| 310539013 | Ankylosis of the elbow joint |
| 310553013 | Ankylosis of the subtalar joint |
| 310779018 | Palindromic rheumatism of the shoulder region |
| 310802015 | Arthralgia of the hand |
| 310819018 | Arthralgia of acromioclavicular joint |
| 310846011 | Arthralgia of talonavicular joint |
| 311293017 | Lumbar spine instability |
| 312788016 | Spinal hydromeningocele |
| 312799019 | Spinal meningocele |
| 312803018 | Lumbar spinal meningocele |
| 312808010 | Lumbar meningomyelocele |
| 318126017 | Open fracture vault of skull without intracranial injury |
| 318238019 | Fracture of orbital floor |
| 318384010 | Closed fracture cervical vertebra, wedge |
| 318437014 | Closed fracture thoracic vertebra, wedge |
| 318439012 | Closed fracture thoracic vertebra, spinous process |
| 318451014 | Open fracture thoracic vertebra, transverse process |
| 318593012 | Open fracture of coccyx with spinal cord lesion |
| 318678018 | Closed fracture pelvis, multiple pubic rami - unstable |
| 318684015 | Open fracture pelvis, multiple pubic rami - unstable |
| 318725011 | Multiple fractures of thoracic spine |
| 318738015 | Closed fracture scapula, blade |
| 318740013 | Closed fracture scapula, neck |
| 318743010 | Open fracture scapula, coracoid |
| 318816014 | Closed fracture of proximal ulna, comminuted |
| 318850017 | Closed Galeazzi fracture |
| 318858012 | Closed fracture distal radius, intra-articular, die-punch |
| 318880010 | Smith's fracture - open |
| 318915012 | Fracture of radius and ulna, NOS |
| 318931016 | Open fracture scaphoid, waist, comminuted |
| 318952012 | Closed fracture finger metacarpal, multiple |
| 318985016 | Closed fracture of one or more phalanges of hand |
| 318994010 | Closed fracture thumb distal phalanx |
| 319003018 | Closed fracture finger proximal phalanx, head |
| 319005013 | Closed fracture finger middle phalanx |
| 319007017 | Closed fracture finger middle phalanx, shaft |
| 319008010 | Closed fracture finger middle phalanx, neck |
| 319030011 | Open fracture thumb distal phalanx, tuft |
| 319031010 | Open fracture thumb distal phalanx, mallet |
| 319044017 | Open fracture finger distal phalanx, base |
| 319045016 | Open fracture finger distal phalanx, shaft |
| 319046015 | Open fracture finger distal phalanx, tuft |
| 319084014 | Closed fracture proximal femur, transcervical |
| 319100018 | Open fracture proximal femur, transcervical |
| 319123015 | Closed fracture proximal femur, intertrochanteric, two part |
| 319124014 | Closed fracture proximal femur, subtrochanteric |
| 319161017 | Closed fracture of femur, lower epiphysis |
| 319180015 | Fracture of femur, NOS |
| 319201014 | Closed fracture proximal tibia, lateral condyle (plateau) |
| 319213016 | Open fracture proximal tibia, lateral condyle (plateau) |
| 319217015 | Open fracture fibula, head |
| 319230019 | Open fracture distal tibia |
| 319254012 | Open fracture ankle, lateral malleolus, low |
| 319261011 | Closed fracture ankle, bimalleolar, low fibular fracture |
| 319328018 | Fracture of great toe |
| 319714014 | Closed spinal dislocation with complete cervical cord lesion |
| 319715010 | Closed spinal dislocation with anterior cervical cord lesion |
| 319866013 | Closed spinal subluxation with anterior thoracic cord lesion |
| 320135012 | Sprain, elbow joint, radial collateral ligament |
| 320151016 | Sprain radial collateral ligament |
| 320261017 | Deltoid ligament ankle sprain |
| 320288010 | Sprain, sacrotuberous ligament |
| 320301017 | Cervical anterior longitudinal ligament sprain |
| 320573016 | Complete tear, sacroiliac ligament |
| 325244017 | Fractures involving head with neck |
| 250085017 | FH: Rheumatoid arthritis |
| 250086016 | FH: Osteoarthritis |
| 253117018 | Headache character |
| 255925015 | O/E-hands-rheumatoid spindling |
| 574014 | Fracture of upper end of humerus |
| 7595017 | Migraine with aura |
| 13705013 | Fracture of ilium |
| 18666015 | Irritable bowel syndrome |
| 30628014 | Congenital deformity of sacroiliac joint |
| 32869011 | Arthritis mutilans |
| 36906016 | Open fracture of three ribs |
| 55358019 | Closed fracture of clavicle |
| 56330019 | Fracture of rib |
| 57701019 | Open Monteggia's fracture |
| 60285012 | Fracture of pubis |
| 80869018 | Muscle strain |
| 93294013 | Migraine without aura |
| 96634011 | Open fracture of four ribs |
| 108062014 | Hereditary peripheral neuropathy |
| 114663015 | Open fracture of seven ribs |
| 129259017 | Climacteric arthritis |
| 135862013 | Sprain of medial collateral ligament of knee |
| 138588018 | Cervical spinal stenosis |
| 7596016 | Migraine with typical aura |
| 14103017 | Alcohol-induced polyneuropathy |
| 45939011 | Fracture of facial bone |
| 78112010 | Fracture of bone of lower limb |
| 299339013 | Headache disorder |
| 312617013 | Fracture of vertebral column |
| 318701015 | Multiple open fractures of pelvis with disruption of pelvic circle |
| 318834013 | Open fracture of shaft of radius AND ulna |
| 319141011 | Open fracture of femur |
| 325323015 | Fracture of cervical spine |
| 325342016 | Fracture of bones of trunk |
| 359172014 | Arthritis of elbow |
| 359383011 | Osteoarthritis of toe joint |
| 31101000006110 | Osteoarthritis of metacarpophalangeal joint |
| 44361000006110 | Open fracture thumb metacarpal base, intra-articular, Rolando |
| 44441000006115 | Open spinal fracture with anterior thoracic cord lesion, T7-12 |
| 106931000006114 | Thoracolumbar ankylosis |
| 108621000006117 | Fracture of temporal bone |
| 131021000006114 | Sprain thumb, metacarpophalangeal joint, radial collateral ligament |
| 159841000006114 | Sacroiliac sprain |
| 164851000006115 | Strain of rotator cuff of shoulder |
| 168761000006118 | Rheumatoid arthritis of 1st metatarsophalangeal joint |
| 192641000006111 | Reactive arthropathy of 1st metatarsophalangeal joint |
| 192761000006118 | Reactive arthropathy of proximal interphalangeal joint of finger |
| 221671000006119 | Polyarthropathy |
| 258091000006115 | Open fracture of phalanx of toe |
| 263781000006117 | Open fracture of vault of skull with intracranial injury, with no loss of consciousness |
| 264161000006114 | Open fracture of base of skull with intracranial injury, with no loss of consciousness |
| 387591000006110 | Fracture at wrist and/or hand level |
| 427481000006111 | Strain of tendon of head and neck |
| 483811000006116 | Pyogenic arthritis of ankle |
| 484101000006114 | Ankylosis of the inferior radioulnar joint |
| 531571000006114 | Closed fracture of first cervical vertebra without spinal cord injury |
| 559801000006115 | Closed fractures involving multiple regions of both lower limbs |
| 559991000006119 | Closed fracture of base of skull with intracranial injury, with more than 24 hours loss of consciousness and return to pre-existing conscious level |
| 560001000006112 | Closed fracture of base of skull with intracranial injury, with 1-24 hours loss of consciousness |
| 560021000006119 | Closed fracture of base of skull with intracranial injury, with no loss of consciousness |
| 560301000006114 | Closed fracture of vault of skull with intracranial injury, with 1-24 hours loss of consciousness |
| 565231000006117 | Closed fracture dislocation digit |
| 565251000006112 | Closed fracture dislocation distal radioulnar joint |
| 565281000006116 | Closed fracture dislocation foot |
| 565451000006113 | Closed fracture dislocation, hip joint |
| 565521000006117 | Closed fracture dislocation, patellofemoral joint |
| 570681000006119 | Closed spinal fracture with anterior cervical cord lesion, C5-7 |
| 570821000006119 | Closed fracture of C5-C7 level with spinal cord injury |
| 570851000006111 | Closed spinal fracture with anterior thoracic cord lesion, T1-6 |
| 601231000006112 | Crushing injuries of thorax with abdomen, lower back and pelvis with limb(s) |
| 623381000006114 | Dislocations, sprains and strains involving multiple regions of upper limb(s) |
| 649231000006118 | Fracture of ethmoid sinus |
| 736171000006110 | Localised, secondary osteoarthritis of the pelvic region and thigh |
| 749201000006118 | LFI - Le Fort I fracture of maxilla |
| 793871000006115 | Fracture dislocation or subluxation ankle |
| 793911000006117 | Fracture dislocation or subluxation knee |
| 794941000006115 | Fracture of frontal sinus |
| 819861000006110 | Helminthiasis with arthropathy of the pelvic region and thigh |
| 1660221000006115 | Benign coital headache |
| 309788014 | Rheumatoid arthritis of spine |
| 318893017 | Open fracture of lower end of forearm |
| 318913017 | Open fracture of ulna |
| 319140012 | Closed fracture of femur |
| 391111016 | Fracture dislocation of joint |
| 319171015 | Open fracture of femoral condyle of femur |
| 363811000006111 | Infective polyarthritis |
| 570541000006112 | Closed fracture of intracapsular section of femur |
| 276488016 | Primary collar stabilisation of cervical spine fracture |
| 312224018 | Fracture malunion |
| 565721000006114 | Closed fracture subluxation shoulder joint |
| 531581000006112 | Open fracture of first cervical vertebra without mention of spinal cord lesion |
| 557951000006118 | Closed fractures involving multiple regions upper with lower limb |
| 562361000006111 | Closed fracture of larynx |
| 564391000006113 | Closed fracture of base of neck of femur |
| 565431000006118 | Closed fracture dislocation, carpometacarpal joint |
| 565561000006111 | Closed fracture subluxation acromioclavicular joint |
| 565571000006116 | Closed fracture subluxation digit |
| 566791000006117 | Closed multiple fractures involving both upper limbs, and upper limb with rib(s) and sternum |
| 570691000006116 | Closed spinal fracture with anterior thoracic cord lesion,T7-12 |
| 570741000006119 | Closed spinal fracture with complete cervical cord lesion, C1-4 |
| 570861000006113 | Closed spinal fracture with complete thoracic cord lesion, T1-6 |
| 726781000006111 | Hypertrophic nonunion of fracture |
| 789621000006119 | Hyperuricaemia without signs of inflammatory arthritis and tophaceous disease |
| 792411000006118 | Fractures involving thorax with lower back and pelvis with limb(s) |
| 793411000006114 | Fracture of scaphoid bone of wrist |
| 1668051000000112 | Open fracture of tibial plateau |
| 1786081000006116 | Benign cough headache |
| 1927491000006111 | Irritable bowel syndrome characterised by alternating bowel habit |
| 402863014 | Closed fracture of radius |
| 47011000006118 | Open wounds involving thorax with abdomen, lower back and pelvis |
| 54641000006115 | Osteoarthritis of wrist |
| 92681000006110 | Traumatic arthropathy of metacarpophalangeal joint |
| 130621000006112 | Sprain finger, distal interphalangeal joint, radial collateral ligament |
| 131011000006118 | Sprain thumb, interphalangeal joint, ulnar collateral ligament |
| 134471000006115 | Fracture of sphenoid bone |
| 149911000006117 | Seropositive rheumatoid arthritis |
| 162141000006118 | Rheumatoid arthritis of sacroiliac joint |
| 219271000000117 | Seronegative arthritis |
| 221491000000111 | Osteoarthritis of elbow |
| 265381000006115 | Open fractures involving multiple regions of both upper limbs |
| 312321000000110 | Closed fracture of base of fifth metatarsal bone |
| 491821000006114 | Arthralgia of distal interphalangeal joint of finger |
| 492111000006116 | Arthritis of acromioclavicular joint |
| 299347013 | Drug induced headache |
| 299365012 | Inflammatory neuropathy |
| 309942011 | Localised osteoarthritis |
| 310011016 | Osteoarthritis of shoulder |
| 310205010 | Arthropathy of joint of hand |
| 311705018 | Neuralgia |
| 312495012 | Disorder of musculoskeletal system |
| 317435015 | Musculoskeletal chest pain |
| 318750014 | Closed fracture of upper end of humerus |
| 318864017 | Closed fracture of lower end of forearm |
| 325487016 | Dislocations, sprains and strains involving multiple body regions |
| 359848019 | Ankylosis of spine |
| 400285018 | Thoracic neuritis |
| 493251000006115 | Arthropathy with other viral disease, of forearm |
| 493331000006119 | Arthropathy with other viral disease, of upper arm |
| 319215011 | Open fracture spine, tibia |
| 889721000006113 | Post-viral arthropathy |
| 890971000006115 | Back disorders - other |
| 891731000006111 | Myalgia/myositis - shoulder |
| 891771000006114 | Neuralgia/neuritis -ankle/foot |
| 891811000006114 | Neuralgia/neuritis - fore arm |
| 892581000006110 | Malunion/nonunion of fracture |
| 894961000006119 | Musculoskeletal x-ray abn. [D] |
| 895571000006117 | Fracture of distal end of ulna |
| 896331000006116 | Sprained knee NOS |
| 896361000006113 | Sprain - ankle NOS |
| 931891000006117 | Headache disorder |
| 932681000006110 | Other peripheral neuropathy |
| 933101000006116 | Neck injury |
| 989371000006119 | Osteoarthritis - knee joint |
| 990091000006111 | Sprained hip |
| 990111000006119 | Sprained knee |
| 473482013 | Sacroiliac sprain |
| 318806018 | Open fracture of distal humerus, not otherwise specified |
| 309464013 | Pyogenic arthritis NOS |
| 318144013 | Fracture of vault of skull NOS |
| 603591000006113 | Crystal arthropathy NOS, of PIP joint of finger |
| 297531019 | Hereditary or idiopathic peripheral neuropathy NOS |
| 455508011 | [X]Closed multiple fractures unspecified |
| 892451000006115 | Path.fracture - fore arm |
| 310148010 | Unspecified polyarthropathy of unspecified site |
| 320316019 | Coccyx sprain |
| 461577011 | [Q] Lateral spinal stenosis |
| 312528016 | [X]Other specific arthropathies, not elsewhere classified |
| 318599011 | Closed fracture of spine with spinal cord lesion unspecified |
| 461006012 | [V]Other specified psychological or physical strain |
| 461440017 | [X]Other physical and mental strain related to work |
| 130561000006119 | Sprain & strain of oth & unspecif parts of should girdle |
| 363821000006115 | [X]Arthropathies in other specified diseases CE |
| 405781000006116 | [X]Oth/unspecif sympt & signs involv nerv/musculosk systems |
| 427431000006110 | [X]Sprain & strain of oth & unspecif parts of should girdle |
| 424981000006114 | [X]Rheumatoid arthritis+involvement/other organs or systems |
| 492171000006113 | Arthritis associated with other disease, IP joint of toe |
| 492201000006112 | Arthritis associated with other disease, MCP joint |
| 492981000006114 | Arthropathy in Reiter's disease |
| 493191000006117 | Arthropathy with other bacterial disease, of other spec site |
| 497081000006112 | Atlanto-axial joint sprain |
| 538831000006115 | Carpal instability, ulnar translocation |
| 563691000006115 | Closed fracture of sacrum with unspec spinal cord lesion |
| 564011000006117 | Closed fracture of tibia and fibula, proximal |
| 564031000006111 | Closed fracture of tibia and fibula, proximal NOS |
| 565211000006111 | Closed fracture, base of neck of femur |
| 565301000006117 | Closed fracture-dislocation lunate (volar) |
| 565321000006110 | Closed fracture-dislocation multiple digits |
| 565401000006114 | Closed fracture-dislocation shoulder joint |
| 565461000006110 | Closed fracture-dislocation, interphalangeal joint thumb |
| 565471000006115 | Closed fracture-dislocation, IPJ, multiple toes |
| 565781000006113 | Closed fracture-subluxation, distal radio-ulnar jt |
| 565891000006117 | Closed fracture-subluxation, subtalar joint |
| 565901000006118 | Closed fracture-subluxation, tarsometatarsal joint |
| 570651000006110 | Cls spinal # with incomplete thoracic cord lesion, T1-6 NOS |
| 603431000006110 | Crystal arthropathy NOS |
| 603451000006115 | Crystal arthropathy NOS, of 1st MTP joint |
| 709441000006113 | Fibromyalgia |
| 779421000006114 | Infective arthritis NOS, of MCP joint |
| 779591000006110 | Infective arthritis NOS, of tibio-fibular joint |
| 823951000006111 | Hip osteoarthitis NOS |
| 162231000006117 | Rheumatoid arthropathy + visceral/systemic involvement NOS |
| 211241000006115 | Postherpetic trigeminal neuralgia |
| 215061000006110 | Polyneuropathy in uraemia |
| 217001000006117 | Post-herpetic neuralgia |
| 255641000006113 | Open fracture-subluxation, ankle joint |
| 255671000006117 | Open fracture-subluxation, distal radio-ulnar joint |
| 255751000006117 | Open fracture-subluxation, metatarsophalangeal joint, single |
| 255981000006110 | Open fracture-dislocation peri-lunate (dorsal) |
| 256191000006118 | Open fracture-dislocation, proximal interphalangeal joint |
| 256201000006115 | Open fracture-dislocation, subtalar joint |
| 256221000006113 | Open fracture-subluxation acromio-clavicular joint |
| 256341000006113 | Open fracture-subluxation of the wrist |
| 256701000006112 | Open fracture-dislocation elbow |
| 256741000006114 | Open fracture-dislocation mid carpal |
| 265841000006115 | Oligoarticular osteoarthritis, unspec, of unspecified sites |
| 318531000006113 | [Q] Open fracture grade 2 |
| 375981000006118 | [X]Crush inj of oth & unspecif parts of abdom/low back/pelv |
| 422991000006117 | [X]Polyneuropathy/systemic connective tissue disorders CE |
| 318212019 | Open fracture mandible (site unspecified) |
| 318235016 | Fracture of malar or maxillary bones, open, NOS |
| 318679014 | Other specified closed fracture pubis |
| 318709018 | Other open fracture of pelvis |
| 318832012 | Open fracture of radius, shaft, unspecified |
| 318909012 | Closed fracture of radius and ulna, NOS |
| 318939019 | Fracture of other metacarpal bone |
| 318945010 | Closed fracture of metacarpal bone (s), site unspecified |
| 319019013 | Open fracture of phalanx or phalanges, unspecified |
| 319056016 | Multiple fractures of hand bones NOS |
| 319152017 | Closed fracture of shaft or unspecified part, NOS |
| 319170019 | Open fracture distal femur, unspecified |
| 319226017 | Open fracture of tibia and fibula, shaft, NOS |
| 319303014 | Open fracture of other tarsal and metatarsal bones |
| 320141017 | Other elbow sprain |
| 320256019 | Knee sprain NOS |
| 320280015 | Foot sprain NOS |
| 320544019 | Other and ill-defined sprains and strains |
| 320546017 | Jaw sprain, unspecified |
| 320566014 | Sternum sprain NOS |
| 323927014 | Other face and neck injuries |
| 325341011 | [X]Fracture of other parts of bony thorax |
| 325417012 | [X]Fracture of other carpal bone(s) |
| 391393013 | Hip fracture NOS |
| 400197012 | Arthropathy NOS, of the pelvic region and thigh |
| 402878019 | Closed fracture of other tarsal and metatarsal bones |
| 18181000006118 | Osteoarthritis of spine |
| 28831000006117 | Other infect/parasit dis with arthropathy of pelvic / thigh |
| 29261000006119 | Other infect/parasit dis with arthropathy of multiple sites |
| 84571000006115 | Type 2 diabetes mellitus with neuropathic arthropathy |
| 92731000006117 | Traumatic arthropathy of sacro-iliac joint |
| 131081000006113 | Sprain ulnar carpal complex non-specific |
| 140571000006110 | Shoulder blade fracture |
| 299349011 | [X]Other disorders of trigeminal nerve |
| 309537014 | Arthropathy associated with mycoses, of the forearm |
| 309597011 | Infective arthritis NOS, of the upper arm |
| 309608017 | Infective arthritis NOS, of wrist |
| 309618010 | Infective arthritis NOS, of ankle |
| 309626019 | Infective arthritis NOS, of other specified site |
| 309685017 | Other crystal arthropathies |
| 309686016 | Other crystal arthropathies of unspecified site |
| 309692010 | Other crystal arthropathies of the lower leg |
| 309701018 | Crystal arthropathy NOS, of the upper arm |
| 309764012 | Arthritis associated with other disease, elbow |
| 309880013 | Generalised osteoarthritis NOS |
| 309953017 | Localised osteoarthritis, unspecified, of the hand |
| 309975015 | Oligoarticular osteoarthritis, unspecified, of upper arm |
| 310016014 | Osteoarthritis NOS, of wrist |
| 310034013 | Other and unspecified arthropathies |
| 310105016 | Climacteric arthritis NOS |
| 310106015 | Transient arthropathy of unspecified site |
| 310167014 | Unspecified monoarthritis of the shoulder region |
| 310169012 | Unspecified monoarthritis of the forearm |
| 310182016 | Other specified arthropathy of unspecified site |
| 310197018 | Arthropathy NOS, of unspecified site |
| 311704019 | Myalgia or myositis NOS |
| 311706017 | Neuralgia unspecified |
| 311768018 | Nonarticular rheumatism NOS |
| 312571013 | [X]Other instability of joint |
| 312630014 | [X]Other dorsalgia |
| 312800015 | Spinal meningocele of unspecified site |
| 82989010 | Closed dislocation of sacroiliac joint |
| 253116010 | Headache site NOS |
| 1165801000000110 | Rheumatology service home visit |
| 875201000006117 | Therap.asp.- musculoskelet.NOS |
| 889891000006116 | Osteoarthritis - wrist joint |
| 896281000006113 | Sprain - lower leg |
| 896411000006117 | Sprained neck |
| 240071000006115 | Patellofemoral osteoarthritis |
| 299771000000110 | Closed fracture of femur, upper epiphysis |
| 559621000000119 | Suspected inflammatory arthritis |
| 2695828012 | Gouty arthritis of toe |
| 2791650019 | Irritable bowel syndrome characterised by constipation |
| 485443016 | Closed fracture of radius and ulna, lower end |
| 494318018 | Multiple fractures of arm |
| 1220743019 | Multiple fractures of ribs |
| 1221187014 | Closed fracture ankle, medial malleolus |
| 1221279016 | Sacroiliac ankylosis |
| 1223016015 | Open fracture intermediate cuneiform |
| 1225102013 | Open fracture cuboid |
| 1225771011 | Open fracture multiple ribs |
| 1227288016 | Closed fracture triquetral |
| 1228040014 | Closed fracture acetabulum |
| 1229092011 | Closed fracture hamate |
| 1229848013 | Closed fracture of cuneiforms |
| 1230401019 | Leg fracture |
| 1231972010 | Open fracture proximal fibula |
| 1232866011 | Open fracture lateral cuneiform |
| 1233086019 | Closed fracture of ulna, coronoid |
| 1233093015 | Closed fracture radius, head |
| 1233358018 | Closed fracture of the proximal radius |
| 1233512015 | Closed fracture radius, neck |
| 1234261013 | Open fracture of the proximal ulna |
| 1235130019 | Open fracture of ulna, coronoid |
| 1492276018 | Osteoarthritis of cervical spine |
| 1492281010 | Osteoarthritis of thoracic spine |
| 1494798013 | Sternum sprain |
| 1495246018 | Sprained toe |
| 1495445016 | Open fracture pelvis, ischium |
| 1495502013 | Wrist fracture - open |
| 1495503015 | Open fracture radius and ulna, distal |
| 1786065018 | Hereditary motor and sensory neuropathy type I |
| 2477021015 | Ochronotic arthropathy |
| 391096011 | Forearm sprain |
| 391105019 | Toe sprain |
| 391282013 | Multiple face fractures |
| 391352017 | Fracture of shaft of radius |
| 393896016 | Intracranial destruction of trigeminal nerve (V) |
| 400247016 | Arthralgia of the upper arm |
| 402839018 | Closed fracture of sixth cervical vertebra |
| 402844013 | Open fracture of third cervical vertebra |
| 443817012 | Elbow fracture - open |
| 451060018 | Open Barton's fracture |
| 455431014 | Prolapsed lumbar intervertebral disc with sciatica |
| 455446019 | Closed multiple fractures of clavicle, scapula and humerus |
| 459740018 | Intractable breast pain |
| 461560011 | [Q] Open fracture grade 3C |
| 473460018 | Charcot's arthropathy |
| 475273015 | Ulnar neuropathy |
| 478502010 | Finger fracture |
| 295342013 | Psychogenic musculoskeletal symptoms |
| 297553017 | Polyneuropathy in disease EC |
| 299136013 | Ossicle ankylosis (excluding malleus) |
| 309552019 | Helminthiasis with arthropathy of the hand |
| 309750012 | Arthropathy associated with respiratory disorders |
| 309881012 | Localised, primary osteoarthritis |
| 309898012 | Localised, primary osteoarthritis of the ankle and foot |
| 309918012 | Localised, secondary osteoarthritis of the shoulder region |
| 310050017 | Traumatic arthropathy of the shoulder region |
| 310056011 | Traumatic arthropathy of the ankle and foot |
| 310068019 | Traumatic arthropathy-hip |
| 310070011 | Traumatic arthropathy-knee |
| 4765019 | Postherpetic neuralgia |
| 29800015 | Closed fracture of five ribs |
| 35988013 | Open wound of back with complication |
| 50702011 | Mononeuritis multiplex |
| 72981015 | Glossopharyngeal neuralgia |
| 81242018 | Closed fracture of hyoid bone |
| 84036012 | Palindromic rheumatism |
| 90489010 | Fracture of shaft of femur |
| 107139013 | Fracture of acetabulum |
| 118266012 | Open fracture of pubis |
| 119023019 | Fracture of skull |
| 128724013 | Open fracture of six ribs |
| 129260010 | Menopausal arthritis |
| 132103012 | Closed fracture of astragalus |
| 133435011 | Open Colles' fracture |
| 138248010 | Basilar migraine |
| 143436010 | Idiopathic peripheral autonomic neuropathy |
| 249890015 | FH: Migraine |
| 252314011 | C/O - low back pain |
| 253109014 | Bilateral headache |
| 253115014 | Parietal headache |
| 253118011 | Aching headache |
| 257897018 | Rheumatoid factor negative |
| 262959010 | Headache caused by oral contraceptive pill |
| 310072015 | Traumatic arthropathy-ankle |
| 310082019 | Allergic arthritis of the shoulder region |
| 310104017 | Climacteric arthritis of multiple sites |
| 311194016 | Idiopathic cervical spinal stenosis |
| 311195015 | Degenerative cervical spinal stenosis |
| 311211016 | Rheumatic torticollis |
| 311234013 | Iatrogenic thoracic spinal stenosis |
| 311256011 | Thoracic and lumbosacral neuritis |
| 311272015 | Sacroiliac disorder |
| 318204010 | Closed fracture of mandible, angle of jaw |
| 318236015 | Fracture of skull and facial bones |
| 318375010 | Closed fracture atlas, isolated arch or articular process |
| 318387015 | Closed fracture cervical vertebra, transverse process |
| 318420019 | Open fracture axis, odontoid process |
| 318426013 | Open fracture cervical vertebra, burst |
| 318428014 | Open fracture cervical vertebra, spondylolysis |
| 318447012 | Open fracture thoracic vertebra, burst |
| 318449010 | Open fracture thoracic vertebra, spondylolysis |
| 318463016 | Open fracture lumbar vertebra, burst |
| 318467015 | Open fracture lumbar vertebra, transverse process |
| 318528011 | Closed fracture of thoracic spine with spinal cord lesion |
| 318562011 | Closed spinal fracture with cauda equina lesion |
| 318581015 | Open fracture of sacrum with spinal cord lesion |
| 318642011 | Cough fracture |
| 318657016 | Closed fracture acetabulum, anterior column |
| 318674016 | Closed fracture pelvis, single pubic ramus |
| 318675015 | Closed fracture pelvis, multiple pubic rami - stable |
| 318694013 | Closed fracture pelvis, iliac wing |
| 318737013 | Closed fracture scapula, glenoid |
| 318739011 | Closed fracture scapula, spine |
| 318751013 | Closed fracture proximal humerus, neck |
| 318752018 | Closed fracture of proximal humerus, anatomical neck |
| 318753011 | Closed fracture proximal humerus, greater tuberosity |
| 318771016 | Open fracture proximal humerus, four part |
| 318789015 | Closed fracture distal humerus, capitellum |
| 318805019 | Open fracture of distal humerus, multiple |
| 318848013 | Smith's fracture - closed |
| 318882019 | Open volar Barton's fracture |
| 318922016 | Closed fracture scaphoid, tuberosity |
| 318943015 | Hand fracture - metacarpal bone |
| 318944014 | Closed fracture of metacarpal bone(s) |
| 318976016 | Open fracture thumb metacarpal head |
| 318997015 | Closed fracture thumb distal phalanx, tuft |
| 319000015 | Closed fracture finger proximal phalanx, base |
| 319018017 | Open fracture of one or more phalanges of hand |
| 319022010 | Open fracture thumb proximal phalanx |
| 319024011 | Open fracture thumb proximal phalanx, shaft |
| 319054018 | Closed multiple fractures of hand bones |
| 319055017 | Open multiple fractures of hand bones |
| 319075014 | Fractures involving multiple regions of both upper limbs |
| 319092017 | Closed fracture proximal femur, subcapital, Garden grade I |
| 319109017 | Open fracture proximal femur,subcapital, Garden grade I |
| 319176013 | Open fracture distal femur, comminuted/intra-articular |
| 319193013 | Open fracture patella, comminuted (stellate) |
| 319212014 | Open fracture proximal tibia, medial condyle (plateau) |
| 319225018 | Open fracture of tibia and fibula, shaft |
| 319231015 | Open fracture distal tibia, extra-articular |
| 319252011 | Closed fracture ankle, lateral malleolus, low |
| 319284016 | Closed fracture calcaneus, intra-articular |
| 319285015 | Open fractures calcaneus, extra-articular |
| 319307010 | Open fracture talus, body |
| 319312011 | Open fracture metatarsal, multiple |
| 319324016 | Open fracture proximal phalanx, toe |
| 319325015 | Open fracture middle phalanx, toe |
| 319341019 | Multiple fractures of lower leg |
| 319763013 | Closed spinal dislocation with complete thoracic cord lesion |
| 320114010 | Coracohumeral sprain |
| 320159019 | Sprain ulnar-carpal meniscus |
| 320162016 | Sprain ulnar collateral ligament |
| 320242016 | Sprain or partial tear, knee, lateral collateral ligament |
| 320278014 | Sprain, flexor tendon, foot |
| 320545018 | Septal cartilage nose sprain |
| 323826018 | Closed injury sciatic nerve |
| 345355014 | Status migrainosus |
| 297569012 | Polyneuropathy in disease NOS |
| 309922019 | Localised, secondary osteoarthritis of the forearm |
| 309934010 | Localised, secondary osteoarthritis of other specified site |
| 320311012 | Sacral sprain, unspecified |
| 400181019 | Localised osteoarthritis, unspecified, of the lower leg |
| 493271000006113 | Arthropathy with other viral disease, of lower leg |
| 891761000006119 | Myalgia/myositis - NOS |
| 891831000006115 | Neuralgia/neuritis - shoulder |
| 892591000006113 | Fracture malunion - NOS |
| 896161000006110 | Dislocations/sprains NOS |
| 990351000006115 | Sprain - fore arm |
| 889851000006110 | Polyarthropathy NOS -inflammat |
| 257908018 | Rheumatoid factor NOS |
| 309849015 | Other specified inflammatory polyarthropathy NOS |
| 684221000006112 | Myalgic encephalomyelitis |
| 400222018 | Joint ankylosis of the upper arm |
| 889621000006117 | Arthritis/arthrosis |
| 892421000006112 | Path.fracture - lower leg |
| 415621000006114 | [X]Other specified injuries of abdomen, lower back & pelvis |
| 427421000006112 | [X]Sprain & strain of oth & unsp parts of lumb spine & pelv |
| 483211000006115 | Angular mal-union of fracture |
| 484171000006115 | Ankylosis/instability of cervical,thoracic or lumbar spine |
| 492731000006111 | Arthropathy associated with other conditions EC |
| 493151000006111 | Arthropathy with other bacterial disease, of forearm |
| 493171000006118 | Arthropathy with other bacterial disease, of lower leg |
| 497061000006119 | Atlanto-axial ankylosis |
| 523461000006116 | Bouchard's nodes with arthropathy |
| 531691000006114 | C6 vertebra open fracture without spinal cord lesion |
| 561311000006117 | Closed dorsal Barton fracture-subluxation |
| 563791000006110 | Closed fracture of skull NOS without intracranial injury |
| 565311000006119 | Closed fracture-dislocation mid carpal |
| 565811000006110 | Closed fracture-subluxation, interphalangeal joint thumb |
| 567561000006119 | Closed spinal dislocation with cervical cord lesion, unspec |
| 567601000006119 | Closed spinal dislocation with lumbar cord lesion, unspec |
| 570631000006115 | Cls spinal # with incomplete cervical cord lesion, C1-4 NOS |
| 700211000006115 | Moebius' ophthalmoplegic migraine |
| 735201000006116 | Late effect of skull fracture |
| 736711000006119 | Localised osteoarthritis, unspecified, of shoulder region |
| 162191000006110 | Rheumatoid arthritis of tibio-fibular joint |
| 211331000006117 | Postinfectious polyneuritis |
| 217671000006115 | Postdysenteric reactive arthropathy of multiple sites |
| 253131000006110 | Other/multiple open fracture of pelvis NOS |
| 255581000006118 | Open fracture-subluxation radiocarpal joint |
| 256121000006115 | Open fracture-dislocation, IPJ, multiple toes |
| 256161000006114 | Open fracture-dislocation, metatarsophalangeal joint, single |
| 256271000006114 | Open fracture-subluxation IPJ, unspecified |
| 256661000006116 | Open fracture-dislocation acromio-clavicular joint |
| 256751000006111 | Open fracture-dislocation multiple digits |
| 256761000006113 | Open fracture-dislocation of pelvis |
| 257931000006116 | Open fracture of tibia and fibula, proximal |
| 259611000006111 | Open fracture base skull without mention intracranial injury |
| 259861000006110 | Open fracture distal humerus, lateral condyle |
| 264471000006113 | Oligoarticular osteoarthritis, unspecified, of pelvis/thigh |
| 363801000006113 | [X]Arthritis in other infectious and parasitic diseases CE |
| 311035015 | Other specified arthropathies |
| 311240018 | Lumbar spinal stenosis secondary to other disease |
| 311271010 | Sacral instability NOS |
| 311746011 | Other musculoskeletal limb symptoms |
| 312527014 | [X]Other specified crystal arthropathies |
| 312529012 | [X]Other specified arthritis |
| 312792011 | Spinal hydromeningocele NOS |
| 318207015 | Closed fracture of mandible, body, other and unspecified |
| 318229016 | Fracture of malar or maxillary bones, closed, NOS |
| 318434019 | Open fracture of cervical spine not otherwise specified |
| 318495015 | Fracture of spine without mention of spinal cord lesion NOS |
| 318557010 | Closed spinal fracture with unspecified lumbar cord lesion |
| 318574010 | Closed fracture of sacrum with other cauda equina injury |
| 318685019 | Other specified open fracture of pubis |
| 318686018 | Open fracture of pubis NOS |
| 318720018 | Fracture of ill-defined bones of trunk |
| 318762013 | Open fracture of proximal humerus, unspecified part |
| 318779019 | Open fracture of humerus, shaft or unspecified part NOS |
| 318910019 | Open fracture of radius and ulna, unspecified part |
| 318916013 | Closed fracture of carpal bone, unspecified |
| 319153010 | Open fracture of femur, shaft or unspecified part |
| 319318010 | Fracture of tarsal and metatarsal bones NOS |
| 319353015 | Open fracture of bones, unspecified |
| 320237012 | Thigh sprain NOS |
| 320251012 | Other specified knee sprain |
| 320252017 | Other specified leg sprain |
| 320270019 | Ankle sprain NOS |
| 320271015 | Foot sprain, unspecified |
| 320313010 | Sacrum sprain NOS |
| 320576012 | Sprain of pelvis NOS |
| 320578013 | Other specified sprains and strains |
| 325381016 | [X]Fracture of other parts of shoulder and upper arm |
| 325400017 | [X]Fracture of forearm, unspecified |
| 325418019 | [X]Fracture of other metacarpal bone |
| 325485012 | [X]Fractures involving other combinations of body regions |
| 400165012 | Arthropathy in Behcet's syndrome of unspecified site |
| 400198019 | Arthropathy NOS, of the lower leg |
| 400223011 | Joint ankylosis of the forearm |
| 402875016 | Closed fracture of femur, unspecified part |
| 402876015 | Closed fracture of tibia and fibula, unspecified part, NOS |
| 460303019 | [V]Personal history of arthritis |
| 28241000006111 | Other infect/parasit dis with arthropathy of shoulder region |
| 34231000006110 | Other congenital musculoskeletal deformity |
| 34791000006116 | Other closed fracture-dislocation |
| 43221000006113 | Opn spinal fracture with unspec thoracic cord lesion, T1-6 |
| 297351016 | Migraine variant NOS |
| 297356014 | Migraine NOS |
| 297494010 | Mononeuritis upper limb NOS |
| 297505016 | Other mononeuritis lower limb |
| 297506015 | Unspecified mononeuritis lower limb |
| 297545013 | Polyneuropathy in collagen vascular disease NOS |
| 309551014 | Helminthiasis with arthropathy of the forearm |
| 309604015 | Infective arthritis NOS, of sternoclavicular joint |
| 309675018 | Gouty arthritis of the upper arm |
| 309676017 | Gouty arthritis of the forearm |
| 309697016 | Other crystal arthropathy NOS |
| 309771019 | Arthritis associated with other disease, hip |
| 309896011 | Localised, primary osteoarthritis of the lower leg |
| 310081014 | Allergic arthritis of unspecified site |
| 310083012 | Allergic arthritis of the upper arm |
| 310098014 | Climacteric arthritis of the forearm |
| 310156013 | Unspecified polyarthropathy of other specified site |
| 310172017 | Unspecified monoarthritis of the lower leg |
| 310181011 | Other specified arthropathy |
| 310217012 | Arthropathy NOS, of multiple sites |
| 310444016 | Carpal instability, other |
| 310557014 | Ankylosis of joint NOS |
| 317142014 | [D]Other nervous and musculoskeletal symptoms |
| 2129721000000119 | Management of irritable bowel syndrome |
| 1756651000000117 | Frequent episodic tension-type headache |
| 1730651000000118 | Greenstick fracture of distal radius |
| 889861000006112 | Osteoarthritis -multiple joint |
| 890391000006115 | Ankylosis - shoulder joint |
| 895331000006114 | #Thoracic spine + cord lesion |
| 896311000006110 | Sprain - medial knee ligament |
| 693461000006115 | Musculoskeletal and connective tissue diseases |
| 807141000006110 | Greenstick fracture |
| 557111000000111 | Minimal trauma fracture |
| 2536004017 | Closed fracture maxilla |
| 495040011 | Pyogenic arthritis |
| 502690012 | Pauciarticular onset juvenile chronic arthritis |
| 504101018 | Polyneuropathy in beriberi |
| 508047017 | Osteoarthritis of spine |
| 1219672018 | Open fracture clavicle, medial end |
| 1219673011 | Closed fracture scapula, acromion |
| 1220723013 | Fracture of thoracic vertebra |
| 1221007014 | Open fracture of metacarpal bone(s) |
| 1221607010 | Hip sprain |
| 1228028014 | Closed fracture of the proximal ulna |
| 1228196017 | Open fracture hamate |
| 1228298012 | Arthralgia of multiple joints |
| 1229331011 | Closed fracture lunate |
| 1229426013 | Open fracture scapula, acromion |
| 1229710011 | Closed fracture of ulna, styloid process |
| 1229819019 | Closed fracture lateral cuneiform |
| 1231198017 | Open fracture lunate |
| 1232763011 | Closed fracture sternum |
| 1233006016 | Open fracture of the scaphoid |
| 1233864019 | Closed fracture navicular |
| 1234315010 | Open fracture triquetral |
| 1234676011 | Hand fracture - carpal bone |
| 1235228012 | Closed fracture clavicle, shaft |
| 1494730015 | Open fracture of mandible, alveolar border of body |
| 399416017 | Inflammatory and toxic neuropathy |
| 400245012 | Arthralgia of the shoulder region |
| 400276018 | Brachial (cervical) neuritis |
| 400282015 | Spinal stenosis, excluding cervical region |
| 402914014 | Sprain of knee and leg |
| 405144016 | Pathological fracture of lumbar vertebra |
| 411304016 | Shoulder fracture - open |
| 411305015 | Closed fracture of femur, subcapital |
| 411308018 | Closed fracture of femur, lesser trochanter |
| 411309014 | Open fracture of femur, lesser trochanter |
| 411310016 | Open fracture of femur, greater trochanter |
| 415489016 | Fracture of lower end of tibia |
| 416139010 | Low back pain |
| 455485014 | Closed fractures involving head with neck |
| 455486010 | Open fractures involving head with neck |
| 456415013 | Late effect of fracture of thoracic vertebra |
| 459296014 | Type 1 diabetes mellitus with neuropathic arthropathy |
| 480728016 | Fracture of bone of hand |
| 484410014 | Sacroiliac instability |
| 283788016 | Fracture therapy follow-up |
| 297337016 | Migraine variants |
| 297543018 | Polyneuropathy in polyarteritis nodosa |
| 251795011 | H/O: osteoarthritis |
| 252573011 | Abdominal migraine - symptom |
| 253105015 | Generalised headache |
| 257896010 | Rheumatoid factor positive |
| 6555013 | Closed fracture of two ribs |
| 7278014 | Arthritis |
| 10843018 | Fracture of neck of femur |
| 19796013 | Hereditary sensory neuropathy |
| 30978014 | Lumbar spinal stenosis |
| 38727013 | Sciatica |
| 48573010 | Open fracture of carpal bone |
| 62810012 | Late effect of fracture of skull and face bones |
| 74641011 | Gonococcal arthritis |
| 79031011 | Open fracture of two ribs |
| 126030010 | Abdominal migraine |
| 130281012 | Fracture of sternum |
| 136121013 | Fracture of carpal bone |
| 136713011 | Chronic pain |
| 158058014 | Intercostal myalgia |
| 194542018 | Fracture of sacrum |
| 309489019 | Arthropathy in Behcet's syndrome of the ankle and foot |
| 309563016 | Reactive arthropathy of acromioclavicular joint |
| 309674019 | Gouty arthritis of the shoulder region |
| 309743013 | Arthropathy in ulcerative colitis |
| 309789018 | Rheumatoid arthritis of shoulder |
| 309798015 | Rheumatoid arthritis of hip |
| 309800010 | Rheumatoid arthritis of knee |
| 309833017 | Juvenile arthritis in Crohn's disease |
| 310059016 | Traumatic arthropathy of shoulder |
| 310064017 | Traumatic arthropathy-wrist |
| 310121014 | Transient arthropathy-elbow |
| 310137015 | Transient arthropathy of subtalar joint |
| 311113014 | Enterobacterial spondylitis |
| 311281014 | Atlanto-occipital ankylosis |
| 311283012 | Cervical spine ankylosis |
| 311305016 | Fatigue fracture of vertebra |
| 311681019 | Spasm of back muscles |
| 318213012 | Open fracture of mandible, condylar process |
| 318249013 | Fracture of alveolus, open |
| 318344018 | Fracture of neck and trunk |
| 318377019 | Closed fracture axis, odontoid process |
| 318382014 | Closed fracture axis, tricolumnar |
| 318383016 | Closed fracture cervical vertebra, burst |
| 318385011 | Closed fracture cervical vertebra, spondylolysis |
| 318435018 | Closed fracture thoracic vertebra |
| 318436017 | Closed fracture thoracic vertebra, burst |
| 318457013 | Closed fracture lumbar vertebra, spondylolysis |
| 318458015 | Closed fracture lumbar vertebra, spinous process |
| 318459011 | Closed fracture lumbar vertebra, transverse process |
| 318462014 | Open fracture lumbar vertebra |
| 318465011 | Open fracture lumbar vertebra, spondylolysis |
| 318571019 | Closed fracture of sacrum with spinal cord lesion |
| 318589018 | Closed fracture of coccyx with complete cauda equina lesion |
| 318655012 | Closed fracture acetabulum, anterior lip alone |
| 318667014 | Open fracture acetabulum, posterior column |
| 318668016 | Open fracture acetabulum, floor |
| 318669012 | Open fracture acetabulum, double column transverse |
| 318692012 | Closed fracture pelvis, anterior superior iliac spine |
| 318747011 | Open fracture scapula, neck |
| 318763015 | Open fracture proximal humerus, neck |
| 318764014 | Open fracture of proximal humerus, anatomical neck |
| 318802016 | Open fracture distal humerus, lateral epicondyle |
| 318819019 | Closed fracture olecranon, intra-articular |
| 318827011 | Open fracture olecranon, intra-articular |
| 318833019 | Open fracture radius and ulna, middle |
| 318881014 | Open Galeazzi fracture |
| 318918014 | Closed fracture scaphoid, proximal pole |
| 318920012 | Closed fracture scaphoid, waist, oblique |
| 318921011 | Closed fracture scaphoid, waist, comminuted |
| 318947019 | Closed fracture finger metacarpal base |
| 318950016 | Closed fracture finger metacarpal head |
| 318953019 | Closed fracture of thumb metacarpal |
| 318955014 | Closed fracture thumb metacarpal shaft |
| 318956010 | Closed fracture thumb metacarpal neck |
| 318968018 | Open fracture finger metacarpal neck |
| 318974018 | Open fracture thumb metacarpal shaft |
| 318975017 | Open fracture thumb metacarpal neck |
| 318992014 | Closed fracture thumb proximal phalanx, neck |
| 318993016 | Closed fracture thumb proximal phalanx, head |
| 318995011 | Closed fracture thumb distal phalanx, base |
| 319014015 | Closed fracture finger distal phalanx, mallet |
| 319027016 | Open fracture thumb distal phalanx |
| 319033013 | Open fracture finger proximal phalanx, base |
| 319087019 | Closed fracture proximal femur, midcervical section |
| 319090013 | Closed fracture head of femur |
| 319093010 | Closed fracture proximal femur, subcapital, Garden grade II |
| 319110010 | Open fracture proximal femur,subcapital, Garden grade II |
| 319128012 | Open fracture of proximal femur, pertrochanteric |
| 319138019 | Pertrochanteric fracture |
| 319164013 | Closed fracture distal femur, bicondylar (T-Y fracture) |
| 319181016 | Closed fracture patella, transverse |
| 319186014 | Closed fracture patella, vertical |
| 319222015 | Closed fracture of tibia and fibula, shaft |
| 319263014 | Open fracture ankle, bimalleolar, low fibular fracture |
| 319267010 | Open fracture ankle, trimalleolar, low fibular fracture |
| 319313018 | Open tarsal fractures, multiple |
| 319779014 | Open spinal dislocation with complete thoracic cord lesion |
| 319837010 | Closed spinal subluxation with anterior cervical cord lesion |
| 319870017 | Closed spinal subluxation with complete lumbar cord lesion |
| 320136013 | Sprain, elbow joint, lateral collateral ligament |
| 320138014 | Sprain, elbow joint, ulnar collateral ligament |
| 320160012 | Sprain triangular fibrocartilage |
| 320177012 | Carpometacarpal sprain |
| 320179010 | Interphalangeal sprain |
| 320214018 | Sprain, flexor pollicis longus tendon |
| 320215017 | Sprain, extensor pollicis longus tendon |
| 320216016 | Sprain tendon of finger |
| 320227019 | Iliofemoral sprain |
| 320310013 | Sacrum sprain |
| 320569019 | Sprain, symphysis pubis |
| 345594014 | Axonal sensorimotor neuropathy |
| 484753013 | Arthralgia of knee |
| 256125011 | O/E - musculoskeletal |
| 299359012 | Mononeuropathy of upper limb |
| 310027011 | Osteoarthritis of subtalar joint |
| 317145011 | Musculoskeletal pain |
| 318248017 | Open fracture of facial bones |
| 318494016 | Open fracture of vertebral column |
| 318846012 | Closed fracture of distal end of ulna |
| 318859016 | Closed extraarticular fracture of distal radius |
| 318890019 | Open extra-articular fracture of distal radius |
| 319147010 | Closed fracture of shaft of femur |
| 319242012 | Closed fracture of fibula |
| 319331017 | Fracture of toe |
| 319339015 | Open fracture of multiple bones of lower limb |
| 359389010 | Osteoarthritis of finger |
| 391102016 | Sprain of joint |
| 399415018 | Mononeuropathy |
| 402874017 | Closed fracture of neck of femur |
| 402912013 | Sprain of hand |
| 500272010 | Fracture of forearm |
| 44451000006118 | Open spinal fracture with complete cervical cord lesion, C1-4 |
| 65811000006110 | Vascular headache |
| 92621000006111 | Traumatic arthropathy of 1st metatarsophalangeal joint |
| 130581000006112 | Late effect of sprain AND/OR strain without tendon injury |
| 130661000006118 | Sprain finger, proximal interphalangeal joint, radial collateral ligament |
| 130681000006111 | Sprain of metacarpophalangeal joint |
| 131001000006116 | Sprain thumb, interphalangeal joint, radial collateral ligament |
| 131351000006115 | Strain of hamstring tendon |
| 131361000006118 | Sprain of hip joint |
| 131421000006118 | Strain of long head of biceps |
| 162041000006117 | Rheumatoid arthritis of distal interphalangeal joint of finger |
| 162131000006111 | Rheumatoid arthritis of proximal interphalangeal joint of finger |
| 212551000006110 | Post-zoster neuralgia |
| 215271000000119 | Neck pain |
| 222331000000113 | Closed fracture of distal end of femur |
| 254521000006118 | Open multiple fractures involving both upper limbs, and upper limb with rib(s) and sternum |
| 257211000006119 | Open fracture of tibia or fibula, shaft |
| 257771000006115 | Open fracture of patella |
| 257921000006119 | Open fracture of upper end of lower leg |
| 399821000006115 | Neuropathy |
| 531661000006118 | Closed fracture of fifth cervical vertebra without spinal cord injury |
| 559791000006116 | Closed dorsal Barton's fracture |
| 560291000006113 | Closed fracture of vault of skull with intracranial injury, with more than 24 hours loss of consciousness and return to pre-existing conscious level |
| 561661000006110 | Closed fracture of base of skull without intracranial injury |
| 565261000006114 | Closed fracture dislocation elbow joint |
| 565531000006119 | Closed fracture dislocation, proximal interphalangeal joint |
| 567641000006117 | Closed spinal dislocation with posterior thoracic cord lesion |
| 567711000006112 | Closed spinal subluxation with posterior cervical cord lesion |
| 570591000006115 | Closed fracture thumb metacarpal base, intra-articular, Rolando |
| 570701000006116 | Closed spinal fracture with central cervical cord lesion , C1-4 |
| 570761000006115 | Closed spinal fracture with complete thoracic cord lesion,T7-12 |
| 570781000006113 | Closed spinal fracture with posterior cervical cord lesion, C5-7 |
| 696421000006117 | Multiple fractures involving both upper limbs, and upper limb with rib(s) and sternum |
| 750721000006113 | Late effect of musculoskeletal and connective tissue injuries |
| 757841000006114 | Ankylosis of joint of hand |
| 775571000006111 | Injury of muscle and tendon of abdomen, lower back and pelvis |
| 780181000006118 | Infective arthritis |
| 793941000006118 | Fracture dislocation/subluxation finger/thumb |
| 1756451000006110 | Fracture of tibial plateau |
| 309720016 | Crystal arthropathy of knee |
| 377491000006117 | [X]Dislocation, sprain and strain of unspecified joint and ligament of trunk |
| 409041000006110 | [X]Other disorders of the musculoskeletal system and connective tissue |

### **Supplementary Table S2. Opioid product code list**

| **prodcodeid** | **Product name** |
| --- | --- |
| 1029741000033111 | Paracetamol & Codeine Dispersible tablets |
| 3343341000033117 | Buprenorphine 5micrograms/hour transdermal patches |
| 11029341000033114 | Butec 5micrograms/hour transdermal patches (Qdem Pharmaceuticals Ltd) |
| 13751341000033119 | Rebrikel 5micrograms/hour transdermal patches (Zentiva) |
| 12603441000033118 | Bunov 5micrograms/hour transdermal patches (Glenmark Pharmaceuticals Europe Ltd) |
| 12325741000033112 | Bupramyl 5micrograms/hour transdermal patches (Mylan) |
| 11756241000033110 | Sevodyne 5micrograms/hour transdermal patches (Aspire Pharma Ltd) |
| 11732541000033112 | Reletrans 5micrograms/hour transdermal patches (Sandoz Ltd) |
| 3343641000033113 | BuTrans 5micrograms/hour transdermal patches (Napp Pharmaceuticals Ltd) |
| 11730941000033119 | Panitaz 5micrograms/hour transdermal patches (Dr Reddy's Laboratories (UK) Ltd) |
| 12409541000033116 | Busiete 5micrograms/hour transdermal patches (Teva UK Ltd) |
| 10598241000033114 | Yemex 12micrograms/hour transdermal patches (Sandoz Ltd) |
| 4022641000033116 | Matrifen 12micrograms/hour transdermal patches (Teva UK Ltd) |
| 6440741000033114 | Fencino 12micrograms/hour transdermal patches (Ethypharm UK Ltd) |
| 11507641000033116 | Victanyl 12micrograms/hour transdermal patches (Accord Healthcare Ltd) |
| 3839341000033117 | Fentanyl 12micrograms/hour transdermal patches |
| 8884441000033117 | Opiodur 12micrograms/hour transdermal patches (Zentiva) |
| 3839441000033111 | Durogesic DTrans 12micrograms/hour transdermal patches (Janssen-Cilag Ltd) |
| 4386641000033113 | Mezolar Matrix 12micrograms/hour transdermal patches (Sandoz Ltd) |
| 9204341000033115 | Mylafent 12micrograms/hour transdermal patches (Mylan) |
| 5300441000033116 | Osmanil 12micrograms/hour transdermal patches (Zentiva) |
| 12106841000033113 | Butec 15micrograms/hour transdermal patches (Qdem Pharmaceuticals Ltd) |
| 13717441000033113 | Sevodyne 15micrograms/hour transdermal patches (Aspire Pharma Ltd) |
| 11077241000033112 | BuTrans 15micrograms/hour transdermal patches (Napp Pharmaceuticals Ltd) |
| 11732341000033117 | Reletrans 15micrograms/hour transdermal patches (Sandoz Ltd) |
| 11077041000033116 | Buprenorphine 15micrograms/hour transdermal patches |
| 12409641000033115 | Busiete 10micrograms/hour transdermal patches (Teva UK Ltd) |
| 11756341000033117 | Sevodyne 10micrograms/hour transdermal patches (Aspire Pharma Ltd) |
| 11732241000033110 | Reletrans 10micrograms/hour transdermal patches (Sandoz Ltd) |
| 3343741000033116 | BuTrans 10micrograms/hour transdermal patches (Napp Pharmaceuticals Ltd) |
| 3343441000033111 | Buprenorphine 10micrograms/hour transdermal patches |
| 11730741000033117 | Panitaz 10micrograms/hour transdermal patches (Dr Reddy's Laboratories (UK) Ltd) |
| 12603241000033119 | Bunov 10micrograms/hour transdermal patches (Glenmark Pharmaceuticals Europe Ltd) |
| 12325941000033110 | Bupramyl 10micrograms/hour transdermal patches (Mylan) |
| 11029441000033115 | Butec 10micrograms/hour transdermal patches (Qdem Pharmaceuticals Ltd) |
| 575041000033113 | Fentanyl 25micrograms/hour transdermal patches |
| 10336841000033116 | Mylafent 25micrograms/hour transdermal patches (Mylan) |
| 5007441000033116 | Victanyl 25micrograms/hour transdermal patches (Accord Healthcare Ltd) |
| 3333041000033118 | Tilofyl 25micrograms/hour transdermal patches (Tillomed Laboratories Ltd) |
| 4022741000033113 | Matrifen 25micrograms/hour transdermal patches (Teva UK Ltd) |
| 4502841000033111 | Osmach 25micrograms/hour transdermal patches (Teva UK Ltd) |
| 6440841000033116 | Fencino 25micrograms/hour transdermal patches (Ethypharm UK Ltd) |
| 8884541000033116 | Opiodur 25micrograms/hour transdermal patches (Zentiva) |
| 4956041000033118 | Osmanil 25micrograms/hour transdermal patches (Zentiva) |
| 4426341000033119 | Fentalis Reservoir 25micrograms/hour transdermal patches (Sandoz Ltd) |
| 490341000033116 | Durogesic 25micrograms transdermal patches (Janssen-Cilag Ltd) |
| 4387041000033117 | Mezolar Matrix 25micrograms/hour transdermal patches (Sandoz Ltd) |
| 3248041000033118 | Durogesic DTrans 25micrograms/hour transdermal patches (Janssen-Cilag Ltd) |
| 10598141000033119 | Yemex 25micrograms/hour transdermal patches (Sandoz Ltd) |
| 12326041000033117 | Bupramyl 20micrograms/hour transdermal patches (Mylan) |
| 12409741000033112 | Busiete 20micrograms/hour transdermal patches (Teva UK Ltd) |
| 12603341000033112 | Bunov 20micrograms/hour transdermal patches (Glenmark Pharmaceuticals Europe Ltd) |
| 11029541000033119 | Butec 20micrograms/hour transdermal patches (Qdem Pharmaceuticals Ltd) |
| 3343841000033114 | BuTrans 20micrograms/hour transdermal patches (Napp Pharmaceuticals Ltd) |
| 11730841000033110 | Panitaz 20micrograms/hour transdermal patches (Dr Reddy's Laboratories (UK) Ltd) |
| 11756441000033111 | Sevodyne 20micrograms/hour transdermal patches (Aspire Pharma Ltd) |
| 11732441000033111 | Reletrans 20micrograms/hour transdermal patches (Sandoz Ltd) |
| 3343541000033112 | Buprenorphine 20micrograms/hour transdermal patches |
| 2737541000033111 | Buprenorphine 35micrograms/hour transdermal patches |
| 12194941000033112 | Relevtec 35micrograms/hour transdermal patches (Sandoz Ltd) |
| 12428441000033117 | Turgeon 35micrograms/hour transdermal patches (Teva UK Ltd) |
| 11484341000033112 | Bupeaze 35micrograms/hour transdermal patches (Dr Reddy's Laboratories (UK) Ltd) |
| 8882741000033116 | Hapoctasin 35micrograms/hour transdermal patches (Accord Healthcare Ltd) |
| 11780641000033117 | Buplast 35micrograms/hour transdermal patches (Mylan) |
| 11507941000033111 | Prenotrix 35micrograms/hour transdermal patches (Genesis Pharmaceuticals Ltd) |
| 12603541000033117 | Carlosafine 35micrograms/hour transdermal patches (Glenmark Pharmaceuticals Europe Ltd) |
| 2737841000033113 | Transtec 35micrograms/hour transdermal patches (Napp Pharmaceuticals Ltd) |
| 8962341000033111 | Mezolar Matrix 37.5microgram/hour transdermal patches (Sandoz Ltd) |
| 8962241000033118 | Fentanyl 37.5microgram/hour transdermal patches |
| 11469241000033116 | Fentanyl 40micrograms/dose transdermal system |
| 12603641000033116 | Carlosafine 52.5micrograms/hour transdermal patches (Glenmark Pharmaceuticals Europe Ltd) |
| 11484441000033118 | Bupeaze 52.5micrograms/hour transdermal patches (Dr Reddy's Laboratories (UK) Ltd) |
| 8882841000033114 | Hapoctasin 52.5micrograms/hour transdermal patches (Accord Healthcare Ltd) |
| 2737941000033117 | Transtec 52.5micrograms/hour transdermal patches (Napp Pharmaceuticals Ltd) |
| 2737641000033112 | Buprenorphine 52.5micrograms/hour transdermal patches |
| 11780741000033114 | Buplast 52.5micrograms/hour transdermal patches (Mylan) |
| 11577141000033119 | Prenotrix 52.5micrograms/hour transdermal patches (Genesis Pharmaceuticals Ltd) |
| 12428541000033116 | Turgeon 52.5micrograms/hour transdermal patches (Teva UK Ltd) |
| 12195041000033112 | Relevtec 52.5micrograms/hour transdermal patches (Sandoz Ltd) |
| 5007541000033115 | Victanyl 50micrograms/hour transdermal patches (Accord Healthcare Ltd) |
| 10598041000033118 | Yemex 50micrograms/hour transdermal patches (Sandoz Ltd) |
| 3248141000033119 | Durogesic DTrans 50micrograms/hour transdermal patches (Janssen-Cilag Ltd) |
| 6440941000033112 | Fencino 50micrograms/hour transdermal patches (Ethypharm UK Ltd) |
| 490441000033110 | Durogesic 50micrograms transdermal patches (Janssen-Cilag Ltd) |
| 3332941000033111 | Tilofyl 50micrograms/hour transdermal patches (Tillomed Laboratories Ltd) |
| 4022841000033115 | Matrifen 50micrograms/hour transdermal patches (Teva UK Ltd) |
| 8884641000033115 | Opiodur 50micrograms/hour transdermal patches (Zentiva) |
| 4426241000033112 | Fentalis Reservoir 50micrograms/hour transdermal patches (Sandoz Ltd) |
| 575141000033112 | Fentanyl 50micrograms/hour transdermal patches |
| 4386741000033116 | Mezolar Matrix 50micrograms/hour transdermal patches (Sandoz Ltd) |
| 9204441000033114 | Mylafent 50micrograms/hour transdermal patches (Mylan) |
| 4956141000033119 | Osmanil 50micrograms/hour transdermal patches (Zentiva) |
| 4502941000033115 | Osmach 50micrograms/hour transdermal patches (Teva UK Ltd) |
| 12356141000033117 | Co-codamol 60mg/1000mg tablets |
| 1741241000033116 | Kapake Insts 60mg/1000mg effervescent powder sachets (Galen Ltd) |
| 4386841000033114 | Mezolar Matrix 75micrograms/hour transdermal patches (Sandoz Ltd) |
| 3248241000033114 | Durogesic DTrans 75micrograms/hour transdermal patches (Janssen-Cilag Ltd) |
| 4956241000033114 | Osmanil 75micrograms/hour transdermal patches (Zentiva) |
| 4426141000033117 | Fentalis Reservoir 75micrograms/hour transdermal patches (Sandoz Ltd) |
| 575241000033117 | Fentanyl 75micrograms/hour transdermal patches |
| 10597941000033116 | Yemex 75micrograms/hour transdermal patches (Sandoz Ltd) |
| 8884741000033112 | Opiodur 75micrograms/hour transdermal patches (Zentiva) |
| 4503041000033113 | Osmach 75micrograms/hour transdermal patches (Teva UK Ltd) |
| 5007241000033117 | Victanyl 75micrograms/hour transdermal patches (Accord Healthcare Ltd) |
| 4022941000033111 | Matrifen 75micrograms/hour transdermal patches (Teva UK Ltd) |
| 6441041000033119 | Fencino 75micrograms/hour transdermal patches (Ethypharm UK Ltd) |
| 490541000033111 | Durogesic 75micrograms transdermal patches (Janssen-Cilag Ltd) |
| 3332841000033115 | Tilofyl 75micrograms/hour transdermal patches (Tillomed Laboratories Ltd) |
| 9204541000033110 | Mylafent 75micrograms/hour transdermal patches (Mylan) |
| 11780841000033116 | Buplast 70micrograms/hour transdermal patches (Mylan) |
| 11577241000033114 | Prenotrix 70micrograms/hour transdermal patches (Genesis Pharmaceuticals Ltd) |
| 12603741000033113 | Carlosafine 70micrograms/hour transdermal patches (Glenmark Pharmaceuticals Europe Ltd) |
| 2737741000033115 | Buprenorphine 70micrograms/hour transdermal patches |
| 12428641000033115 | Turgeon 70micrograms/hour transdermal patches (Teva UK Ltd) |
| 12195141000033111 | Relevtec 70micrograms/hour transdermal patches (Sandoz Ltd) |
| 11484541000033117 | Bupeaze 70micrograms/hour transdermal patches (Dr Reddy's Laboratories (UK) Ltd) |
| 2738041000033119 | Transtec 70micrograms/hour transdermal patches (Napp Pharmaceuticals Ltd) |
| 13754841000033114 | Morphine sulfate 500micrograms/5ml oral solution |
| 5007341000033110 | Victanyl 100micrograms/hour transdermal patches (Accord Healthcare Ltd) |
| 574941000033113 | Fentanyl 100micrograms/hour transdermal patches |
| 490241000033114 | Durogesic 100micrograms transdermal patches (Janssen-Cilag Ltd) |
| 4956341000033116 | Osmanil 100micrograms/hour transdermal patches (Zentiva) |
| 4426041000033116 | Fentalis Reservoir 100micrograms/hour transdermal patches (Sandoz Ltd) |
| 4503141000033112 | Osmach 100micrograms/hour transdermal patches (Ratiopharm UK Ltd) |
| 8884341000033111 | Opiodur 100micrograms/hour transdermal patches (Zentiva) |
| 3333141000033119 | Tilofyl 100micrograms/hour transdermal patches (Tillomed Laboratories Ltd) |
| 6441141000033115 | Fencino 100micrograms/hour transdermal patches (Ethypharm UK Ltd) |
| 9204641000033111 | Mylafent 100micrograms/hour transdermal patches (Mylan) |
| 4386941000033118 | Mezolar Matrix 100micrograms/hour transdermal patches (Sandoz Ltd) |
| 10597841000033112 | Yemex 100micrograms/hour transdermal patches (Sandoz Ltd) |
| 4023041000033118 | Matrifen 100micrograms/hour transdermal patches (Teva UK Ltd) |
| 3248341000033116 | Durogesic DTrans 100micrograms/hour transdermal patches (Janssen-Cilag Ltd) |
| 625141000033115 | Galcodine 3mg/5ml linctus paediatric (Thornton & Ross Ltd) |
| 336041000033115 | Codeine 3mg/5ml linctus paediatric |
| 931041000033118 | Morphine Hydrochloride Mixture 916 micrograms |
| 728641000033114 | Hydromorphone 1.3mg capsules |
| 1029141000033112 | Palladone 1.3mg capsules (Napp Pharmaceuticals Ltd) |
| 1564541000033112 | Zydol SR 100mg tablets (Grunenthal Ltd) |
| 12350041000033118 | Tilodol SR 100mg tablets (Sandoz Ltd) |
| 4417741000033112 | Zeridame SR 100mg tablets (Actavis UK Ltd) |
| 1549241000033116 | Zamadol SR 100mg capsules (Mylan) |
| 930741000033113 | Morcap SR 100mg capsules (Hospira UK Ltd) |
| 1462641000033113 | Tramadol 100mg modified-release capsules |
| 930041000033110 | Morphine 100mg modified-release granules sachets sugar free |
| 1702141000033113 | Tramadol 100mg effervescent powder sachets sugar free |
| 2753641000033115 | Filnarine SR 100mg tablets (Teva UK Ltd) |
| 2078341000033117 | Dromadol SR 100mg tablets (Teva UK Ltd) |
| 12353641000033115 | Zytram SR 100mg tablets (Qdem Pharmaceuticals Ltd) |
| 6133941000033117 | Palexia SR 100mg tablets (Grunenthal Ltd) |
| 2180641000033111 | Morphine Sulfate Suppositories 100 mg |
| 2912441000033113 | Morphgesic SR 100mg tablets (Advanz Pharma) |
| 1924241000033110 | Morphine 100mg modified-release capsules |
| 940141000033110 | MST Continus Suspension 100mg granules sachets (Napp Pharmaceuticals Ltd) |
| 3996441000033116 | Tradorec XL 100mg tablets (Endo Ventures Ltd) |
| 12346941000033118 | Oldaram 100mg modified-release tablets (Ranbaxy (UK) Ltd) |
| 4824241000033110 | Marol 100mg modified-release tablets (Teva UK Ltd) |
| 4523141000033117 | Tramquel SR 100mg capsules (Mylan) |
| 3909541000033113 | Mabron 100mg modified-release tablets (Teva UK Ltd) |
| 1014041000033118 | Oramorph Sr M/R tablets 100 mg |
| 1701941000033115 | Tramake Insts 100mg sachets (Galen Ltd) |
| 4028341000033114 | Larapam SR 100mg tablets (Sandoz Ltd) |
| 4459341000033112 | Tramulief SR 100mg tablets (Advanz Pharma) |
| 941041000033119 | MST Continus 100mg tablets (Napp Pharmaceuticals Ltd) |
| 6133241000033114 | Tapentadol 100mg modified-release tablets |
| 11567841000033116 | Maneo 100mg modified-release tablets (Mylan) |
| 12346441000033111 | Invodol SR 100mg tablets (Ennogen Healthcare Ltd) |
| 936541000033119 | Morphine 100mg modified-release tablets |
| 1462341000033117 | Tramadol 100mg modified-release tablets |
| 1561241000033112 | Zomorph 100mg modified-release capsules (Ethypharm UK Ltd) |
| 4899241000033117 | Maxitram SR 100mg capsules (Chiesi Ltd) |
| 6389141000033118 | Tramadol 100mg/ml oral drops |
| 1981941000033112 | Oxycodone 10mg modified-release tablets |
| 2912141000033117 | Morphgesic SR 10mg tablets (Advanz Pharma) |
| 929741000033117 | Morphine 10mg modified-release tablets |
| 10333241000033114 | Oxeltra 10mg modified-release tablets (Wockhardt UK Ltd) |
| 1561141000033117 | Zomorph 10mg modified-release capsules (Ethypharm UK Ltd) |
| 430341000033110 | Dextromoramide 10mg tablets |
| 373141000033110 | Co-dydramol 10mg/500mg tablets |
| 3179641000033116 | Dipipanone 10mg / Cyclizine 30mg tablets |
| 470841000033115 | Diconal tablets (Amdipharm Plc) |
| 428241000033118 | Dextromoramide Suppositories 10 mg |
| 12666341000033119 | Ixyldone 10mg modified-release tablets (Morningside Healthcare Ltd) |
| 7886341000033112 | Longtec 10mg modified-release tablets (Qdem Pharmaceuticals Ltd) |
| 1041241000033118 | Palfium 10mg tablets (Roche Products Ltd) |
| 10984941000033117 | Carexil 10mg modified-release tablets (Sandoz Ltd) |
| 1987241000033111 | Oxycodone 10mg capsules |
| 11808041000033113 | Leveraxo 10mg modified-release tablets (Mylan) |
| 1013941000033115 | Oramorph Sr M/R tablets 10 mg |
| 12347241000033113 | Eroset 500mg/10mg tablets (M & A Pharmachem Ltd) |
| 12636541000033115 | Renocontin 10mg modified-release tablets (Glenmark Pharmaceuticals Europe Ltd) |
| 8537441000033117 | Lynlor 10mg capsules (Accord Healthcare Ltd) |
| 936441000033115 | Morphine 10mg tablets |
| 1923941000033116 | Morphine 10mg modified-release capsules |
| 13708541000033112 | Oxycodone 10mg tablets |
| 940941000033112 | MST Continus 10mg tablets (Napp Pharmaceuticals Ltd) |
| 1403141000033111 | Syndol caplets (Sanofi) |
| 9177341000033118 | Reltebon 10mg modified-release tablets (Accord Healthcare Ltd) |
| 1278541000033116 | Sevredol 10mg tablets (Napp Pharmaceuticals Ltd) |
| 12664541000033119 | Oxypro 10mg modified-release tablets (Ridge Pharma Ltd) |
| 1273741000033117 | Sevredol Suppositories 10 mg |
| 10641541000033118 | Abtard 10mg modified-release tablets (Ethypharm UK Ltd) |
| 1987641000033114 | OxyContin 10mg modified-release tablets (Napp Pharmaceuticals Ltd) |
| 1988141000033117 | OxyNorm 10mg capsules (Napp Pharmaceuticals Ltd) |
| 1039441000033119 | Palfium Suppositories 10 mg |
| 1041941000033110 | Papaveretum Tablets 10 mg |
| 2753341000033111 | Filnarine SR 10mg tablets (Teva UK Ltd) |
| 9061641000033115 | Shortec 10mg capsules (Qdem Pharmaceuticals Ltd) |
| 12185341000033113 | Onexila XL 10mg tablets (Aspire Pharma Ltd) |
| 8048141000033110 | Oxylan 10mg modified-release tablets (Healthcare Pharma Ltd) |
| 934141000033117 | Morphine sulfate 10mg suppositories |
| 11243241000033114 | Zomestine 10mg modified-release tablets (Accord Healthcare Ltd) |
| 1140541000033115 | Propain caplets (Ceuta Healthcare Ltd) |
| 4898341000033115 | Targinact 20mg/10mg modified-release tablets (Napp Pharmaceuticals Ltd) |
| 4898241000033113 | Oxycodone 20mg / Naloxone 10mg modified-release tablets |
| 1988341000033119 | OxyNorm 10mg/ml concentrate oral solution (Napp Pharmaceuticals Ltd) |
| 12187141000033118 | Shortec 10mg/ml concentrate oral solution (Qdem Pharmaceuticals Ltd) |
| 13118741000033113 | Morphine (Opium tincture) 10mg/ml oral drops sugar free |
| 1987541000033113 | Oxycodone 10mg/ml oral solution sugar free |
| 2968841000033110 | Panadol Ultra 12.8mg/500mg tablets (GlaxoSmithKline Consumer Healthcare) |
| 2968641000033114 | Solpaflex tablets (GlaxoSmithKline Consumer Healthcare) |
| 2228341000033113 | Nurofen Plus tablets (Reckitt Benckiser Healthcare (UK) Ltd) |
| 12708841000033116 | Ibuprofen 200mg / Codeine 12.8mg tablets |
| 3077941000033110 | Cuprofen PLUS tablets (SSL International Plc) |
| 12583641000033114 | Solpadeine Max soluble tablets (Omega Pharma Ltd) |
| 5334541000033117 | Co-codamol 12.8mg/500mg tablets |
| 9292941000033111 | Solpadeine Max 12.8mg/500mg tablets (Omega Pharma Ltd) |
| 433441000033114 | DHC Continus 120mg tablets (Napp Pharmaceuticals Ltd) |
| 10492241000033118 | Longtec 120mg modified-release tablets (Qdem Pharmaceuticals Ltd) |
| 2068541000033117 | Morphine 120mg modified-release capsules |
| 6125341000033115 | Oxycodone 120mg modified-release tablets |
| 469441000033116 | Dihydrocodeine 120mg modified-release tablets |
| 944041000033117 | MXL 120mg capsules (Napp Pharmaceuticals Ltd) |
| 6125741000033119 | OxyContin 120mg modified-release tablets (Napp Pharmaceuticals Ltd) |
| 2068641000033116 | Morphine 150mg modified-release capsules |
| 4417641000033115 | Zeridame SR 150mg tablets (Actavis UK Ltd) |
| 4028541000033119 | Larapam SR 150mg tablets (Sandoz Ltd) |
| 4523241000033112 | Tramquel SR 150mg capsules (Mylan) |
| 1462441000033111 | Tramadol 150mg modified-release tablets |
| 4459441000033118 | Tramulief SR 150mg tablets (Advanz Pharma) |
| 12353741000033112 | Zytram SR 150mg tablets (Qdem Pharmaceuticals Ltd) |
| 6134041000033115 | Palexia SR 150mg tablets (Grunenthal Ltd) |
| 944141000033118 | MXL 150mg capsules (Napp Pharmaceuticals Ltd) |
| 1850141000033116 | Zydol XL 150mg tablets (Grunenthal Ltd) |
| 4824341000033117 | Marol 150mg modified-release tablets (Teva UK Ltd) |
| 6133341000033116 | Tapentadol 150mg modified-release tablets |
| 11568141000033114 | Maneo 150mg modified-release tablets (Mylan) |
| 12346641000033113 | Invodol SR 150mg tablets (Ennogen Healthcare Ltd) |
| 3909641000033114 | Mabron 150mg modified-release tablets (Teva UK Ltd) |
| 12350141000033119 | Tilodol SR 150mg tablets (Sandoz Ltd) |
| 1462741000033116 | Tramadol 150mg modified-release capsules |
| 3344141000033117 | Zamadol 24hr 150mg modified-release tablets (Mylan) |
| 1564641000033113 | Zydol SR 150mg tablets (Grunenthal Ltd) |
| 2183441000033116 | Dromadol XL 150mg tablets (IVAX Pharmaceuticals UK Ltd) |
| 1549341000033114 | Zamadol SR 150mg capsules (Mylan) |
| 2078441000033111 | Dromadol SR 150mg tablets (Teva UK Ltd) |
| 4899341000033110 | Maxitram SR 150mg capsules (Chiesi Ltd) |
| 10333341000033116 | Oxeltra 15mg modified-release tablets (Wockhardt UK Ltd) |
| 12636741000033111 | Renocontin 15mg modified-release tablets (Glenmark Pharmaceuticals Europe Ltd) |
| 6125441000033114 | OxyContin 15mg modified-release tablets (Napp Pharmaceuticals Ltd) |
| 6125041000033117 | Oxycodone 15mg modified-release tablets |
| 10491541000033114 | Longtec 15mg modified-release tablets (Qdem Pharmaceuticals Ltd) |
| 929941000033119 | Morphine 15mg modified-release tablets |
| 934641000033110 | Morphine hydrochloride 15mg suppositories |
| 12664641000033118 | Oxypro 15mg modified-release tablets (Ridge Pharma Ltd) |
| 940041000033111 | MST Continus 15mg tablets (Napp Pharmaceuticals Ltd) |
| 6431341000033116 | Co-codamol 15mg/500mg capsules |
| 9809641000033116 | Reltebon 15mg modified-release tablets (Accord Healthcare Ltd) |
| 933841000033114 | Morphine sulfate 15mg suppositories |
| 371141000033111 | Codeine 15mg tablets |
| 10641641000033117 | Abtard 15mg modified-release tablets (Ethypharm UK Ltd) |
| 6431441000033110 | Codipar 15mg/500mg capsules (Advanz Pharma) |
| 2850041000033116 | Codipar 15mg/500mg tablets (Advanz Pharma) |
| 2875341000033110 | Co-codamol 15mg/500mg tablets |
| 6137941000033114 | Kapake 15mg/500mg tablets (Galen Ltd) |
| 6386441000033115 | Co-codamol 15mg/500mg effervescent tablets sugar free |
| 6386541000033119 | Codipar 15mg/500mg effervescent tablets (Advanz Pharma) |
| 1036641000033110 | Palladone SR 16mg capsules (Napp Pharmaceuticals Ltd) |
| 738241000033114 | Hydromorphone 16mg modified-release capsules |
| 2093241000033114 | Morphine sulfate powder |
| 2086141000033111 | Codeine phosphate powder |
| 2093141000033119 | Morphine hydrochloride powder |
| 4434241000033119 | Morphine sulfate 5mg/5ml oral solution |
| 1987441000033112 | Oxycodone 5mg/5ml oral solution sugar free |
| 11444741000033116 | Morphine 0.1% in Intrasite gel |
| 12187041000033117 | Shortec liquid 1mg/ml oral solution (Qdem Pharmaceuticals Ltd) |
| 1988441000033113 | OxyNorm liquid 1mg/ml oral solution (Napp Pharmaceuticals Ltd) |
| 5234441000033111 | Targinact 5mg/2.5mg modified-release tablets (Napp Pharmaceuticals Ltd) |
| 5234241000033110 | Oxycodone 5mg / Naloxone 2.5mg modified-release tablets |
| 1029241000033117 | Palladone 2.6mg capsules (Napp Pharmaceuticals Ltd) |
| 728741000033117 | Hydromorphone 2.6mg capsules |
| 140841000033118 | Benylin with Codeine oral solution (Pfizer Consumer Healthcare Ltd) |
| 940241000033115 | MST Continus Suspension 200mg granules sachets (Napp Pharmaceuticals Ltd) |
| 3909741000033117 | Mabron 200mg modified-release tablets (Teva UK Ltd) |
| 4028641000033118 | Larapam SR 200mg tablets (Sandoz Ltd) |
| 4459541000033117 | Tramulief SR 200mg tablets (Advanz Pharma) |
| 12346741000033116 | Invodol SR 200mg tablets (Ennogen Healthcare Ltd) |
| 1462541000033112 | Tramadol 200mg modified-release tablets |
| 4817241000033113 | Filnarine SR 200mg tablets (Teva UK Ltd) |
| 941341000033117 | MST Continus 200mg tablets (Napp Pharmaceuticals Ltd) |
| 1549441000033115 | Zamadol SR 200mg capsules (Mylan) |
| 896441000033119 | Meptid 200mg tablets (Almirall Ltd) |
| 1561341000033119 | Zomorph 200mg modified-release capsules (Ethypharm UK Ltd) |
| 896341000033113 | Meptazinol 200mg tablets |
| 933141000033115 | Morphine 200mg modified-release tablets |
| 944241000033113 | MXL 200mg capsules (Napp Pharmaceuticals Ltd) |
| 3996541000033115 | Tradorec XL 200mg tablets (Endo Ventures Ltd) |
| 6133441000033110 | Tapentadol 200mg modified-release tablets |
| 1564741000033116 | Zydol SR 200mg tablets (Grunenthal Ltd) |
| 6134141000033116 | Palexia SR 200mg tablets (Grunenthal Ltd) |
| 11568441000033118 | Maneo 200mg modified-release tablets (Mylan) |
| 2078541000033112 | Dromadol SR 200mg tablets (Teva UK Ltd) |
| 1850241000033111 | Zydol XL 200mg tablets (Grunenthal Ltd) |
| 13712041000033119 | Brimisol PR 200mg tablets (Bristol Laboratories Ltd) |
| 4899441000033116 | Maxitram SR 200mg capsules (Chiesi Ltd) |
| 4824441000033111 | Marol 200mg modified-release tablets (Teva UK Ltd) |
| 1462841000033114 | Tramadol 200mg modified-release capsules |
| 1924341000033117 | Morphine 200mg modified-release capsules |
| 4417541000033116 | Zeridame SR 200mg tablets (Actavis UK Ltd) |
| 930141000033114 | Morphine 200mg modified-release granules sachets sugar free |
| 12350241000033114 | Tilodol SR 200mg tablets (Sandoz Ltd) |
| 12353841000033119 | Zytram SR 200mg tablets (Qdem Pharmaceuticals Ltd) |
| 4523341000033119 | Tramquel SR 200mg capsules (Mylan) |
| 2183541000033115 | Dromadol XL 200mg tablets (IVAX Pharmaceuticals UK Ltd) |
| 3344241000033112 | Zamadol 24hr 200mg modified-release tablets (Mylan) |
| 9061741000033112 | Shortec 20mg capsules (Qdem Pharmaceuticals Ltd) |
| 12664741000033110 | Oxypro 20mg modified-release tablets (Ridge Pharma Ltd) |
| 12666541000033114 | Ixyldone 20mg modified-release tablets (Morningside Healthcare Ltd) |
| 8048241000033115 | Oxylan 20mg modified-release tablets (Healthcare Pharma Ltd) |
| 1987741000033117 | OxyContin 20mg modified-release tablets (Napp Pharmaceuticals Ltd) |
| 9177241000033111 | Reltebon 20mg modified-release tablets (Accord Healthcare Ltd) |
| 1982041000033118 | Oxycodone 20mg modified-release tablets |
| 11808141000033112 | Leveraxo 20mg modified-release tablets (Mylan) |
| 940741000033114 | MST Continus Suspension 20mg granules sachets (Napp Pharmaceuticals Ltd) |
| 939541000033110 | Morphine 20mg tablets |
| 7886441000033118 | Longtec 20mg modified-release tablets (Qdem Pharmaceuticals Ltd) |
| 2068741000033113 | Morphine 20mg modified-release capsules |
| 934241000033112 | Morphine sulfate 20mg suppositories |
| 1987341000033118 | Oxycodone 20mg capsules |
| 10333441000033110 | Oxeltra 20mg modified-release tablets (Wockhardt UK Ltd) |
| 3032841000033119 | Morphine 20mg modified-release granules sachets sugar free |
| 930841000033115 | Morcap SR 20mg capsules (Hospira UK Ltd) |
| 336741000033117 | Codafen Continus tablets (Napp Pharmaceuticals Ltd) |
| 10985141000033118 | Carexil 20mg modified-release tablets (Sandoz Ltd) |
| 13708641000033113 | Oxycodone 20mg tablets |
| 8537541000033116 | Lynlor 20mg capsules (Accord Healthcare Ltd) |
| 11243341000033116 | Zomestine 20mg modified-release tablets (Accord Healthcare Ltd) |
| 12185541000033118 | Onexila XL 20mg tablets (Aspire Pharma Ltd) |
| 1278641000033115 | Sevredol 20mg tablets (Napp Pharmaceuticals Ltd) |
| 1988241000033112 | OxyNorm 20mg capsules (Napp Pharmaceuticals Ltd) |
| 10641741000033114 | Abtard 20mg modified-release tablets (Ethypharm UK Ltd) |
| 1273841000033110 | Sevredol Suppositories 20 mg |
| 12636941000033114 | Renocontin 20mg modified-release tablets (Glenmark Pharmaceuticals Europe Ltd) |
| 7859041000033111 | Dypracet 20mg/500mg tablets (Auden McKenzie (Pharma Division) Ltd) |
| 3229141000033111 | Ibuprofen 300mg modified-release / Codeine 20mg tablets |
| 3180241000033119 | Co-dydramol 20mg/500mg tablets |
| 1164541000033115 | Remedeine tablets (Crescent Pharma Ltd) |
| 5234541000033112 | Targinact 40mg/20mg modified-release tablets (Napp Pharmaceuticals Ltd) |
| 5234341000033117 | Oxycodone 40mg / Naloxone 20mg modified-release tablets |
| 1014941000033117 | Oramorph 20mg/ml concentrated oral solution (Boehringer Ingelheim Ltd) |
| 1744741000033112 | Morphine Sulfate Concentrated Oral Solution 20 mg/ml |
| 931841000033113 | Morphine sulfate 100mg/5ml oral solution unit dose vials sugar free |
| 1752041000033116 | Sevredol 20mg/ml concentrated oral solution (Napp Pharmaceuticals Ltd) |
| 931641000033112 | Morphine sulfate 20mg/ml oral solution sugar free |
| 9160141000033113 | Tapentadol 20mg/ml oral solution sugar free |
| 1014541000033111 | Oramorph 100mg/5ml oral solution unit dose vials (Boehringer Ingelheim Ltd) |
| 9160241000033118 | Palexia 20mg/ml oral solution (Grunenthal Ltd) |
| 738441000033110 | Hydromorphone 24mg modified-release capsules |
| 1036841000033111 | Palladone SR 24mg capsules (Napp Pharmaceuticals Ltd) |
| 6134241000033111 | Palexia SR 250mg tablets (Grunenthal Ltd) |
| 6133541000033111 | Tapentadol 250mg modified-release tablets |
| 609341000033114 | Fortral 25mg tablets (Zentiva) |
| 1066841000033118 | Pethidine Hydrochloride Tablets 25 mg |
| 1068041000033114 | Pentazocine 25mg tablets |
| 1036741000033118 | Palladone SR 2mg capsules (Napp Pharmaceuticals Ltd) |
| 11789141000033119 | Buprenorphine 2mg oral lyophilisates sugar free |
| 738341000033116 | Hydromorphone 2mg modified-release capsules |
| 11789341000033116 | Espranor 2mg oral lyophilisates (Martindale Pharmaceuticals Ltd) |
| 1014841000033113 | Oramorph 10mg/5ml oral solution (Boehringer Ingelheim Ltd) |
| 5891241000033112 | Co-dydramol 10mg/500mg/5ml oral solution |
| 1752541000033114 | Sevredol 10mg/5ml oral solution (Napp Pharmaceuticals Ltd) |
| 931741000033115 | Morphine sulfate 10mg/5ml oral solution unit dose vials sugar free |
| 13582941000033117 | Dihydrocodeine 10mg/5ml oral suspension |
| 442841000033115 | Dihydrocodeine 10mg/5ml oral solution |
| 3851441000033111 | Co-dydramol 10mg/500mg/5ml oral suspension |
| 1014441000033110 | Oramorph 10mg/5ml oral solution unit dose vials (Boehringer Ingelheim Ltd) |
| 10044141000033116 | Morphine 0.2% in Intrasite gel |
| 7859941000033112 | Morphine hydrochloride 10mg/5ml oral solution |
| 931541000033111 | Morphine sulfate 10mg/5ml oral solution |
| 3344341000033119 | Zamadol 24hr 300mg modified-release tablets (Mylan) |
| 3996641000033119 | Tradorec XL 300mg tablets (Endo Ventures Ltd) |
| 2183641000033119 | Dromadol XL 300mg tablets (IVAX Pharmaceuticals UK Ltd) |
| 1849941000033112 | Tramadol 300mg modified-release tablets |
| 1850341000033118 | Zydol XL 300mg tablets (Grunenthal Ltd) |
| 1025241000033119 | Oxycodone Pectinate Suppositories 30 mg |
| 934341000033119 | Morphine Hydrochloride Suppositories 30 mg |
| 295341000033111 | Co-Codamol 30/500 Caplets |
| 10641841000033116 | Abtard 30mg modified-release tablets (Ethypharm UK Ltd) |
| 6125141000033118 | Oxycodone 30mg modified-release tablets |
| 1014141000033119 | Oramorph Sr M/R tablets 30 mg |
| 371241000033116 | Codeine 30mg tablets |
| 941141000033115 | MST Continus 30mg tablets (Napp Pharmaceuticals Ltd) |
| 2753441000033117 | Filnarine SR 30mg tablets (Teva UK Ltd) |
| 6125541000033110 | OxyContin 30mg modified-release tablets (Napp Pharmaceuticals Ltd) |
| 9809741000033113 | Reltebon 30mg modified-release tablets (Accord Healthcare Ltd) |
| 468641000033111 | Dihydrocodeine Tartrate Tablets 30 mg |
| 944341000033115 | MXL 30mg capsules (Napp Pharmaceuticals Ltd) |
| 2912241000033112 | Morphgesic SR 30mg tablets (Advanz Pharma) |
| 10491641000033110 | Longtec 30mg modified-release tablets (Qdem Pharmaceuticals Ltd) |
| 1924041000033119 | Morphine 30mg modified-release capsules |
| 3032941000033110 | Morphine 30mg modified-release granules sachets sugar free |
| 12637241000033119 | Renocontin 30mg modified-release tablets (Glenmark Pharmaceuticals Europe Ltd) |
| 12664841000033117 | Oxypro 30mg modified-release tablets (Ridge Pharma Ltd) |
| 10333541000033111 | Oxeltra 30mg modified-release tablets (Wockhardt UK Ltd) |
| 940841000033116 | MST Continus Suspension 30mg granules sachets (Napp Pharmaceuticals Ltd) |
| 1561441000033113 | Zomorph 30mg modified-release capsules (Ethypharm UK Ltd) |
| 12666641000033110 | Ixyldone 30mg modified-release tablets (Morningside Healthcare Ltd) |
| 936641000033118 | Morphine 30mg modified-release tablets |
| 11808241000033117 | Leveraxo 30mg modified-release tablets (Mylan) |
| 1273941000033119 | Sevredol Suppositories 30 mg |
| 933941000033118 | Morphine sulfate 30mg suppositories |
| 468541000033110 | Dihydrocodeine 30mg tablets |
| 1698041000033118 | Solpadol 30mg/500mg capsules (Sanofi) |
| 12684741000033112 | Emcozin 30mg/500mg tablets (M & A Pharmachem Ltd) |
| 1352541000033110 | Solpadol 30mg/500mg effervescent tablets (Sanofi) |
| 295441000033117 | Co-codamol 30mg/500mg capsules |
| 1621841000033112 | Kapake Insts 30mg/500mg effervescent powder sachets (Galen Ltd) |
| 796641000033119 | Kapake 30mg/500mg tablets (Galen Ltd) |
| 1830841000033114 | Kapake 30mg/500mg capsules (Galen Ltd) |
| 1588741000033112 | Co-codamol 30mg/500mg effervescent powder sachets sugar free |
| 2746141000033115 | Zapain 30mg/500mg tablets (Advanz Pharma) |
| 3057541000033118 | Kapake 30mg/500mg effervescent tablets (Galen Ltd) |
| 1363841000033114 | Solpadol 30mg/500mg caplets (Sanofi) |
| 3180141000033114 | Co-dydramol 30mg/500mg tablets |
| 1479641000033111 | Tylex 30mg/500mg capsules (UCB Pharma Ltd) |
| 3331541000033110 | Medocodene 30mg/500mg effervescent tablets (Mylan) |
| 3334541000033113 | Medocodene 30mg/500mg capsules (UCB Pharma Ltd) |
| 1479741000033119 | Tylex 30mg/500mg effervescent tablets (UCB Pharma Ltd) |
| 2745941000033112 | Zapain 30mg/500mg capsules (Advanz Pharma) |
| 1157541000033117 | Remedeine Forte tablets (Crescent Pharma Ltd) |
| 7859141000033110 | Dypracet 30mg/500mg tablets (Auden McKenzie (Pharma Division) Ltd) |
| 326141000033118 | Co-codamol 30mg/500mg effervescent tablets |
| 370641000033114 | Co-codamol 30mg/500mg tablets |
| 468141000033118 | Distalgesic 32.5mg/325mg tablets (Meda Pharmaceuticals Ltd) |
| 373241000033115 | Co-proxamol 32.5mg/325mg tablets |
| 3180341000033112 | Tramadol 37.5mg / Paracetamol 325mg tablets |
| 5595941000033118 | Tramadol 37.5mg / Paracetamol 325mg effervescent tablets sugar free |
| 5596041000033111 | Tramacet 37.5mg/325mg effervescent tablets (Grunenthal Ltd) |
| 3057641000033117 | Tramacet 37.5mg/325mg tablets (Grunenthal Ltd) |
| 2645041000033118 | Codeine 15mg/5ml linctus sugar free |
| 335841000033117 | Codeine Phosphate Linctus 15 mg/5 ml |
| 368441000033116 | Codeine 15mg/5ml linctus |
| 831641000033117 | Linctus Of Codeine Linctus 15 mg/5 ml |
| 624441000033112 | Galcodine 15mg/5ml linctus (Thornton & Ross Ltd) |
| 335641000033118 | Codeine Linctus Diabetic Linctus 15 mg/5 ml |
| 1850441000033112 | Zydol XL 400mg tablets (Grunenthal Ltd) |
| 2183741000033111 | Dromadol XL 400mg tablets (IVAX Pharmaceuticals UK Ltd) |
| 1850041000033115 | Tramadol 400mg modified-release tablets |
| 3344441000033113 | Zamadol 24hr 400mg modified-release tablets (Mylan) |
| 1987841000033110 | OxyContin 40mg modified-release tablets (Napp Pharmaceuticals Ltd) |
| 433141000033118 | DF 118 Forte 40mg tablets (Martindale Pharmaceuticals Ltd) |
| 9177141000033116 | Reltebon 40mg modified-release tablets (Accord Healthcare Ltd) |
| 11808341000033110 | Leveraxo 40mg modified-release tablets (Mylan) |
| 462841000033114 | Dihydrocodeine 40mg tablets |
| 11243441000033110 | Zomestine 40mg modified-release tablets (Accord Healthcare Ltd) |
| 12666741000033118 | Ixyldone 40mg modified-release tablets (Morningside Healthcare Ltd) |
| 7886541000033117 | Longtec 40mg modified-release tablets (Qdem Pharmaceuticals Ltd) |
| 12185641000033117 | Onexila XL 40mg tablets (Aspire Pharma Ltd) |
| 10333641000033112 | Oxeltra 40mg modified-release tablets (Wockhardt UK Ltd) |
| 8048341000033113 | Oxylan 40mg modified-release tablets (Healthcare Pharma Ltd) |
| 12664941000033113 | Oxypro 40mg modified-release tablets (Ridge Pharma Ltd) |
| 1982141000033119 | Oxycodone 40mg modified-release tablets |
| 12637341000033112 | Renocontin 40mg modified-release tablets (Glenmark Pharmaceuticals Europe Ltd) |
| 10641941000033112 | Abtard 40mg modified-release tablets (Ethypharm UK Ltd) |
| 1036941000033115 | Palladone SR 4mg capsules (Napp Pharmaceuticals Ltd) |
| 738541000033111 | Hydromorphone 4mg modified-release capsules |
| 2068841000033115 | Morphine 50mg modified-release capsules |
| 4523041000033116 | Tramquel SR 50mg capsules (Mylan) |
| 1702241000033118 | Tramadol 50mg effervescent powder sachets sugar free |
| 1564341000033117 | Zydol 50mg capsules (Grunenthal Ltd) |
| 1276641000033119 | Sevredol 50mg tablets (Napp Pharmaceuticals Ltd) |
| 13300641000033110 | Pethidine 50mg capsules |
| 1063141000033116 | Pentazocine 50mg suppositories |
| 1454241000033114 | Tramadol 50mg capsules |
| 2980441000033112 | Zamadol Melt 50mg tablets (Mylan) |
| 6132041000033110 | Tapentadol 50mg tablets |
| 607341000033118 | Fortral 50mg suppositories (Sterwin Medicines) |
| 1065641000033110 | Pethidine 50mg tablets |
| 2980341000033118 | Tramadol 50mg orodispersible tablets sugar free |
| 1454541000033111 | Tramake 50mg capsules (Galen Ltd) |
| 2180441000033114 | Morphine Sulfate Suppositories 50 mg |
| 4259241000033118 | Zydol SR 50mg tablets (Grunenthal Ltd) |
| 938241000033117 | Morphine 50mg tablets |
| 1564841000033114 | Zydol 50mg soluble tablets (Grunenthal Ltd) |
| 6132241000033119 | Palexia 50mg tablets (Grunenthal Ltd) |
| 930941000033111 | Morcap SR 50mg capsules (Hospira UK Ltd) |
| 1549541000033119 | Zamadol SR 50mg capsules (Mylan) |
| 599141000033118 | Fortral Capsules 50 mg |
| 6133841000033113 | Palexia SR 50mg tablets (Grunenthal Ltd) |
| 1548041000033116 | Zamadol 50mg capsules (Mylan) |
| 1702041000033114 | Tramake Insts 50mg sachets (Galen Ltd) |
| 1044641000033117 | Pentazocine 50mg capsules |
| 4259141000033113 | Tramadol 50mg modified-release tablets |
| 4899141000033112 | Maxitram SR 50mg capsules (Chiesi Ltd) |
| 1465741000033113 | Tramadol 50mg soluble tablets sugar free |
| 6133141000033119 | Tapentadol 50mg modified-release tablets |
| 1462941000033118 | Tramadol 50mg modified-release capsules |
| 8048041000033111 | Oxylan 5mg modified-release tablets (Healthcare Pharma Ltd) |
| 13708741000033116 | Oxyact 5mg tablets (Kent Pharmaceuticals Ltd) |
| 2748441000033110 | Oxycodone 5mg modified-release tablets |
| 11808441000033116 | Leveraxo 5mg modified-release tablets (Mylan) |
| 430541000033115 | Dextromoramide 5mg tablets |
| 10333141000033119 | Oxeltra 5mg modified-release tablets (Wockhardt UK Ltd) |
| 12636441000033116 | Renocontin 5mg modified-release tablets (Glenmark Pharmaceuticals Europe Ltd) |
| 2748541000033111 | OxyContin 5mg modified-release tablets (Napp Pharmaceuticals Ltd) |
| 7886241000033119 | Longtec 5mg modified-release tablets (Qdem Pharmaceuticals Ltd) |
| 12666841000033111 | Ixyldone 5mg modified-release tablets (Morningside Healthcare Ltd) |
| 1987141000033116 | Oxycodone 5mg capsules |
| 12665041000033113 | Oxypro 5mg modified-release tablets (Ridge Pharma Ltd) |
| 13708441000033111 | Oxycodone 5mg tablets |
| 1041341000033111 | Palfium 5mg tablets (Roche Products Ltd) |
| 8537341000033111 | Lynlor 5mg capsules (Accord Healthcare Ltd) |
| 9177041000033115 | Reltebon 5mg modified-release tablets (Accord Healthcare Ltd) |
| 1988041000033116 | OxyNorm 5mg capsules (Napp Pharmaceuticals Ltd) |
| 940441000033119 | MST Continus 5mg tablets (Napp Pharmaceuticals Ltd) |
| 11243141000033119 | Zomestine 5mg modified-release tablets (Accord Healthcare Ltd) |
| 10984841000033113 | Carexil 5mg modified-release tablets (Sandoz Ltd) |
| 930341000033112 | Morphine 5mg modified-release tablets |
| 10641441000033119 | Abtard 5mg modified-release tablets (Ethypharm UK Ltd) |
| 9061541000033116 | Shortec 5mg capsules (Qdem Pharmaceuticals Ltd) |
| 4898141000033118 | Oxycodone 10mg / Naloxone 5mg modified-release tablets |
| 4898441000033114 | Targinact 10mg/5mg modified-release tablets (Napp Pharmaceuticals Ltd) |
| 13576541000033116 | Myloxifin 10mg/5mg modified-release tablets (Zentiva) |
| 368541000033115 | Codeine 25mg/5ml oral solution |
| 2972441000033117 | Co-proxamol 32.5mg/325mg/5ml oral suspension |
| 944441000033114 | MXL 60mg capsules (Napp Pharmaceuticals Ltd) |
| 9809841000033115 | Reltebon 60mg modified-release tablets (Accord Healthcare Ltd) |
| 10492041000033114 | Longtec 60mg modified-release tablets (Qdem Pharmaceuticals Ltd) |
| 10642041000033118 | Abtard 60mg modified-release tablets (Ethypharm UK Ltd) |
| 940341000033113 | MST Continus Suspension 60mg granules sachets (Napp Pharmaceuticals Ltd) |
| 1561541000033114 | Zomorph 60mg modified-release capsules (Ethypharm UK Ltd) |
| 2912341000033119 | Morphgesic SR 60mg tablets (Advanz Pharma) |
| 941241000033110 | MST Continus 60mg tablets (Napp Pharmaceuticals Ltd) |
| 6125641000033111 | OxyContin 60mg modified-release tablets (Napp Pharmaceuticals Ltd) |
| 1924141000033115 | Morphine 60mg modified-release capsules |
| 930241000033119 | Morphine 60mg modified-release granules sachets sugar free |
| 936741000033110 | Morphine 60mg modified-release tablets |
| 11808541000033115 | Leveraxo 60mg modified-release tablets (Mylan) |
| 468741000033119 | Dihydrocodeine 60mg modified-release tablets |
| 416141000033118 | Dextropropoxyphene 60mg capsules |
| 371341000033114 | Codeine 60mg tablets |
| 6125241000033113 | Oxycodone 60mg modified-release tablets |
| 2753541000033116 | Filnarine SR 60mg tablets (Teva UK Ltd) |
| 12637441000033118 | Renocontin 60mg modified-release tablets (Glenmark Pharmaceuticals Europe Ltd) |
| 1014241000033114 | Oramorph Sr M/R tablets 60 mg |
| 10333741000033115 | Oxeltra 60mg modified-release tablets (Wockhardt UK Ltd) |
| 433341000033115 | DHC Continus 60mg tablets (Napp Pharmaceuticals Ltd) |
| 12665141000033112 | Oxypro 60mg modified-release tablets (Ridge Pharma Ltd) |
| 472441000033119 | Doloxene Capsules 65 mg |
| 13417741000033113 | Co-codamol 30mg/500mg/5ml oral solution sugar free |
| 931941000033117 | Morphine sulfate 30mg/5ml oral solution unit dose vials sugar free |
| 1014641000033112 | Oramorph 30mg/5ml oral solution unit dose vials (Boehringer Ingelheim Ltd) |
| 1043041000033113 | Paramol tablets (SSL International Plc) |
| 86841000033118 | Aspav dispersible tablets (Actavis UK Ltd) |
| 3849841000033110 | Aspirin 500mg / Papaveretum 7.71mg dispersible tablets sugar free |
| 12471541000033111 | Tramadol 75mg / Dexketoprofen 25mg tablets |
| 6132341000033112 | Palexia 75mg tablets (Grunenthal Ltd) |
| 2078141000033115 | Tramadol 75mg modified-release tablets |
| 6132141000033114 | Tapentadol 75mg tablets |
| 2078241000033110 | Dromadol SR 75mg tablets (IVAX Pharmaceuticals UK Ltd) |
| 12353941000033110 | Zytram SR 75mg tablets (Qdem Pharmaceuticals Ltd) |
| 11783041000033115 | Tramadol 75mg / Paracetamol 650mg tablets |
| 932041000033111 | Morphine Oral solution 8.4 mg/ml |
| 11243541000033111 | Zomestine 80mg modified-release tablets (Accord Healthcare Ltd) |
| 10642141000033119 | Abtard 80mg modified-release tablets (Ethypharm UK Ltd) |
| 7886641000033116 | Longtec 80mg modified-release tablets (Qdem Pharmaceuticals Ltd) |
| 12185741000033114 | Onexila XL 80mg tablets (Aspire Pharma Ltd) |
| 12665241000033117 | Oxypro 80mg modified-release tablets (Ridge Pharma Ltd) |
| 12667041000033119 | Ixyldone 80mg modified-release tablets (Morningside Healthcare Ltd) |
| 1987941000033119 | OxyContin 80mg modified-release tablets (Napp Pharmaceuticals Ltd) |
| 10333841000033113 | Oxeltra 80mg modified-release tablets (Wockhardt UK Ltd) |
| 11808641000033119 | Leveraxo 80mg modified-release tablets (Mylan) |
| 1982541000033111 | Oxycodone 80mg modified-release tablets |
| 9176841000033112 | Reltebon 80mg modified-release tablets (Accord Healthcare Ltd) |
| 8048441000033119 | Oxylan 80mg modified-release tablets (Healthcare Pharma Ltd) |
| 1365241000033111 | Solpadeine Tablets |
| 11789241000033114 | Buprenorphine 8mg oral lyophilisates sugar free |
| 3934441000033118 | Codis 500 dispersible tablets (Reckitt Benckiser Healthcare (UK) Ltd) |
| 738641000033112 | Hydromorphone 8mg modified-release capsules |
| 918241000033114 | Migraleve tablets (McNeil Products Ltd) |
| 318141000033113 | Codis Dispersible tablets |
| 1512141000033114 | Veganin tablets (Omega Pharma Ltd) |
| 916741000033116 | Migraleve Pink tablets (McNeil Products Ltd) |
| 86941000033114 | Aspirin Paracetamol And Codeine Tablets |
| 1037041000033119 | Palladone SR 8mg capsules (Napp Pharmaceuticals Ltd) |
| 1040041000033117 | Paracetamol & Codeine Tablets |
| 371041000033112 | Co-codaprin 8mg/400mg dispersible tablets |
| 4590641000033113 | Co-codamol 8mg/500mg caplets (Vantage) |
| 11789441000033110 | Espranor 8mg oral lyophilisates (Martindale Pharmaceuticals Ltd) |
| 8246141000033119 | Solpadeine Plus capsules (Omega Pharma Ltd) |
| 4432041000033115 | Paracodol 8mg/500mg capsules (Bayer Plc) |
| 1030141000033119 | Paracodol 8mg/500mg effervescent tablets (Bayer Plc) |
| 370941000033119 | Co-codaprin 8mg/400mg tablets |
| 3934341000033112 | Aspirin 500mg / Codeine 8mg dispersible tablets sugar free |
| 294841000033118 | Co-codamol 8mg/500mg capsules |
| 922541000033116 | Migraleve Yellow tablets (McNeil Products Ltd) |
| 372941000033118 | Co-codamol 8mg/500mg tablets |
| 11736241000033114 | Solpadeine Plus soluble tablets (Omega Pharma Ltd) |
| 373041000033111 | Co-codamol 8mg/500mg effervescent tablets |
| 13497541000033118 | Co-codamol 8mg/500mg effervescent tablets sugar free |
| 433541000033110 | DHC Continus 90mg tablets (Napp Pharmaceuticals Ltd) |
| 944541000033110 | MXL 90mg capsules (Napp Pharmaceuticals Ltd) |
| 469541000033115 | Dihydrocodeine 90mg modified-release tablets |
| 2068441000033118 | Morphine 90mg modified-release capsules |

### **Supplementary Table S3. Comparison of results of mixed effects logistic regression, Cox’s competing risk regression and time-dependent Cox regression**

|  | Mixed-effect logistic regression | Cox’s competing risk model | Time dependent Cox regression |
| --- | --- | --- | --- |
| **Factors** | **aHR (95%CI)** | **aSHR (95%CI)** | **aHR (95%CI)** |
| **Age** | 0.99 (0.99 to 0.99) * | 0.99 (0.99 to 1.00) * | 0.99 (0.99 to 1.00) * |
| **Sex - Male** | 1.01 (0.98 to 1.03) | 0.98 (0.98 to 0.99) * | 0.98 (0.95 to 1.00) |
| **Ethnicity** |  |  |  |
| White (Ref.) |  |  |  |
| Asian | 1.31 (1.23 to 1.39) * | 1.12 (1.10 to 1.14) * | 1.31 (1.24 to 1.39) * |
| Black | 1.21 (1.13 to 1.31) * | 1.10 (1.08 to 1.12) * | 1.26 (1.18 to 1.35) * |
| Mixed | 1.16 (0.99 to 1.34) | 1.06 (1.03 to 1.10) * | 1.15 (0.99 to 1.33) |
| Other | 1.05 (0.94 to 1.17) | 1.06 (1.04 to 1.09) * | 1.06 (0.95 to 1.18) |
| Unknown | 1.00 (0.95 to 1.04) | 1.06 (1.05 to 1.07) * | 1.07 (1.02 to 1.11) * |
| **Smoking status** |  |  |  |
| Current (Ref.) |  |  |  |
| Never | 1.09 (1.05 to 1.12) * | 1.11 (1.10 to 1.12) * | 1.12 (1.08 to 1.16) * |
| Former | 1.05 (1.02 to 1.09) * | 1.07 (1.07 to 1.08) * | 1.06 (1.02 to 1.09) * |
| Unknown | 0.94 (0.80 to 1.10) | 0.66 (0.64 to 0.69) * | 1.05 (0.90 to 1.22) |
| **Drinking status** |  |  |  |
| Current (Ref.) |  |  |  |
| Never | 0.92 (0.89 to 0.95) * | 0.93 (0.92 to 0.94) * | 0.90 (0.87 to 0.93) * |
| Former | 0.91 (0.81 to 1.01) | 0.92 (0.90 to 0.94) * | 0.88 (0.79 to 0.98) * |
| Unknown | 0.96 (0.93 to 0.99) * | 0.94 (0.93 to 0.94) * | 0.99 (0.97 to 1.02) |
| **Opioid characteristics** |  |  |  |
| Average MME/day | 0.98 (0.98 to 0.98) * | 0.99 (0.99 to 0.99) * | 1.01 (1.01 to 1.01) * |
| Strong and Long-acting opioids (Ref.) |  |  |  |
| Weak and Short-acting opioids | 1.50 (1.39 to 1.61) * | 1.53 (1.51 to 1.55) * | 1.36 (1.26 to 1.49) * |
| Strong and Short-acting opioids | 1.64 (1.51 to 1.77) * | 1.36 (1.33 to 1.38) * | 1.25 (1.10 to 1.41) * |
| Weak and Long-acting opioids | 1.66 (1.26 to 2.19) * | 1.29 (1.22 to 1.35) * | 0.99 (0.73 to 1.34) |
| **Region** |  |  |  |
| London (Ref.) |  |  |  |
| North East | 0.80 (0.72 to 0.89) * | 0.90 (0.88 to 0.91) * | 0.79 (0.74 to 0.85) * |
| North West | 0.85 (0.80 to 0.91) * | 0.91 (0.90 to 0.92) * | 0.90 (0.86 to 0.94) * |
| Yorkshire & the Humber | 0.85 (0.76 to 0.95) * | 0.90 (0.88 to 0.91) * | 0.82 (0.77 to 0.88) * |
| East Midlands | 0.94 (0.83 to 1.07) | 0.92 (0.90 to 0.94) * | 0.92 (0.85 to 0.99) * |
| West Midlands | 0.89 (0.84 to 0.95) * | 0.93 (0.92 to 0.94) * | 0.89 (0.85 to 0.93) * |
| East of England | 1.00 (0.90 to 1.12) | 0.95 (0.93 to 0.96) * | 1.06 (0.99 to 1.13) |
| South West | 0.95 (0.88 to 1.02) | 0.96 (0.95 to 0.98) * | 0.97 (0.92 to 1.01) |
| South Central | 0.89 (0.82 to 0.96) * | 0.96 (0.95 to 0.97) * | 0.90 (0.86 to 0.94) * |
| South East Coast | 0.90 (0.83 to 0.97) * | 0.95 (0.94 to 0.96) * | 0.91 (0.86 to 0.96) * |
| **IMD** |  |  |  |
| Q5 (most deprived, Ref.) |  |  |  |
| Q4 | 1.04 (1.00 to 1.08) * | 1.05 (1.04 to 1.05) * | 1.06 (1.03 to 1.10) * |
| Q3 | 1.06 (1.02 to 1.10) * | 1.08 (1.07 to 1.09) * | 1.14 (1.10 to 1.19) * |
| Q2 | 1.05 (1.01 to 1.10) * | 1.11 (1.10 to 1.12) * | 1.15 (1.11 to 1.19) * |
| Q1 (least deprived) | 1.09 (1.04 to 1.14) * | 1.16 (1.15 to 1.17) * | 1.25 (1.20 to 1.30) * |
| Unknown | 1.11 (1.01 to 1.23) * | 1.12 (1.10 to 1.14) * | 1.11 (1.02 to 1.21) * |
| **Comorbidities** |  |  |  |
| RA | 0.91 (0.85 to 0.97) * | 0.89 (0.87 to 0.90) * | 0.77 (0.69 to 0.87) * |
| OA | 1.06 (1.02 to 1.11) * | 1.06 (1.05 to 1.07) * | 1.06 (1.02 to 1.11) * |
| Anxiety | 1.04 (1.01 to 1.07) * | 1.02 (1.02 to 1.03) * | 1.12 (1.07 to 1.18) * |
| Depression | 1.02 (0.99 to 1.05) | 0.97 (0.96 to 0.98) * | 0.98 (0.95 to 1.01) |
| Schizophrenia | 0.88 (0.81 to 0.96) * | 0.94 (0.92 to 0.95) * | 0.88 (0.81 to 0.95) * |
| Epilepsy | 0.92 (0.86 to 0.99) * | 0.91 (0.90 to 0.93) * | 0.89 (0.83 to 0.95) * |
| Substance misuse disorder | 0.77 (0.72 to 0.83) * | 0.82 (0.81 to 0.83) * | 0.72 (0.67 to 0.78) * |
| Alcohol dependence | 0.88 (0.82 to 0.94) * | 0.91 (0.90 to 0.92) * | 0.87 (0.82 to 0.93) * |
| **CCI** |  |  |  |
| High (>=5, Ref.) |  |  |  |
| Medium (3-4) | 1.04 (0.94 to 1.15) | 0.99 (0.97 to 1.01) | 1.18 (1.08 to 1.30) * |
| Low (0-2) | 1.05 (0.89 to 1.24) | 1.04 (0.97 to 1.08) | 1.37 (1.17 to 1.62) * |
| **Concurrent medications** |  |  |  |
| Gabapentinoids | 0.83 (0.69 to 1.00) * | 0.91 (0.88 to 0.94) * | 0.51 (0.36 to 0.71) * |
| Antidepressants | 0.89 (0.87 to 0.92) * | 0.89 (0.89 to 0.90) * | 0.91 (0.87 to 0.95) * |
| Muscle relaxants | 1.04 (0.94 to 1.16) | 0.99 (0.97 to 1.02) | 1.17 (1.06 to 1.29) * |
| NSAIDs | 1.06 (1.03 to 1.09) * | 1.07 (1.06 to 1.08) * | 1.03 (1.00 to 1.06) * |
| Benzodiazepines | 1.08 (1.05 to 1.11) * | 1.00 (0.99 to 1.01) | 1.20 (1.14 to 1.26) * |
| Z-drugs | 0.97 (0.93 to 1.01) | 0.93 (0.92 to 0.94) * | 0.95 (0.92 to 0.99) * |
| **tvc** |  |  |  |
| Age | - | - | 1.00 (1.00 to 1.00) * |
| Average daily dose | - | - | 1.00 (1.00 to 1.00) * |
| RA | - | - | 1.00 (0.99 to 1.00) * |
| Anxiety | - | - | 1.00 (1.00 to 1.00) * |
| Gabapentinoids | - | - | 1.01 (1.01 to 1.01) * |
| Antidepressants | - | - | 1.00 (1.00 to 1.00) * |
| Benzodiazepines | - | - | 1.00 (1.00 to 1.00) * |
| Combination | - | - | 1.00 (1.00 to 1.00) * |

Note: IMD = Index of Multiple Deprivation; RA: Rheumatoid Arthritis; OA: Osteoarthritis; CCI: Charlson Comorbidity Index; NSAIDs: Non-steroidal anti-inflammatory drug; Combination = weak and short-acting opioids or weak and long-acting opioids or strong and short-acting opioids or strong and long-acting opioids; aSHR: aHR: adjusted Hazard Ratio; adjusted Subdistribution Hazard Ratio

### **Supplementary Table S4. STROBE Statement—Checklist of items that should be included in reports of cohort studies**

|  | Item No | Recommendation | Page No |
| --- | --- | --- | --- |
| **Title and abstract** | 1 | (*a*) Indicate the study’s design with a commonly used term in the title or the abstract | 1 |
|  |  | (*b*) Provide in the abstract an informative and balanced summary of what was done and what was found | 2 |
| Introduction | | |  |
| Background/rationale | 2 | Explain the scientific background and rationale for the investigation being reported | 4 |
| Objectives | 3 | State specific objectives, including any prespecified hypotheses | 5 |
| Methods | | |  |
| Study design | 4 | Present key elements of study design early in the paper | 5 |
| Setting | 5 | Describe the setting, locations, and relevant dates, including periods of recruitment, exposure, follow-up, and data collection | 5 |
| Participants | 6 | (*a*) Give the eligibility criteria, and the sources and methods of selection of participants. Describe methods of follow-up | 5 |
|  |  | (*b*) For matched studies, give matching criteria and number of exposed and unexposed | N/A |
| Variables | 7 | Clearly define all outcomes, exposures, predictors, potential confounders, and effect modifiers. Give diagnostic criteria, if applicable | 6 |
| Data sources/ measurement | 8* | For each variable of interest, give sources of data and details of methods of assessment (measurement). Describe comparability of assessment methods if there is more than one group | 6 |
| Bias | 9 | Describe any efforts to address potential sources of bias | 7 |
| Study size | 10 | Explain how the study size was arrived at | N/A |
| Quantitative variables | 11 | Explain how quantitative variables were handled in the analyses. If applicable, describe which groupings were chosen and why | 6-7 |
| Statistical methods | 12 | (*a*) Describe all statistical methods, including those used to control for confounding | 7 |
|  |  | (*b*) Describe any methods used to examine subgroups and interactions | N/A |
|  |  | (*c*) Explain how missing data were addressed | 7 |
|  |  | (*d*) If applicable, explain how loss to follow-up was addressed | N/A |
|  |  | (*e*) Describe any sensitivity analyses | 7 |
| Results | | |  |
| Participants | 13* | (a) Report numbers of individuals at each stage of study—eg numbers potentially eligible, examined for eligibility, confirmed eligible, included in the study, completing follow-up, and analysed | 8 |
|  |  | (b) Give reasons for non-participation at each stage | Supplementary material p84 |
|  |  | (c) Consider use of a flow diagram | Supplementary material p82 |
| Descriptive data | 14* | (a) Give characteristics of study participants (eg demographic, clinical, social) and information on exposures and potential confounders | 8 |
|  |  | (b) Indicate number of participants with missing data for each variable of interest | 22-23 |
|  |  | (c) Summarise follow-up time (eg, average and total amount) | N/A |
| Outcome data | 15* | Report numbers of outcome events or summary measures over time | 8 |
| Main results | 16 | (*a*) Give unadjusted estimates and, if applicable, confounder-adjusted estimates and their precision (eg, 95% confidence interval). Make clear which confounders were adjusted for and why they were included | 8-9 |
|  |  | (*b*) Report category boundaries when continuous variables were categorized | 22-23 |
|  |  | (*c*) If relevant, consider translating estimates of relative risk into absolute risk for a meaningful time period | N/A |
| Other analyses | 17 | Report other analyses done—eg analyses of subgroups and interactions, and sensitivity analyses | 10 |
| Discussion | | |  |
| Key results | 18 | Summarise key results with reference to study objectives | 10-11 |
| Limitations | 19 | Discuss limitations of the study, taking into account sources of potential bias or imprecision. Discuss both direction and magnitude of any potential bias | 14-15 |
| Interpretation | 20 | Give a cautious overall interpretation of results considering objectives, limitations, multiplicity of analyses, results from similar studies, and other relevant evidence | 11-14 |
| Generalisability | 21 | Discuss the generalisability (external validity) of the study results | 15 |
| Other information | | |  |
| Funding | 22 | Give the source of funding and the role of the funders for the present study and, if applicable, for the original study on which the present article is based | 17-18 |

*Give information separately for exposed and unexposed groups.

**Note:** An Explanation and Elaboration article discusses each checklist item and gives methodological background and published examples of transparent reporting. The STROBE checklist is best used in conjunction with this article (freely available on the Web sites of PLoS Medicine at http://www.plosmedicine.org/, Annals of Internal Medicine at http://www.annals.org/, and Epidemiology at http://www.epidem.com/). Information on the STROBE Initiative is available at http://www.strobe-statement.org.

### **Supplementary Figure S1. Flowchart of identification of study cohort**

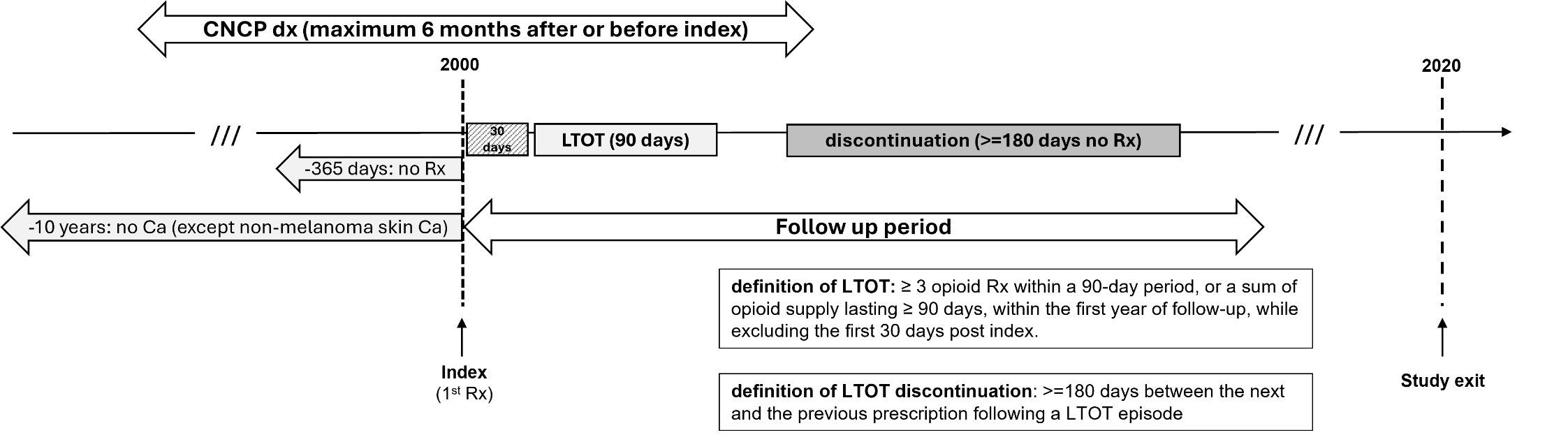

Note: dx=diagnosis; Rx=description; Ca=cancer

### **Supplementary Figure S2. Decisions made for opioid drug preparation to derive daily dose**

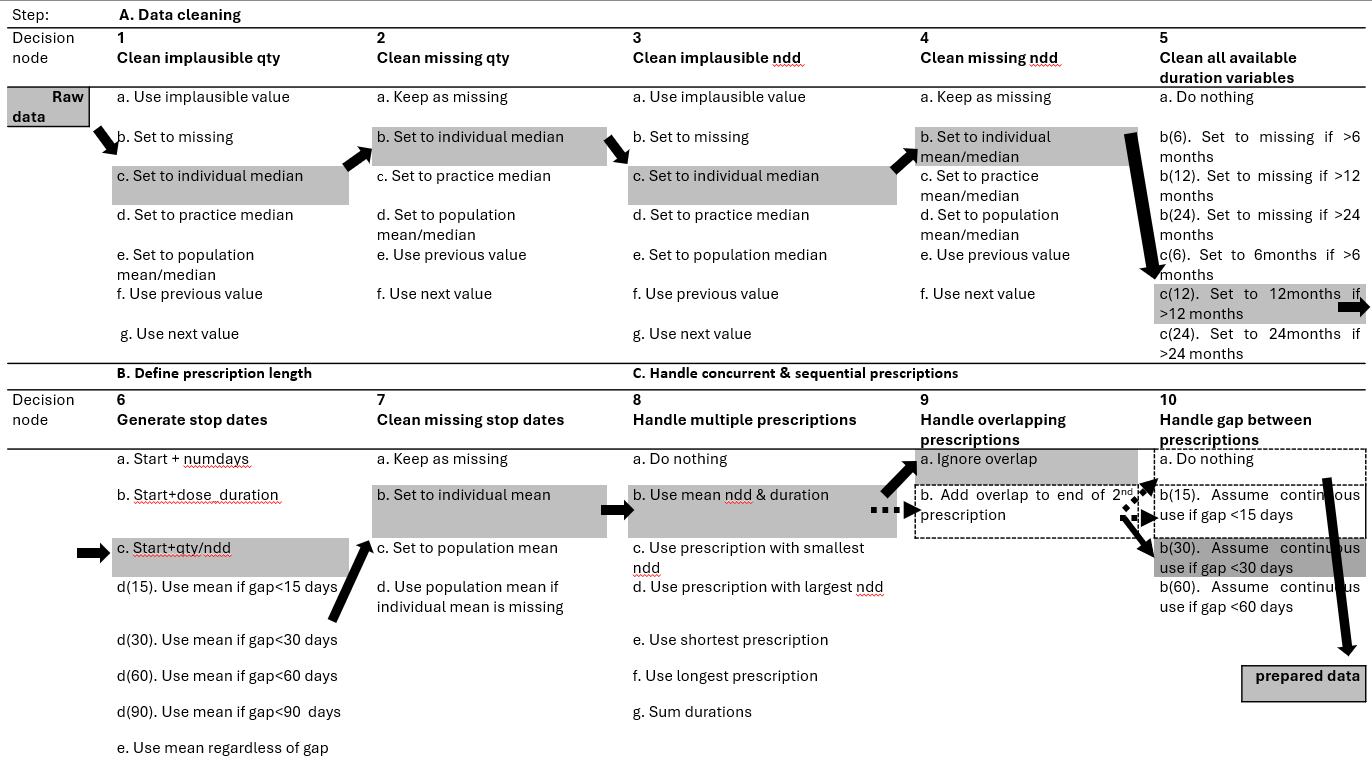

Note: qty=quantity; ndd=daily dose; numdays=number of days; The decisions in the dark grey boxes were made for the primary analysis, while those in the dashed boxes were used for the sensitivity analyses.

### **Supplementary Figure S3. Flowchart of identification of L-TOT users and L-TOT discontinuers**

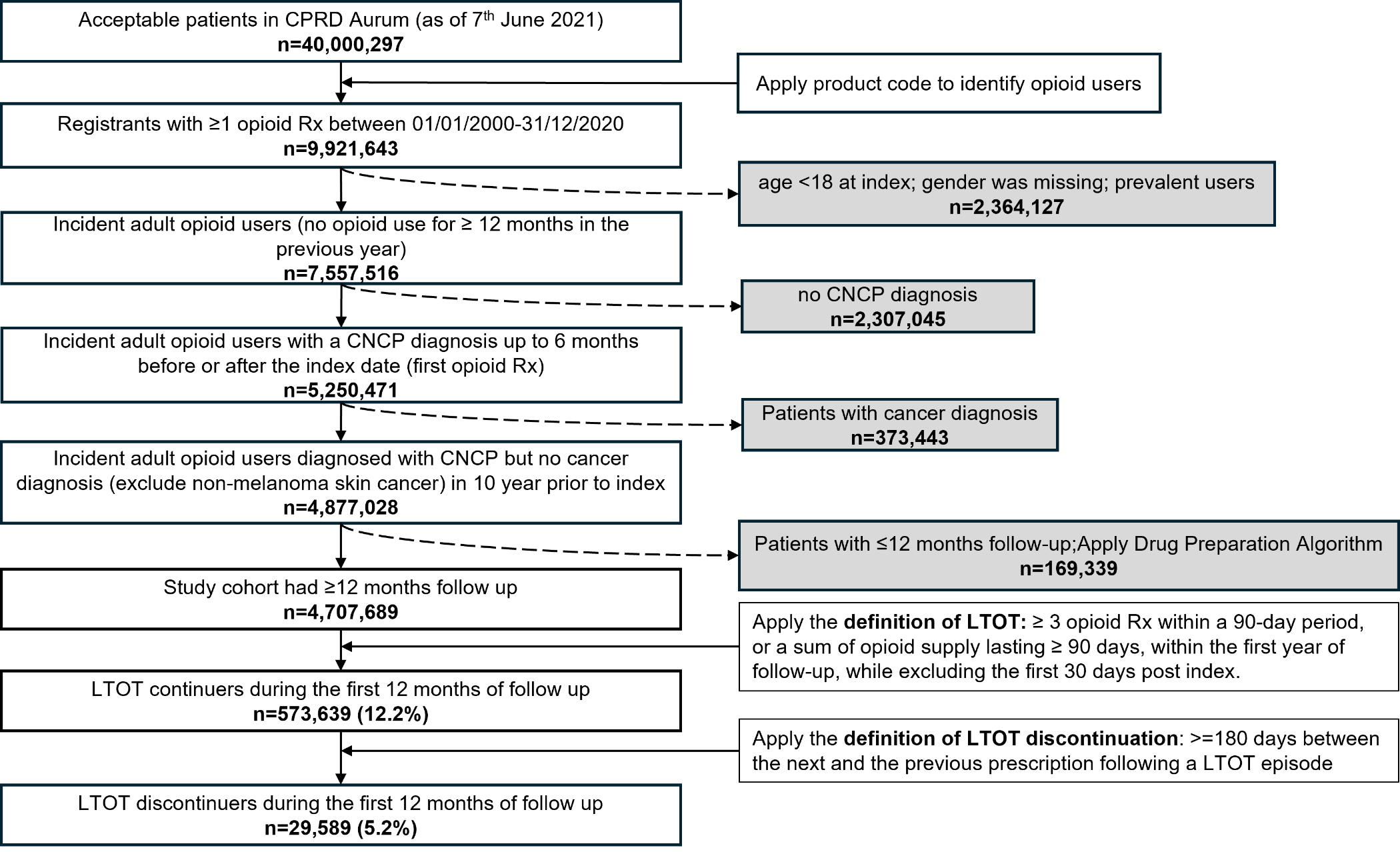
